# Modelling hepatitis C infection acquired from blood transfusions in the UK between 1970 and 1991 for the Infected Blood Inquiry

**DOI:** 10.1101/2023.11.03.23298029

**Authors:** Sarah Hayes, Ruth McCabe, Daniela De Angelis, Christl A Donnelly, Stephen JW Evans, Graham F Medley, David J Spiegelhalter, Sheila M Bird

## Abstract

The Statistics Expert Group was convened at the request of the Infected Blood Inquiry to provide estimates of the number of infections and deaths from bloodborne infections including hepatitis B virus; human immunodeficiency virus; hepatitis C virus and variant Creutzfeldt Jakob disease, as a direct result of contaminated blood and blood products administered in the United Kingdom of Great Britain and Northern Ireland (UK). In the absence of databases of HCV infections and related deaths for all nations of the UK, a statistical model was required to estimate the number of infections and subsequent deaths from HCV acquired from blood transfusions throughout January 1970 – August 1991.

We present this statistical model in detail alongside the results of its application to each of the four nations in the UK. We estimated that 26,800 people (95% uncertainty interval 21,300 – 38,800) throughout the UK were chronically infected with HCV because of contaminated blood transfusions between January 1970 and August 1991. The number of deaths up to the end of 2019 that occurred as a result of this chronic infection is estimated to be 1,820 (95% uncertainty interval 650 – 3,320).

## Introduction

The Infected Blood Inquiry (IBI), which became officially established on 2 July 2018 at the request of the Prime Minister (1) after years of campaigning by those affected (2), was tasked to “examine the circumstances in which men, women and children treated by National Health Services in the United Kingdom were given infected blood and infected blood products, in particular since 1970” (3). During an emergency parliamentary debate in the UK House of Commons in 2017, this incident was referred to as “the worst treatment disaster in the history of our NHS, and one of the worst peacetime disasters ever to take place in this country” (4). The IBI was guided by a List of Issues (5), which included ascertainment of the magnitude of infections and deaths from bloodborne infections including hepatitis B virus (HBV); human immunodeficiency virus (HIV); hepatitis C virus (HCV) and variant Creutzfeldt Jakob disease (vCJD), as a direct result of treatment with contaminated blood and blood products.

Each of these infections can cause substantial morbidity and may result in death among those infected, even when appropriate treatment is given. For HIV, a decade-long interval from infection to HIV-related symptoms (6), and a relatively quickly developed antibody test meant that registers of diagnosed infections could be developed and most of the HIV ascertainment questions posed by the IBI could be answered by analysis of such confidential databases held by organisations such as the United Kingdom Health Security Agency (UKHSA), Public Health Scotland (PHS) and the United Kingdom Haemophilia Centre Doctors’ Organisation (UKHCDO) (7). HCV, on the other hand, had been around for a long time as non-A, non-B hepatitis before being identified. Also, its natural history is more complex: around a quarter of those infected spontaneously clear the infection while the rest develop chronic HCV infection which may remain clinically silent for up to 20 years (8) and can ultimately be responsible for hepatocellular cancer, liver failure and other serious conditions (9,10). The long incubation period for chronic HCV means that many individuals infected with HCV through transfusion of contaminated blood may never have been aware of their infection and would not feature on a registry. Those that were diagnosed with HCV may have had their diagnosis and exposure-route recorded, but this was likely to be many years after the HCV-implicated transfusion. Lookback exercises, whereby individuals who have received blood from donors subsequently identified as HCV-infected are notified and advised to undergo testing, can provide some additional information, but only apply in respect to repeat blood donors. Alternative approaches to the ascertainment questions in respect to HCV were therefore required.

The Penrose Inquiry (PI), chaired by the Rt. Hon. Lord Penrose, into HCV and HIV infection acquired following treatment by NHS Scotland with contaminated blood and blood products was announced in 2008 (11). The PI lacked powers to compel evidence, applied to Scotland only and reported in 2015 and so is distinct from the IBI, although much of the scope is overlapping (12). For the PI, Schnier and Goldberg (13) developed a statistical model to estimate the number of people who acquired HCV infection as a consequence of blood transfusion in Scotland between 1970 and 1991 and, of those, the number alive in 2011. These estimates provided the first quantification of the magnitude of this incident in Scotland.

For the IBI, a statistical model building on the work of Schnier and Goldberg (13) was developed to estimate the number of people infected with, and those who subsequently died from, HCV transmitted from blood transfusions throughout the United Kingdom of Great Britain and Northern (UK) across January 1970 – August 1991. Reliable screening of blood donations for HCV antibodies commenced in September 1991, preventing HCV infections from contaminated blood after this date. This paper presents this statistical model and associated results, as submitted as evidence to the IBI in September 2022 (7). Whilst there is inevitable overlap with the relevant chapter of the published report, this paper serves to present the model in detail and provide results to the wider scientific community.

## Methods

### The Statistics Expert Group

The Hon. Mr Justice Langstaff (Sir Brian) was appointed to chair the IBI sitting on his own and opted to convene a series of Expert Groups to provide evidence on specific topics related to the List of Issues published by the IBI. The Statistics Expert Group (SEG) (14) was convened to provide expert statistical advice to the IBI in accordance with a Letter of Instruction in 2019 (15), focussing on estimation of the number of people infected with and who subsequently died from infection with the aforementioned viruses acquired through treatment with contaminated blood or blood products. The emergence of the novel SARS-CoV-2 virus in late 2019 resulted in all SEG members becoming heavily involved in research and advisory roles in response to the subsequent pandemic. A further exchange of letters between the SEG and Sir Brian (16) resulted in agreement to limit the scope of what might be done to the highest quality within a reasonable time, allowing Sir Brian and the Inquiry to complete their work without further delays to the Inquiry’s time-frame beyond those imposed by national public health and social measures implemented to control the spread of SARS-CoV-2. Prior to (16), the record-linkage work essential for SEG’s modelling had been approved in the public interest and put in hand with support from the IBI.

### Statistical model to estimate the HCV infections and deaths from contaminated blood transfusions

A statistical model was used to provide estimates of the quantities of interest for which no direct data were available, namely the number infected and consequent deaths from HCV transmitted via blood transfusions, by leveraging the variety of data that were available, for example, on the proportion of HCV-infected blood donors at the time of introduction of testing and the annual number of blood transfusions. The model developed by Schnier and Goldberg (13) provided a clear and logical means by which to approach this question of ascertainment by breaking the overall estimation problem into six smaller, chronological tasks, as described below and outlined in Figure 1. The output of each task was then used as input into the subsequent task. Each task was dependent on several assumptions and often contained further subtasks which also required statistical modelling. An overview of the model assumptions and inputs can be found in Table 1 and Table 2, respectively, and extensive further information is provided in the Supplementary Information, which derives from the SEG Report (7).

**Figure 1:**
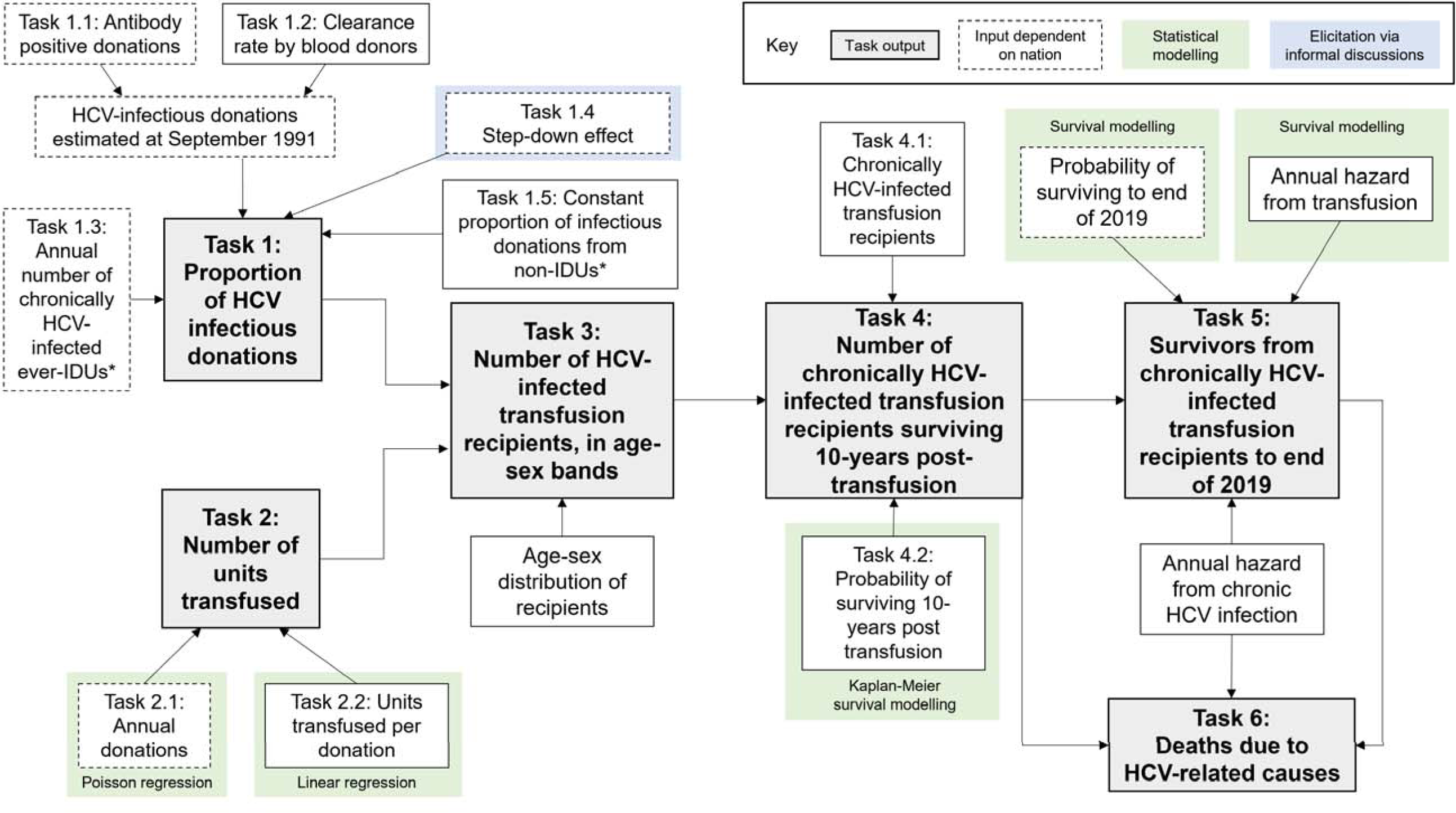
Structure of model used to estimate HCV infections following blood transfusions based on the model developed by Schnier and Goldberg (13) for the Penrose Inquiry. The model is broken into six tasks, shown in grey boxes with black outline, with each preceding task informing the subsequent tasks. This model structure is adapted to the four nations through the use of different inputs: white boxes with a solid black line indicate model inputs which are the same across the four nations and white boxes with a dotted black line indicate a nation-specific input. Green boxes indicate the use of further statistical modelling ahead of input to the model, and the blue box indicates information from our elicitation exercise. All other inputs came from the literature. Note that from Task 3, outputs of all tasks are age-sex disaggregated. Further details of the model can be found in Table 1 and Table 2. *Abbreviations: Injecting drug users (IDUs).

**Table 1:** Overview of the list of assumptions in the statistical model.

| Assumption label | Task | Details |
| --- | --- | --- |
| a | 1 | Prevalence of HCV-infectious donations is estimated using a 'hybrid' model with two components.<br>Component 1 is proportional to chronic HCV prevalence in past or current injecting drug users (ever-IDUs) in the year of transfusion.<br>Component 2 comes from non-IDUs and is set at a constant level. |
| b | 1.2 | A constant proportion of those donors previously infected with HCV were actively HCV-infectious and so would transmit the virus in their blood. The proportion is assumed to be 74% (95% CI 71% – 78%). |
| c | 1.4 | In addition to HIV antibody screening of donors, guidance to prospective donors in the mid-1980s led to a substantial fall (deferral) in HCV-infectious donations. The baseline assumption is that this occurred in 1985 in England, Wales and Northern Ireland, and in 1984 in Scotland. |
| d | 1.4 | Self-deferral led to around a 67% reduction in donations from HCV-infectious ever-IDUs. |
| e | 1.5 | The contribution to prevalence from non-IDUs (component 2) is a fixed percentage of the proportion of donations that were found to be HCV-infectious when testing began in 1991. The baseline assumption for this percentage is 25%. |
| f | 3 | All recipients of HCV-infectious units are assumed to develop HCV infection. |
| g | 3 | There is a negligible chance of a transfusion recipient receiving two infected units. |
| h | 4.1 | A proportion (18% (95% CI 13% – 24%)) of transfusion recipients with HCV infection is assumed to clear the virus within 6 months of acquisition. |
| i | 4.2 | Chronic HCV infection does not influence recipients' survival for the first 10 years post-transfusion. |
| j | 5 | Having a transfusion is associated with an increase in the annual risk of dying up to 20 years after transfusion. |
| k | 6 | Chronic HCV infection increases the annual risk of dying from 10 years post-infection: the baseline assumption is a 53% (95% CI 17% – 100%) increased risk. |

**Table 2:** Model inputs including probabilistic distributions for each of the four nations of the UK. Where 95% confidence intervals/uncertainty intervals are extracted from published studies or model estimates, the standard deviation is estimated by dividing the width of the interval by (1.96 X 2). Abbreviations: Infected Blood Inquiry (IBI); Public Health Laboratory Service (PHLS); United Kingdom Health Security Agency (UKHSA); National Blood Transfusion Service (NBTS); Office for National Statistics (ONS).

| Task | Input | Source |  |  |  |
| --- | --- | --- | --- | --- | --- |
|  |  | England | Northern Ireland | Scotland | Wales |
| 1.1 | 1991 HCV-infected donor prevalence | Central value: 0.066%<br>Uncertainty:<br>Beta(532,808938) | Central value: 0.066%<br>Uncertainty:<br>Beta(532,808938) | Central value:<br>0.088%<br>Uncertainty:<br>Beta(159, 180,000) | Central value: 0.066%<br>Uncertainty:<br>Beta(532,808938) |
|  |  | *Soldan (18)* | *Assumed to be as in England and Wales* | *Schnier and Goldberg (13)* | *Soldan (18)* |
| 1.2 | HCV-infectious donations | Central value: 26%<br>Normal(0.26,0.018) where 0.26 represents the mean (reported central estimate) and 0.018 the reported standard deviation. |  |  |  |
|  |  | *Micallef et al. (8)* |  |  |  |
| 1.3 | Number of HCV-infectious ever-IDUs | Randomly sample one set of figures from the 1000 estimates of the number of IDUs provided. | Randomly sample one set of figures from the 1000 estimates of the number of IDUs provided. | No uncertainty provided and so values taken directly as presented in the source. | Randomly sample one set of figures from the 1000 estimates of the number of IDUs provided. |
|  |  | *UKHSA modelling (unpublished)* | *UKHSA modelling (unpublished)* | *Schnier and Goldberg (13)* | *UKHSA modelling (unpublished)* |
| 1.4 | Year of deferral reduction | 1985 | 1985 | 1984 | 1985 |
|  |  | *Informal discussions with members of IBI Expert Groups* | *Informal discussions with members of IBI Expert Groups* | *Schnier and Goldberg (13)* | *Informal discussions with members of IBI Expert Groups* |
| 1.4 | Effect of deferral reduction | Central value: 67%<br>Uncertainty: Normal (0.67, 0.0765) | Central value: 67%<br>Uncertainty: Normal (0.67, 0.0765) | Central value: 67% | Central value: 67%<br>Uncertainty: Normal (0.67, 0.0765) |
|  |  | *Informal discussions with members of Inquiry Expert Groups* | *Informal discussions with members of Inquiry Expert Groups* | *Schnier and Goldberg (13)* | *Informal discussions with members of Inquiry Expert Groups* |
| 1.5 | Percentage | 25% |  |  |  |
|  | contribution to prevalence from non-IDUs | *Based on judgement (and sensitivity analysis reveals that the precise value has little impact).* |  |  |  |
| 2.1 | Number of blood donations | Supplementary Table 5 |  |  |  |
|  |  | *NBTS (22,23)* | *Pro-rata from NBTS (22,23)* | *Schnier and Goldberg (13)* | *NBTS (22,23)* |
| 2.2 | Units used per donation | Supplementary Table 7<br>For each year: Normal (mean, standard deviation) where the mean is taken from a linear regression model and the standard deviation is estimated by treating the 95% prediction intervals as the 95% uncertainty intervals.* |  |  |  |
|  |  | *National Blood Transfusion Service Statistics (24)* |  |  |  |
| 3 | Number of antibody positive donations | For each year: Binomial (number of donations, antibody prevalence in donors) |  |  |  |
| 3 | Number of RNA positive donations | For each year: Binomial (number of antibody positive donations, 1 – HCV clearance in donors) |  |  |  |
| 3 | Number RNA positive units transfused (number people infected) | For each year: Number RNA positive donations multiplied by units transfused from each donation |  |  |  |
| 3 | Age-sex distribution of recipients | Supplementary Table 12<br>Dirichlet (Proportion of people in each age-sex band) |  |  |  |
|  |  | *Wallis et al. (25)* |  |  |  |
| 3 | Number infected by age and sex in each year | Multinomial (number infected per year, proportion of recipients by age and sex) |  |  |  |
| 4.1 | Percentage of people infected with HCV who naturally clear the virus | Central value: 18%<br>Uncertainty: Normal (0.18,0.028) where 0.18 is the mean and 0.028 is the standard deviation. |  |  |  |
|  |  | *Micallef et al. (8)* |  |  |  |
| 4.1 | Number of people chronically infected | Binomial (number of people infected, 1 – HCV clearance in survivors) |  |  |  |
| 4.2 | Survival to 10 years post-transfusion | Supplementary Table 16 |  |  |  |
|  |  | *Wallis et al. (25) and Morley et al. (27)* |  |  |  |
| 5 | Probability of surviving until the end of 2019 | *ONS life-tables for England (32)* | *ONS life-tables for Northern Ireland (33)* | *ONS life-tables for Scotland (34)* | *ONS life-tables for Wales (35)* |
| 5 | Additional risk to survival from having a transfusion | Supplementary Table 28<br>*Scotland NBTS (31)* |  |  |  |
| 6 | Additional risk to survival from chronic HCV infection | Central estimate: 1.53<br>Uncertainty: Exponential value selected on log scale from Normal(0.425, 0.137) matching estimated hazard ratio of 1.53 (95% interval 1.17 – 2.00).<br>*UKHSA modelling (unpublished)* |  |  |  |

A separate model was applied to each of the four nations of the UK (ordered throughout alphabetically: England, Northern Ireland, Scotland and Wales), incorporating setting-specific information where possible. The metrics of particular interest were the number of people: infected (in total and chronically); chronically HCV-infected persons surviving 10 years post-transfusion; thereafter surviving to the end of 2019 and the number of deaths attributable to chronic HCV infection. End of December 2019 was selected as the end date given that annual hazards were substantially impacted by the COVID-19 pandemic from 2020 onwards, and to accommodate delays in death-registration outside of Scotland (17). Although Schnier and Goldberg had already produced estimates for Scotland, we were able to provide estimates for survivors to the end of 2019 rather than 2011, as presented to the PI.

First, a deterministic version of the model was developed and used to conduct extensive sensitivity analyses. Stochasticity was then incorporated using Monte Carlo simulation to propagate the uncertainty arising within each task, for example, via the statistical models deployed in the development of some subtasks, through to the final results of the model.

We now describe each of the six model tasks in detail before discussing the application of the model to each of the four nations and providing an overview of the sensitivity analyses considered.

### Task 1 What proportion of blood donations received between January 1970 – August 1991 was HCV-infectious?

The process of estimating the proportion of HCV-infectious blood donations is illustrated in Figure 2. In short, we use the observed HCV prevalence in blood donors in September 1991 and the trend in HCV prevalence in injecting drug users (IDUs) to extrapolate the annual HCV prevalence in blood donors back to 1970. We then adjust for the effect of changing donation advice and sources of HCV in blood donations other than from IDUs.

**Figure 2:**
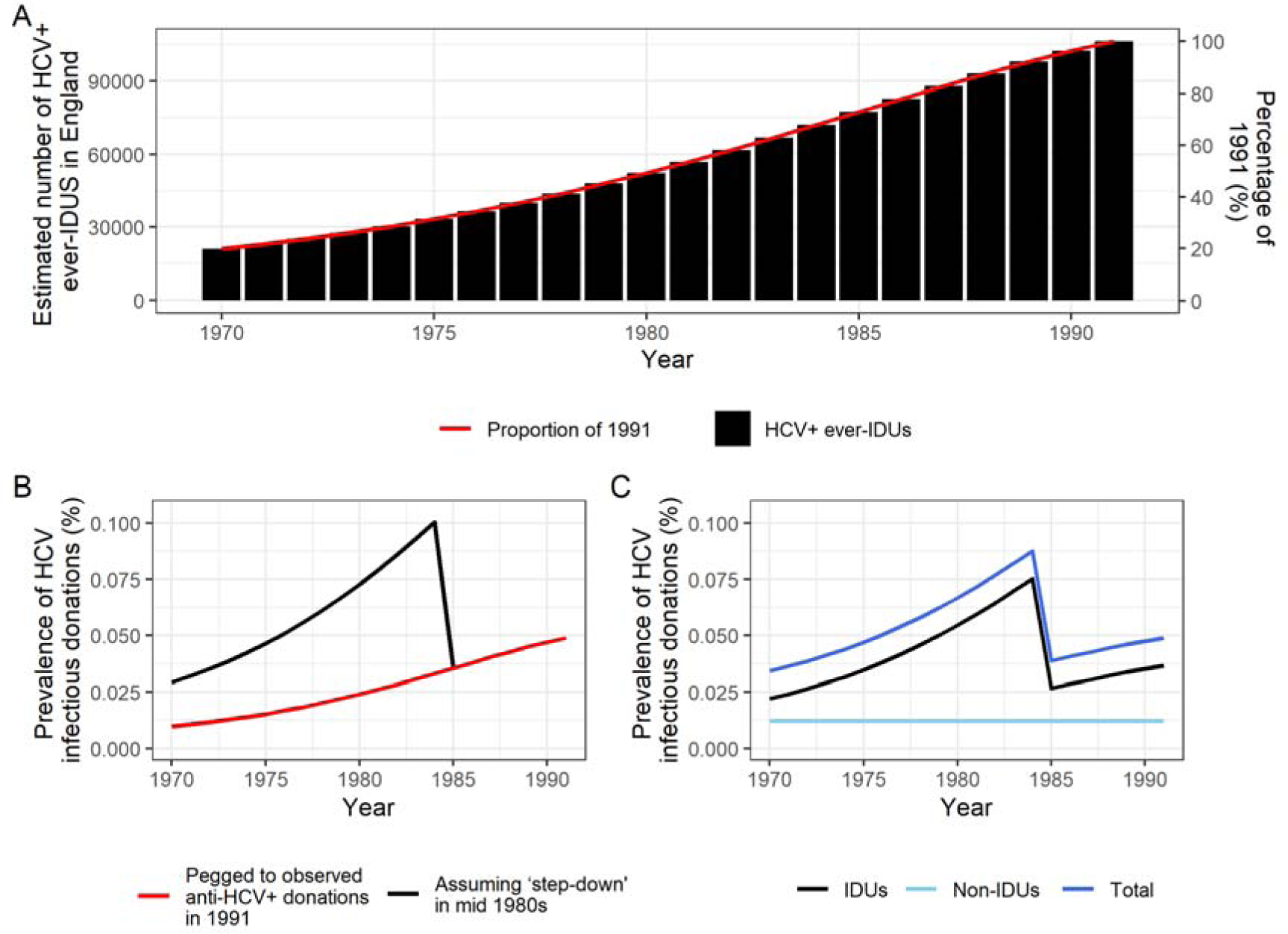
Stages in estimating the prevalence of HCV-infectious donations. Example data from England. Underlying data can be found in Supplementary Table 1. (A) Estimated number of ever-injecting drug users (ever-IDUs, black columns) and percentage that each year contributes to the value observed in 1991 (red line). (B) The trend observed in (A) is pegged to an observed value of HCV antibody positive donations in 1991 of 0.049% (red line). The black line remains pegged to the observed value in 1991 but now also incorporates an estimated effect (67%) of the deferral policy in 1985, which caused prevalence to increase beforehand (Table 4.6 column (d)). (C) The black line in (B) is adjusted to account for the scenario that 25% of the observed 1991 HCV antibody prevalence is not from IDUs and is constant across the period (light blue line). The dark blue line shows the resulting percentage of HCV antibody positive donations from these two sources.

#### Task 1.1. Just after screening of blood donors started in September 1991, what proportion of donations was HCV antibody positive?

Reliable screening of blood donations for antibodies for HCV began in September 1991, with the prevalence of HCV antibodies in blood donations unknown prior to this time. Between September and December 1991, 532 of 808,938 (0.066%, 95% confidence interval (CI) 0.060% - 0.072%) donations tested in England and Wales were found to be positive for HCV antibodies (18,19). In Scotland 159 of 180,000 (0.088%) (95% CI 0.075% - 0.103%) donors tested between September 1991 and February 1992 were HCV antibody positive (13).

#### Task 1.2. Just after HCV antibody screening of donors started in September 1991, what proportion of donors was HCV-infectious?

Of those donors that are found to be HCV antibody positive, a proportion will have cleared the virus naturally and no longer be infectious. We assume a clearance rate of 26% (95% CI 22% – 29%) within the donor population based a systematic review by Micallef et al. (8), giving an estimate of 74% (95% CI 71% – 78%) of HCV antibody-positive donors remaining infectious. We refer to these antibody positive infectious donors as chronically HCV-infected.

#### Task 1.3 How many chronically HCV-infected ever-injecting drug users (ever-IDUs) were alive during each year between 1970 and 1991?

Injecting drug use is a key risk factor for transmission of HCV (20). Schnier and Goldberg (13) assumed that HCV-infectious donations came predominantly from those who had ever injected drugs (ever-IDUs) or had been indirectly infected by ever-IDUs. The change in HCV prevalence among the donor population was assumed to be proportional to the change in the estimated number of HCV-infected ever-IDUs. We adopted that assumption in part (see Task 1.5).

Previously unpublished estimates of the number of current and past HCV-infectious IDUs and their sum (HCV-infectious ever-IDUs) for England for the period 1971-1991 were provided as 1,000 posterior correlated realizations which underpin a complex analysis of HCV prevalence and sequelae by Harris et al. (9). The estimates for Northern Ireland and Wales were extrapolated by population size from the data for England. For Scotland, estimates of HCV-infected ever-IDUs from Hutchison et al. (21), as presented in Schnier and Goldberg (13), were used.

#### Task 1.4 What was the step-down in donations from ever-IDUs due to self-deferral guidance and HIV antibody testing in the mid-1980s?

During the mid-1980s, strong self-deferral guidance was issued advising current-IDUs, and later ever-IDUs, to avoid donating blood due to the risk of HIV transmission. We assume that there was a ‘step-down’ in donations from the HCV-infectious population in response to this advice. For England, Northern Ireland and Wales, we assume the step down occurred in 1985, whilst in Scotland it was assumed to occur in 1984 as advice against donations from ever-IDUs was introduced earlier in Scotland. Across all four nations of the UK, a 67% reduction in HCV-infectious donations was assumed. There is an absence of evidence for the effect of the changed guidance and so assumptions around the impact of the altered guidance are based on the Schnier and Goldberg report (13) for Scotland and, for the other three nations of the UK, on elicitation via informal discussions with members of the IBI Expert Groups who were knowledgeable about both the contemporary circumstances and the effects of guidance on behaviour (see Supplementary Material).

#### Task 1.5. What constant level of HCV-infectious non-IDU donors might it be reasonable to assume between January 1970 and August 1991?

We relaxed the assumption made by Schnier and Goldberg (13) that chronically HCV-infected ever-IDUs would be the sole driver of HCV contamination of the blood supply. Other routes may have included iatrogenic transmission historically, UK residents being treated overseas, and immigration from countries where HCV was prevalent. We assumed that the alternative contribution to the prevalence of chronic HCV infection in the blood supply was constant between January 1970 and August 1991. A baseline value of 25% of the HCV antibody prevalence in 1991 coming from this alternative route was assumed, but values from 0 to 100% were explored as part of the sensitivity analyses (see Methods: Deterministic sensitivity analysis).

### Task 2 How many blood components were transfused?

We consider blood component units (shortened hereafter to units) to denote any labile component of a whole blood donation: for example, red blood cells, platelets, plasma or cryoprecipitate. Reliable estimates for the number of units transfused over the period of interest were not available. We thus initially estimated the number of blood donations and then estimated, on average, how many units were transfused per donation received.

#### Task 2.1. How many blood donations were there annually in England between January 1970 and August 1991?

Data on the number of donations per year for England and Wales were available for 1975 to 1976 and for 1978 to 1990 (22,23), with Poisson regression used to impute the number of donations for years for which data were missing (Supplementary Figure 1). The annual number of donations attributable to England and Wales separately was estimated by scaling the value for the number of donations by the respective population size of each country. Estimates for Northern Ireland were also produced by assuming that the annual number of donations was proportional to the population of Northern Ireland in comparison to England. For Scotland, the annual numbers of donations published by Schnier and Goldberg (13) were used. Estimates for each nation are presented in Supplementary Table 5.

#### Task 2.2. Per donation, on average how many units were transfused?

Data on the number of units transfused per donation were very limited, with records only available in England and Wales for 1982 to 1988 (24). Linear regression was thus used to obtain estimates for the years for which data were not available (Supplementary Figure 3; Supplementary Table 7).

### Task 3 How many transfusion recipients were infected with HCV?

The annual number of transfusion recipients infected with HCV was estimated by multiplying the estimated number of HCV-infectious donations for each year (Task 1) by the estimated number of units transfused for that year (Task 2) (Supplementary Tables 8 – 11). We considered that all units from HCV-infectious donors are equally infectious and that every recipient of a contaminated unit would subsequently develop HCV infection. We also assumed that there was a negligible chance of a transfusion recipient receiving two infected units. Thus, every infected unit transfused was assumed to lead to a unique infected recipient.

To stratify our estimated number of infected transfusion recipients by age- and sex-bands, we used data on the age-sex distribution of transfusion recipients provided by Wallis et al. (25) who studied 2,899 transfusions recipients in June 1994 in a single transfusion centre in the north-east of England. This distribution was applied to the estimated number of infected units transfused to give the estimates for infected transfusion recipients in each age-sex band (Supplementary Table 12).

### Task 4 How many chronically HCV-infected transfusion recipients survived 10 years post-transfusion?

#### Task 4.1 How many transfusion recipients were chronically infected?

We estimate the nominal number of chronically HCV-infected transfusion recipients were they to survive to 6 months post-transfusion. Micallef et al. (8) found that 18% (95% CI 13% - 24%) of transfusion recipients with HCV-infection clear the virus within 6 months of acquisition. Consequently, we estimate those who are chronically HCV-infected by scaling the number of people estimated to be infected by 82% (95% CI 76% – 87%) within each age-sex band.

#### Task 4.2 How many chronically HCV-infected transfusion recipients survived 10 years post transfusion?

We assume that chronic HCV infection is not associated with recipients’ survival for the first 10 years post-transfusion, as evidenced by Thein et al. (26) who reported the progression rate to cirrhosis by 20 years after HCV infection is 7% (95% CI 4% - 12%) in non-clinical settings. We accounted for this in our model by estimating the number of chronically HCV-infected individuals likely to survive 10 years post transfusion, in age-sex bands.

We began by applying Kaplan-Meier methods to the data presented in Wallis et al. (25), as used by Schnier and Goldberg (13). However, as these data only contained information up to 7 years post follow-up, data from the Epidemiology and Survival of Transfusion Recipients (EASTR) study were also considered (27). While the EASTR study included follow-up to 10 years post-transfusion, the data were from a period much later than our study period and in less granular age-bands than required in this analysis. Consequently, we extended the 5- year survival probabilities estimated using the data from Wallis et al. (25) to 10 years using the change in survival between 5 and 10 years in EASTR, quantified as the ratio of the logarithms of these two probabilities, derived under the assumption of Cox’s proportional hazard model (Supplementary Table 13; Supplementary Table 16).

We then multiplied the survival probabilities by the number of transfusion recipients with chronic HCV infections in each age-sex band to obtain our estimates of the number of chronically HCV-infected recipients surviving 10 years post-transfusion (Supplementary Figure 5; Supplementary Table 17).

### Task 5 How many chronically HCV-infected transfusion recipients who survived to 10 years post-transfusion would have survived to the end of 2019, assuming no excess risk from HCV?

The Office for National Statistics (ONS) provides the hazard, the average annual risk of dying in any year, given age at the beginning of that year, separately for males and females (28). Using these data for each nation, we estimated the probability of surviving from 10- years post-transfusion to the end of 2019 for each age-sex band conditional on their year of transfusion (Supplementary Tables 19 – 26).

Having had a blood transfusion affects a person’s longer-term survival beyond the 10 years and needed to be accounted for in this model, since the reason for having the transfusion can be associated with increased death rates (29,30).

A series of record-linkage studies needed to be established or updated to determine cohorts’ survival status to the end of December 2019. The required linkages were prioritized so that, by June 2022, the necessary data (see Task 6) were available via the UKHCDO, Scottish National Blood Transfusion Centre and UKHSA (31). Using these data, we could estimate age-sex-specific hazards between 11- and 20-years post-transfusion. Lacking evidence beyond 20 years post-transfusion, but mindful that hazard ratios decreased markedly during the second decade of follow-up, we assume that there is no residual transfusion-related hazard for those who survived for 20 years post-transfusion (Supplementary Tables 27 and 28).

Transfusion-adjusted hazard rates were then obtained by multiplying the relevant hazards from the nation-specific life-tables for 11-15 years and 16-20 years post-transfusion. The estimated number of survivors was then derived by multiplying the number of 10-year survivors by the corresponding transfusion-adjusted survival probabilities (Supplementary Tables 29 – 36).

Task 6: Of those chronically HCV-infected through transfusion between January 1970 and August 1991, how many died of HCV-related causes by the end of 2019? Finally, we consider the additional risk of death for being chronically HCV-infected via transfusion.

Colleagues from UKSHA undertook a case-control study (not yet publicly available at time of writing) to estimate the additional annual hazard incurred from HCV infection. Participants in this study were individuals who had received HCV-implicated transfusions in England, for example, from a repeat donor who had tested HCV-antibody positive but not necessarily HCV-infectious. Recipients were identified and traced in in the 1990s, meaning that the traced individuals were survivor-selected and hence typically younger. The aforementioned traced recipients who consented were subsequently tested for HCV and became participants. The hazard ratio (HR) of interest could be derived from comparison of those who were HCV-RNA-positive (924 participants) versus those who were HCV-RNA-negative (443 participants). The analysis for IBI was left-truncated to allow for survivor-selection so that person-years of follow-up in the first decade post-transfusion were minimal. Hence, the resulting HR pertains essentially to follow-up after the first decade.

This study produced a HR of 1.53 (95% CI 1.17 – 2.00) (equivalent to an increased risk of 53% (95% CI (17% - 100%)) for all-cause mortality among HCV-infectious patients compared to transfused controls who were not HCV-infected. This estimate remained constant across three decades of follow up and, as such, we applied this hazard ratio to the annual, transfusion-adjusted mortality hazards as described previously within each age-sex band every year from 10 years post-transfusion (Supplementary Tables 38 – 45).

The attributable fraction (AF) among those exposed to risk is denoted:

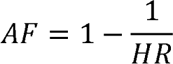

Therefore, for HR = 1.53, we would assume the fraction of deaths attributable to chronic HCV infection 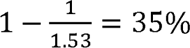.

This allowed us to estimate the number of deaths attributable to HCV via:

*Number of deaths attributable to HCV*

*= (Number chronically infected*

*- Number surviving to 2019 under HCV and transfusion - adjusted hazards)*

*- (Number chronically infected*

*- Number chronically infected surviving 10 years post transfusion)] x AF*

*= Number chronically infected surviving 10 years post transfusion -*

*Number surviving to 2019 under HCV and transfusion -*

*adjusted hazards]* x *AF.*

### Application to each nation

The model structure was applied to each nation within the UK, with the only differences coming from the data inputs. The majority of parameters and assumptions was applicable to each nation, with only a handful of information required to adapt the model to each setting (Figure 1; Table 2). Where data were missing for Wales or Northern Ireland, values were assumed to be proportional by population to those observed for England (as estimates for all parameters were available for England and the trend in IDUs in England was considered more likely to reflect that in Wales and Northern Ireland than the trend in IDUs in Scotland). Even the number of blood donations in Northern Ireland had to be estimated by assuming population pro-rata of Northern Ireland in comparison to England.

The baseline model is a stochastic model and thus inputs for each parameter were selected from a specified distribution on each run of the model. The model was run 10,000 times and with results presented throughout as median and 95% uncertainty intervals. The distributions for each input parameter used for each nation are shown in Table 2. For each of the 10,000 runs of the stochastic model for England, the number of HCV-infectious IDUs was randomly sampled from one of 1,000 realizations, previously unpublished, estimated for a vector of 22 per-annum infectious IDUs distributions provided by UKHSA. For Scotland, the estimates for the number of HCV-infectious IDUs were taken from Schnier and Goldberg (13) and thus did not incorporate any uncertainty.

#### Estimates for the UK

For the total for the UK, we added the unrounded medians for the four nations, and the uncertainty as a weighted average of the constituent nations, as follows. The stochastic uncertainties for the four nations depended on essentially the same quantities, and so were almost perfectly correlated. We defined the ’multipliers’ as the ratios of the upper and lower ends of the interval to the median, and assumed the logarithms of UK multipliers were the average of the logarithms of the nation-specific multipliers, weighted by their medians.

### Deterministic sensitivity analysis

The model is based on several assumptions. To test their suitability and influence on our results, extensive sensitivity analyses were conducted on several key assumptions. Although every subset of variable combinations was considered, attention is focussed on nine principal scenarios as outlined in Table 3.

**Table 3:** Details of the 9 principal sensitivity analysis scenarios considered. All other variables held constant at the corresponding baseline value as detailed in Table 2. Assumptions are set out in full in Table 1.

| Label | Scenario | Nations | Assumption(s) under consideration | Variable | Value |
| --- | --- | --- | --- | --- | --- |
| A | The first component in HCV-infectious donations comprise past-IDUs rather than ever-IDUs, the deferral occurred in 1987 and led to only a 33% reduction in infectious donations. | England<br>Wales<br>Northern Ireland | a | IDUs | Past-IDUs |
|  |  |  | c | Year of deferral | 1987 |
|  |  |  | d | Impact of deferral | 33% |
| B | There was no second component of non-IDU infectious donors. | All | e | Proportion from non-IDUs | 0% |
| C | There was no IDU component to the model. | All | e | Proportion from non-IDUs | 100% |
| D | The deferral occurred one year later. | England<br>Wales<br>Northern Ireland | c | Year of deferral | 1986 |
|  |  | Scotland |  |  | 1985 |
| E | The deferral effect was lower than supported by some of the expert respondents. | All | c | Impact of deferral | 33% |
| F | There was a higher contribution to prevalence from non-IDUs. | All | e | Proportion from non-IDUs | 50% |
| G | There was no additional risk to the annual hazard of dying from having had a transfusion after survival to 10-years. | All | j | Hazard ratio arising from transfusion | 1 |
| H | There was a non-age-stratified risk to the annual hazard of dying from having had a transfusion after survival to 10-years. | All | j | Hazard ratio arising from transfusion | Non-age-stratified |
| I | There was no increase in the annual risk of dying from HCV after survival to 10-years. | All | k | Hazard ratio arising from chronic HCV-infection | 1 |

## Results

This section focuses on the main quantities of interest in each of the four nations. The results of each task are presented in the Supplementary Material.

### Infections and attributable deaths in the UK

We estimate that 22,000 people (95% uncertainty interval 17,300 – 31,900) throughout the UK were chronically HCV-infected because of contaminated blood transfusions between January 1970 and August 1991. The number of deaths up to the end of 2019 that occurred as a result of this chronic infection is estimated to be 1,820 (95% uncertainty interval 650 – 3,320). The results for the six main quantities of interest for the UK are shown in Table 4. Estimates of the number of chronic HCV infections and number of HCV-related deaths by age-sex band for each of the four nations of the UK are shown in Table 5 and Table 6, respectively.

**Table 4:** Median estimates and 95% uncertainty intervals of the main quantities of interest for the UK from the statistical model of HCV infections from blood transfusions.

| Quantity of Interest | Median estimate | 95% Uncertainty interval |
| --- | --- | --- |
| Number of people infected with HCV through blood transfusion between January 1970 and August 1991 | 26,800 | 21,300 – 38,800 |
| Number chronically HCV-infected (were they to survive 6 months post-transfusion) | 22,000 | 17,300 – 31,900 |
| Number chronically HCV-infected who survived to 10 years after transfusion | 8,120 | 6,330 – 11,900 |
| Number chronically HCV-infected and survived to the end of 2019 (assuming extra HCV risk) | 2,700 | 2,050 – 3,910 |
| Number chronically HCV-infected and died by the end of 2019 | 19,300 | 15,100 – 28,200 |
| Number chronically HCV-infected who survived to 10-years post-transfusion but died before the end of 2019 | 5,420 | 4,280 – 7,990 |
| Number of deaths by the end of 2019 related to HCV infection | 1,820 | 650 – 3,320 |

**Table 5:**
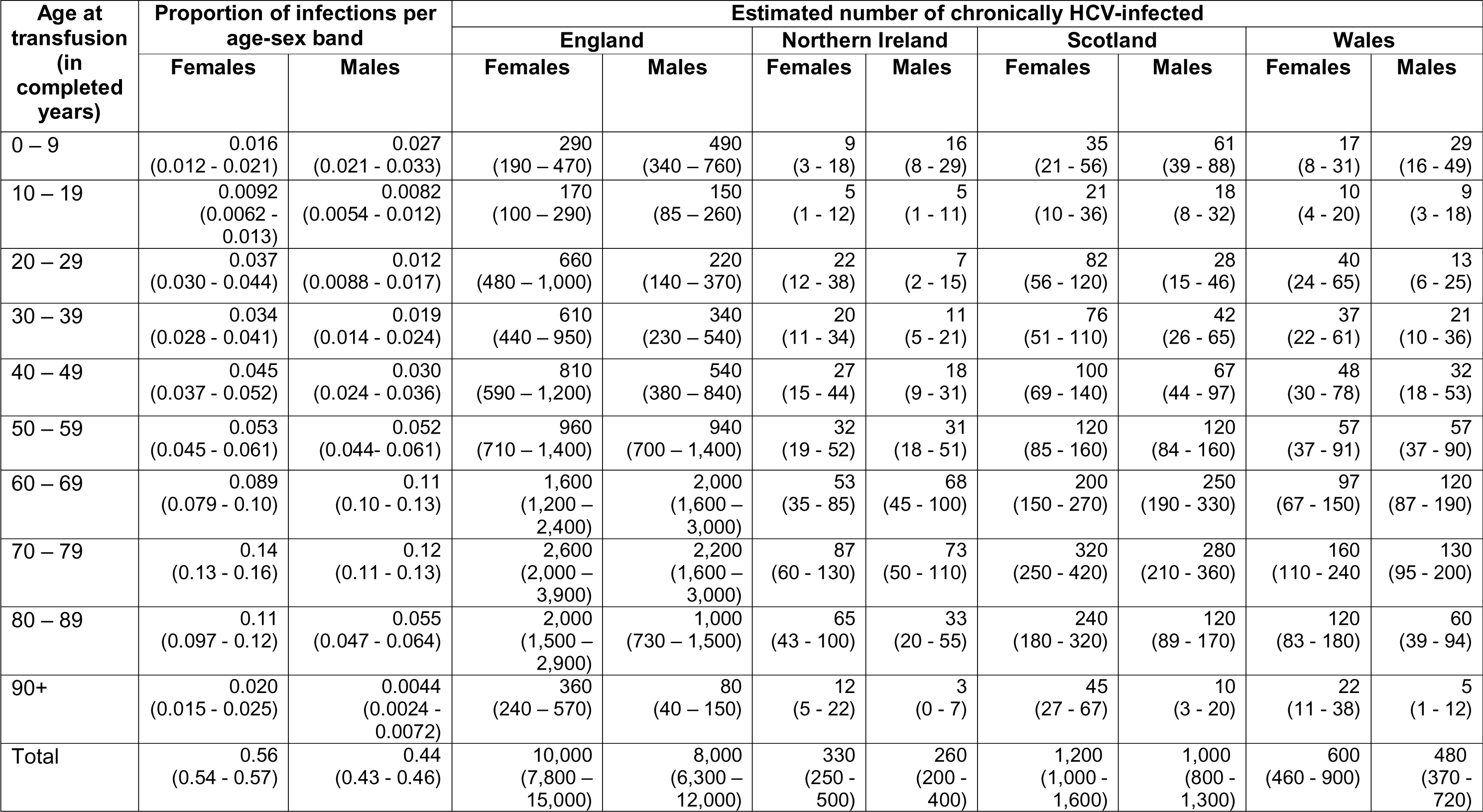
Estimated age-sex distribution of those chronically HCV-infected through transfusion in each of the four nations, January 1970 – August 1991. Estimates are provided as median and 95% uncertainty intervals.

**Table 6:** Estimated age-sex distribution of HCV-related deaths in each of the four nations, up until December 2019. Estimates are provided as median and 95% uncertainty intervals.

| Age at transfusion<br>(in completed years) | Proportion of infections per age-sex band |  | Estimated number HCV-related deaths |  |  |  |  |  |  |  |
| --- | --- | --- | --- | --- | --- | --- | --- | --- | --- | --- |
|  |  |  | England |  | Northern Ireland |  | Scotland |  | Wales |  |
|  | Females | Males | Females | Males | Females | Males | Females | Males | Females | Males |
| 0 – 9 | 0.016<br>(0.012 - 0.021) | 0.027<br>(0.021 - 0.033) | 0<br>(0 – 10) | 15<br>(0 – 35) | 0<br>(0 - 0) | 0<br>(0 - 0) | 0<br>(0 - 0) | 0<br>(0 - 4) | 0<br>(0 - 0) | 0<br>(0 - 0) |
| 10 – 19 | 0.0092<br>(0.0062 - 0.013) | 0.0082<br>(0.0054 - 0.012) | 0<br>(0 – 10) | 0<br>(0 – 10) | 0<br>(0 - 0) | 0<br>(0 - 0) | 0<br>(0 - 0) | 0<br>(0 - 0) | 0<br>(0 - 0) | 0<br>(0 - 0) |
| 20 – 29 | 0.037<br>(0.030 - 0.044) | 0.012<br>(0.0088 - 0.017) | 30<br>(10 – 70) | 10<br>(0 – 30) | 0<br>(0 - 0) | 0<br>(0 - 0) | 0<br>(0 - 10) | 0<br>(0 - 2) | 0<br>(0 - 3) | 0<br>(0 - 0) |
| 30 – 39 | 0.034<br>(0.028 - 0.041) | 0.019<br>(0.014 - 0.024) | 45<br>(15 – 100) | 35<br>(10 – 70) | 0<br>(0 - 3) | 0<br>(0 - 2) | 2<br>(0 - 14) | 1<br>(0 - 9) | 0<br>(0 - 6) | 0<br>(0 - 4) |
| 40 – 49 | 0.045<br>(0.037 - 0.052) | 0.030<br>(0.024 - 0.036) | 95<br>(35 – 190) | 75<br>(30 – 150) | 1<br>(0 - 7) | 1<br>(0 - 6) | 11<br>(0 - 23) | 8<br>(0 - 19) | 4<br>(0 - 13) | 3<br>(0 - 10) |
| 50 – 59 | 0.053<br>(0.045 - 0.061) | 0.052<br>(0.044 - 0.061) | 130<br>(50 – 250) | 120<br>(45 – 220) | 2<br>(0 - 9) | 2<br>(0 - 8) | 15<br>(3 - 29) | 14<br>(3 - 27) | 6<br>(0 - 16) | 6<br>(0 - 15) |
| 60 – 69 | 0.089<br>(0.079 - 0.10) | 0.11<br>(0.10 - 0.13) | 210<br>(85 – 380) | 250<br>(100 – 440) | 5<br>(0 - 13) | 6<br>(0 - 16) | 25<br>(9 - 44) | 30<br>(11 - 52) | 11<br>(2 - 24) | 14<br>(3 - 28) |
| 70 – 79 | 0.14<br>(0.13 - 0.16) | 0.12<br>(0.11 - 0.13) | 290<br>(120 – 510) | 140<br>(55 – 250) | 8<br>(1 - 18) | 2<br>(0 - 16) | 35<br>(13 - 60) | 16<br>(4 - 30) | 16<br>(4 - 32) | 6<br>(0 - 16) |
| 80 – 89 | 0.11<br>(0.097 - 0.12) | 0.055<br>(0.047 - 0.064) | 65<br>(25 – 130) | 10<br>(1 – 35) | 1<br>(0 - 4) | 0<br>(0 - 1) | 7<br>(0 - 16) | 0<br>(0 - 4) | 2<br>(0 - 8) | 0<br>(0 - 2) |
| 90+ | 0.020<br>(0.015 - 0.025) | 0.0044<br>(0.0024 - 0.0072) | 0<br>(0 – 10) | 0<br>(0 – 0) | 0<br>(0 - 0) | 0<br>(0 - 0) | 0<br>(0 - 1) | 0<br>(0 - 0) | 0<br>(0 - 0) | 0<br>(0 - 0) |
| Total | 0.56<br>(0.54 - 0.57) | 0.44<br>(0.43 - 0.46) | 880<br>(350 – 1,600) | 660<br>(250 – 1,200) | 18<br>(1 - 54) | 13<br>(1 - 42) | 71<br>(20 - 140) | 71<br>(20 - 180) | 41<br>(8 - 100) | 30<br>(4 - 73) |

### Deterministic sensitivity analysis

The results of the sensitivity analyses set out in Table 3 are presented in Supplementary Table 51.

We find that the major driver of our results is the assumption of the effect size of the stepdown in infectious donations in the mid-1980s (Scenarios A, D, E). For example, if the deferral effect size is halved to 33% with all else held constant, the estimated number of infections, and consequently the other quantities of interest, falls by 28% in England, Northern Ireland and Wales, and by 12% in Scotland (Scenario E). As the trend in prevalence was pegged to the estimated prevalence in September 1991, a smaller deferral effect would mean that the prevalence of HCV in donors prior to the mid-1980s (when the deferral effect took place) would be lower than if a larger deferral effect were observed, which drives this initially counterintuitive finding. A similar reduction was observed when following the IDU trend in past-IDUs rather than ever-IDUs in England, Northern Ireland and Wales, with a reduced deferral effect of 33% implemented one year later in 1987 (Scenario A). Estimates for Scotland were not modelled as no data were available for past-IDUs in Scotland and extrapolation from data for England (as was done for Wales and NI) was not appropriate in Scotland due to difference in the injecting drug use epidemic. In contrast, delaying the impact of the deferral year alone only resulted in an approximately 5% increase in estimated infections in all nations (Scenario D).

The constant proportion of the prevalence assumed to come from non-IDUs had a modest impact on the number of infections (Scenarios B, C, F), regardless of the proportion, in England, Northern Ireland and Wales. For example, when the non-IDU component is removed, infections increase by 3% (Scenario B). On the contrary, when the non-IDU component replaces the IDU component altogether, infections decrease by 9% (Scenario C). However, the number of IDUs in Scotland increases substantially faster than that modelled in England (Supplementary Figure 10). Consequently, this parameter plays a more important role in the model for Scotland, with infections falling by 18% when non-IDU component is excluded in the model and rising by 50% when driven by IDUs alone.

In comparison to age-stratified transfusion hazards, non-age-stratified hazards reduce the number of deaths by 1% in each of the nations, demonstrating that this assumption has a minimal impact on the results overall (Scenario H). Even without any additional transfusion hazards, deaths reduce by less than 2% compared to the baseline scenario (Scenario G). This is a similar reduction to that observed by considering age-stratified transfusion risks but without any additional hazards related to chronic HCV infection (Scenario I).

Bringing this together, a major driver of the model is the assumption around self-deferral, which is one of the quantities for which there was little direct evidence.

### Validation

Several funds have been set up to provide financial support to those infected or affected by bloodborne infections as a result of contaminated blood products. The Skipton Fund and the Caxton Fund were established for people infected with chronic HCV prior to September 1991 (36,37). After 2017, these funds were replaced by individual schemes for the four UK nations. Data from these funds can provide a useful calibration of our estimates.

Data from the Skipton Fund in 2016 report 4,165 approved applications in England. Of these, approximately 50% of the applications were by those with bleeding disorders and we thus assume that the approximately 2,080 other claimants were chronically HCV-infected from blood transfusions. The Skipton Fund was aware of the deaths of 619 of its UK claimants. We thus estimate that, *pro rata*, approximately 484 of these deaths would have occurred in England meaning that an estimated 1,596 people who were chronically HCV-infected by blood transfusion survived until 2016. Our estimate for the number of people in England that were chronically HCV-infected and survived to 2019 was 2,200. Whilst this is above the 1,596 surviving to 2016 estimated from the Skipton Fund, it is not substantially higher, especially when we consider that it is likely that many of those who were chronically HCV-infected by transfusion would not have registered a claim.

Information regarding surviving fund claimants who were chronically HCV-infected without HIV co-infection at 31 December 2014 was requested from the Department of Health and Social Care (DHSC). Within these DHSC data, exposure was classified as ‘haemophiliac’ or ‘other’. We assumed ‘haemophiliac’ refers to people with bleeding disorders and ‘other’ refers predominantly to those who acquired infection via blood transfusion. Within the category ‘other’, females outnumbered males by 1.5 to 1 (718/473; percentage females: 60.3% (95% CI 57.4% - 63.1%). We compared the sex distribution of surviving beneficiaries from information from funds to the sex distribution estimated by our model, finding that modelled estimates of female survivors outnumbered males by 1.77 to 1 (1,614/910; percentage females: 63.9% (95% CI (62.0% - 65.8%)) (Supplementary Table 52), which was a similar proportion to that which we observed in our model in 2019 (1,425/804; percentage females: 63.9% (95% CI (61.9% - 65.9%)).

## Discussion

This paper presents the statistical model developed by the SEG to estimate, for the IBI, the number of HCV infections, and subsequent deaths, transmitted from contaminated blood transfusions within the UK across January 1970 – August 1991. We have estimated that 26,800 people (95% uncertainty interval 21,300 – 38,800) throughout the UK were chronically infected with HCV because of contaminated blood transfusions in this period, with an estimated 1,820 (95% uncertainty interval 650 – 3,320) deaths having occurred to the end of 2019 as a result of HCV infection. These figures were presented as evidence to the IBI in 2022 (7).

Our model is adapted from that developed by Schnier and Goldberg (13), who answered a similar ascertainment question for the PI in Scotland in 2011. After careful consideration, we chose to work from Schnier and Goldberg as we believed it was the best way to estimate the numbers we wished to provide. In particular, this model’s clear and logical structure, which involved decomposing the complex process of HCV transmission into a series of six smaller “tasks”, each one representing a step in the transmission process. As is always the case, our model is an imperfect representation of reality and required a number of assumptions and data to parameterise it. A major issue that we encountered was the extensive amount of missing or poor-quality data across each of the six tasks. For example, estimates for the annual number of HCV-infected recipients are dependent on the annual number of blood transfusions and the HCV prevalence within the donor population during each year, but these data were not readily available for the entire time period being modelled. Consequently, most tasks within our model required further sub-modelling to fill these data gaps. The stochastic version of the model propagates uncertainty derived from imputing data through to the final results, which drives the relatively large uncertainty intervals presented.

We tested the impact of our model assumptions and parameterisation on the results through extensive sensitivity analyses, totalling over 750 distinct scenarios across the 4 nations of the UK. For brevity and clarity, we have only presented 9 principal scenarios in this paper which were determined to drive the key metrics of interest. We found that the step-down effect in HCV-infectious donations resulting from changing guidance in the mid-1980s was one of the biggest determinants of the model results. However, this was also the only part of the model for which no data at all were available, resulting in the SEG informally consulting members of other IBI Expert Groups who were knowledgeable about this change in guidance and its likely impact on blood donations. This is an important limitation of our model which should be considered when interpreting the results. Some assumptions were more difficult to test than others. For example, we followed Schnier and Goldberg in assuming that transfusion recipients only have a negligible chance of receiving two infected units of blood (assumption g, Table 1). However, this may not hold for rare blood groups, in which the chance of having two units from the same donor could be greater due to a more limited supply, but this would have a minimal effect on the overall results. Similarly, we assume, in the absence of any evidence to the contrary, that all infectious units cause an infection. Additionally, we did not account for the sensitivity and specificity of the assays used for detecting HCV antibodies that informed the prevalence estimates for the donor population in September 1991. This is particularly important when analysing differences between nations in which different assays may have been used in England and Wales than in Scotland. We have treated the four nations of the UK as independent epidemiological units. However, translocation of people between the nations will have occurred given the time scales involved, but we have no way of untangling such spatial effects. However, we expect that they will be relatively minor if net translocation is close to zero.

Given the public interest in and severity of this tragedy, alternative estimates of the number of individuals infected with HCV from transfusion in the UK nations have been produced by other groups. Schnier and Goldberg estimated 1,500 HCV infections in Scotland in 2012 (13), which is substantially lower than our corresponding central estimate of 2,740 HCV infections and is driven by our decision to implement a ‘hybrid’ model of HCV infection in the donor population (as detailed in Task 1) in comparison to Schnier and Goldberg’s assumption that HCV prevalence in the donor population was driven entirely by HCV- infectious ever-IDUs. In 2002, Soldan et al. (38) estimated approximately 23,500 HCV infections acquired from transfusions in England based on a number of similar assumptions to those implemented in our model. A key difference between the models was that Soldan et al. assumed the HCV prevalence in the donor population observed in September 1991 was constant from 1970, rather than being driven by the trend in ever-IDUs, and thus their estimate of 23,500 is slightly higher than our median estimate for England of 22,000. In 2011, the Department of Health extrapolated Soldan’s estimate to the entire UK on a population *pro rata* basis, resulting in an estimate of 28,000 HCV infections (39), which is similar to our central estimate of 26,800. Soldan et al. (40) also applied their model for England to Scotland for the PI under the higher observed HCV prevalence in the Scottish donor population, resulting in an estimate of almost 3,500 infections, which is substantially higher than our median estimate of 2,740 and Schnier and Goldberg’s estimate of 1,500. Although there have been estimates of up to 400,000 HCV infections, these appear to be largely based on unduly-elevated estimates of the prevalence of HCV, such as: in the UK donor population of at least 2% between 1970 and 1985 and 1% between 1986 and 1991 as presented by Ramsay et al. (41). However, there are concerns about the statistics used in this analysis. For example, the estimate of a 2.6% prevalence of HCV infection in transfusion recipients is based on a sample of HCV test results performed in the 1990s. These tests are likely to have been performed due to a suspicion of HCV infection (i.e., due to the presence of symptoms or being in a high-risk group), and thus are a biased sample and do not reflect the likely prevalence within all transfusion recipients.

Based on our experience of undertaking and presenting this work to the IBI, the SEG was explicitly asked for recommendations regarding future data collection and surveillance measures for bloodborne viruses. Twelve different recommendations were assessed by the authors of the SEG Report and full details of these can be found the Supplementary Report (42). Whilst some of these recommendations were specific for people with bleeding disorders, who are not the focus of this paper, others are more pertinent to transmission of infection via blood transfusion. Authors considered it a high priority that three surviving high-risk subgroups identified within the SEG Report who received transfusions prior to September 1991 should be advised to request an HCV test if they were unsure of their HCV- status as they may still benefit from diagnosis and treatment with directly acting antiviral therapy. Asking patients newly diagnosed with a blood-borne infection about their history of receipt or donation of blood or tissue could provide public health benefit if questions were framed appropriately. Limited data are currently available on the long-term outcomes of transfusion recipients within the UK. Establishing 5-yearly calendar cohorts of transfusion recipients to be followed up for mortality via record-linkage would improve monitoring of the use of transfusions and allow better assessment of the influence of factors such as demography, transfused units and underlying disease on patients’ survival. Long-term storage of representative residual samples from blood donations is recommended as it would allow the historical prevalence of future novel pathogens to be determined, whilst linkage of historical transfusions to a patient’s electronic health record would enable better risk-assessment by both patients and practitioners. In the event of a new transfusion-transmitted infection which may be transmitted sexually or from mother to baby, early testing of at-risk contacts and establishment of a research cohort to assess the impacts of these secondary infections is also recommended. It is possible that other novel blood-borne transmissible diseases will be identified in the future (43) and the recommendations outlined above would be relevant to the prompt identification and management of such diseases.

Infection via blood transfusion has inflicted enormous suffering upon many people (44). In amongst all of the technical detail provided in this paper, it is of the utmost importance to remember that every number we have presented here relates to a human life. While figures are unable to capture the suffering of individuals and their loved ones, it is only by collating the occurrence and sequelae of these incident infections as we have done that we can properly assess the magnitude of what has happened.

## Data Availability

All code is available at: https://github.com/ruthmccabe/IBI-SEG-HCV-model. Data which we are able to share are included throughout the manuscript and supplementary material.

## Funding

SH, RM, SMB and DJS gratefully acknowledge financial support from the IBI to undertake this work. This work was also supported by the NIHR Health Protection Research Unit (HPRU) in Emerging and Zoonotic Infections, a partnership between UKHSA, University of Oxford, University of Liverpool and Liverpool School of Tropical Medicine [grant number NIHR200907 supporting SH, RM and CAD]; and the MRC Centre for Global Infectious Disease Analysis [grant number MR/R015600/1 supporting CAD], which is jointly funded by the UK Medical Research Council (MRC) and the UK Foreign, Commonwealth and Development Office (FCDO), under the MRC/FCDO Concordat agreement and is also part of the EDCTP2 programme supported by the European Union (EU).

### Disclaimer

“The views expressed are those of the authors and not necessarily those of the Infected Blood Inquiry, United Kingdom (UK) Department of Health and Social Care, EU, FCDO, MRC, National Health Service, NIHR, or PHE. The funding bodies had no role in the design of the study, analysis and interpretation of data and in writing the manuscript.”

## Acknowledgements

The work undertaken by SEG for the IBI was prepared rapidly and under tight deadlines, and would not have been possible without the generous assistance of numerous individuals and organisations. We would particularly like to highlight:

- The Infected Blood Inquiry team – including Moore Flannery, Tom Dunning, Annabel Sharma, Connor Mitton, Michelle Secker, and Elli Brooks – for their extensive advice and support in producing this report, particularly in commissioning three types of linkage study (as advised by Sheila Bird), helping access data from numerous agencies, and in detecting a mass of relevant documents in Relativity.
- The team at the Scottish National Blood Transfusion Service and Edinburgh University for conducting the record-linkage follow-up to 31 December 2019 of four cohorts of transfusion recipients in 1999; 2004; 2009 and 2014.

◦ Professor Marc Turner, Director, Scottish National Blood Transfusion Service
◦ Dr Katherine J Forrester, Transfusion Researcher, Scottish National Blood Transfusion Service
◦ Amanda Stewart, Head of Analytics and Planning, Scottish National Blood Transfusion Service
◦ Dr Vanda Inacio, Lecturer in Statistics, School of Mathematics, Edinburgh University
- Colleagues on other Infected Blood Inquiry expert groups who advised on the possible effect of donor guidance in the mid-1980s.
- The team at UKHSA for unpublished annual data on estimated number [1971-1991] of chronically HCV infected a) ever-IDUs; b) past-IDUs; c) current-IDUs

◦ Dr Ross Harris, Senior Statistician, Statistics Modelling and Economics Department, UK Health Security Agency
◦ Professor Daniela De Angelis, Deputy Director, MRC Biostatistics Unit, Cambridge University
- The team at UKHSA for updated analysis of follow-up on their HCV look-back case control cohort study.

◦ Dr Ross Harris, Senior Statistician, Statistics Modelling and Economics Department, UK Health Security Agency
◦ Dr. Helen Harris, Clinical Scientist – Epidemiology, UKHSA
◦ Annastella Costella, Hepatitis Scientist, UKHSA
◦ Dr. Sema Mandal, Medical Consultant Epidemiologist, UKHSA
◦ Dr. Monica Desai, Medical Consultant Epidemiologist, UKHSA
◦ Dr. Mary Ramsay, Head of Immunisation Department, UKHSA also the HCV National Register Steering Group.
- The team from Wallis et al. (2014) for providing their original data.
- The team at NHSBT for providing detailed survival analyses of EASTR data.
- The Department of Health and Social Care/EIBSS analytical team who provided demographic and survival information about EIBSS claimants.

◦ Andrew Parker, Principal Operational Research Analyst, Department of Health and Social Care (DHSC)
◦ Joshua Walden, Senior Economic Adviser, DHSC
◦ Omar Idriss, Deputy Director, DHSC.

## Author contributions

SH and RM are joint first authors and contributed equally to the work in this manuscript. SH and RM were responsible for the analysis and preparation of this manuscript, under close supervision from DJS and SMB and with input from DDA, CAD, SJWE and GFM. All coauthors contributed to redrafting.

## Notes

### Competing Interest Statement

The authors have declared no competing interest.

