## Supplementary Material for "Modelling hepatitis C infection acquired from blood transfusions in the UK between 1970 and 1991 for the Infected Blood Inquiry"

### Supplementary Information

Sarah Hayes[1,2,*], Ruth McCabe[1,2,*], Daniela De Angelis[3], Christl A Donnelly[1,2,4,5], Stephen JW Evans[6], Graham F Medley[6], David J Spiegelhalter[7,+], Sheila M Bird[3,8,+]

[1] Department of Statistics, University of Oxford, Oxford, UK

[2] NIHR Health Protection Research Unit in Emerging and Zoonotic Infections, Liverpool, UK

[3] MRC Biostatistics Unit, University of Cambridge, School of Clinical Medicine, Cambridge Institute of Public Health, Cambridge, UK

[4] Pandemic Sciences Institute, University of Oxford, Oxford, UK

[5] MRC Centre for Global Infectious Disease Analysis, Imperial College London, London, UK

[6] London School of Hygiene & Tropical Medicine, London, UK

[7] Centre for Mathematical Sciences, University of Cambridge, Cambridge, UK

[8] Edinburgh University's College of Medicine and Veterinary Medicine, Edinburgh, UK

[+] Joint senior authors

### Task 1: What proportion of blood donations received between January 1970 and August 1991 were HCV-infectious?

Supplementary Table 1 presents the overall results of Task 1 for England.

*Supplementary Table 1: Results from our baseline model for England, showing steps in obtaining estimates of the percentage of HCV-infectious donations each year. Abbreviations: Hepatitis C virus (HCV); Injecting drug user (IDU).*

| Year | (a) Estimated number of chronically HCV-infected ever-IDUs in England | (b) Estimated ratio of chronically HCV-infected ever-IDUs in England, relative to number in 1991 (set to 100) | (c) Estimated percentage of HCV-infectious donations (assuming ever-IDUs are sole source), pegged to estimated HCV-infectious donations as in September 1991 | (d) Estimated percentage of HCV-infectious donations assuming ever-IDUs are sole source, and assuming 'step-down' in 1985) | (e) Estimated percentage of HCV-infectious donations, assuming a constant % from non-IDUs, fixed at 25% of observed 1991 rates |
|---|---|---|---|---|---|
| 1970 | 21,000 (18,000 - 26,000) | 20 (16 - 25) | 0.010% | 0.029% | 0.034% |
| 1971 | 23,000 (20,000 - 27,000) | 22 (18 - 26) | 0.011% | 0.032% | 0.036% |
| 1972 | 26,000 (22,000 - 29,00) | 24 (20 - 28) | 0.012% | 0.035% | 0.039% |
| 1973 | 28,000, (24,000 - 32,000) | 26 (22 - 31) | 0.013% | 0.039% | 0.041% |
| 1974 | 31,000 (26,000 - 34,000) | 29 (24 - 34) | 0.014% | 0.042% | 0.044% |
| 1975 | 33,000 (29,000 - 38,000) | 32 (27 - 38) | 0.015% | 0.046% | 0.047% |
| 1976 | 37,000 (32,000 - 41,000) | 35 (29 - 41) | 0.017% | 0.051% | 0.050% |
| 1977 | 40,000 (35,000 - 45,000) | 38 (32 - 45) | 0.018% | 0.056% | 0.054% |
| 1978 | 45,000 (39,000 - 50,000) | 42 (36 - 49) | 0.020% | 0.061% | 0.058% |
| 1979 | 49,000 | 46 | 0.022% | 0.067% | 0.062% |

| | | | | | |
|---|---|---|---|---|---|
| | (43,000 - 55,000) | (39 - 54) | | | |
| 1980 | 52,000<br>(47,000 - 58,000) | 49<br>(42 - 58) | 0.024% | 0.073% | 0.067% |
| 1981 | 56,000<br>(50,000 - 62,000) | 53<br>(45 - 61) | 0.026% | 0.079% | 0.072% |
| 1982 | 60,000<br>(54,000 - 66,000) | 56<br>(49 - 66) | 0.028% | 0.086% | 0.077% |
| 1983 | 64,000<br>(57,00 - 72,000) | 60<br>(52 - 71) | 0.031% | 0.093% | 0.082% |
| 1984 | 69,000<br>(60,000 - 79,000) | 65<br>(55 - 77) | 0.033% | 0.100% | 0.088% |
| 1985 | 74,000<br>(65,000 - 84,000) | 70<br>(60 - 83) | 0.036% | 0.036% | 0.039% |
| 1986 | 80,00<br>(71,000 - 90,000) | 76<br>(64 - 89) | 0.038% | 0.038% | 0.041% |
| 1987 | 87,000<br>(76,000 - 97,000) | 829<br>(70 - 96) | 0.040% | 0.040% | 0.043% |
| 1988 | 93,000<br>(81,000 - 100,000) | 88<br>(74 - 100) | 0.043% | 0.043% | 0.044% |
| 1989 | 100,000<br>(87,000 - 110,000) | 94<br>(80 - 110) | 0.045% | 0.045% | 0.046% |
| 1990 | 100,000<br>(90,000 - 112,000) | 97<br>(82 - 110) | 0.047% | 0.047% | 0.047% |
| 1991 | 110,000<br>(94,000 - 120,000) | 100 | 0.049% | 0.049% | 0.049% |

*Task 1.3: How many chronically HCV-infected ever-injecting drug users (ever-IDUs) were alive during each year between 1970 and 1991?*

We were provided with estimates of the number of past, current and ever IDUs in England across 1971 – 1991. As the scope of the model encompasses 1970, we estimated a value for 1970 by:

$$I_{1970} = I_{1971} \times \left(\frac{I_{1971}}{I_{1975}}\right)^{\frac{1}{4}}.$$

#### *Task 1.4: What was the step-down in donations from ever-IDUs due to self-deferral guidance and HIV antibody testing in the mid-1980s?*

A key uncertainty in forward projection of the numbers of people infected with HCV in England by blood transfusion during 1970-1991 is how effective self-deferral information-leaflets for blood donors in the HIV/AIDS era, together with the advent of HIV antibody testing of the blood supply, were in dissuading donations from past and current IDUs who (unknowingly) were HCV-infected.

At short notice, we sought expert opinion from selected members who serve on other Expert Groups for the Infected Blood Inquiry and were in professional practice in the 1980s. This was relatively successful: only one non-response (on account of annual leave) out of eight, and six provided some quantification.

Six assessments are summarised in Supplementary Table 2 in terms of the % of HCV-infected ever-IDUs who were dissuaded from donation by self-deferral leaflets, HIV antibody testing or other awareness about the risk of blood-borne infection. Our informants had been given the following instructions: *"Please place 20 betting tokens in the betting sheets below to express your opinion about the likely percentage-reduction in HCV-infected donations from early in 1985 to 1986.".*

The resultant distribution is bimodal. Ignoring outlying votes which sum to fewer than 3 (n=9), the lower mode has accrued 36/110 votes. These 36 votes support a mean % reduction of 31% (1130/36; most likely 30%). The upper mode accrued 74/110 votes. These 74 votes support a mean % reduction of 67% (4960/74; most likely 70%). Consistent with the elicited bimodal distribution, the baseline scenario in our simulation model is a 67% reduction, with a sensitivity analysis of a one-third reduction for England; as well as no reduction to mimic the English HCV lookback model assumptions.

In addition, a seventh assessor judged that, on account of venous access, the number of current-IDUs who would have attended as blood donors would be very low – even before HIV antibody testing was introduced. There would have been a further 50% reduction (with 50% confidence).

Other contexts that respondents mentioned included:

- greater impact on HCV-infected current-IDUs;
- potentially greater impact from HIV antibody testing than from self-deferral leaflets;
- literature on limited impact on public health behaviours via leaflets;
- reluctance of blood donors to be turned down and hence minimization or oversight of historical risk-factors;
- attendance as blood donor specifically to access HIV antibody testing;
- whether potential donors appreciated that past history of having been ever-IDU mattered (vs being a current-IDU);
- sensitivity of the HIV antibody test used for initial screening of blood supply;
- how risks to blood supply from non-A, non-B hepatitis infections in ever-IDUs were appreciated in contemporary medical reports, let alone by potential donors;
- in the early 1990s, the proportion of blood donors who, having tested HCV antibody positive, admitted a history of injecting or sexual risk being less than 40%;
- much health education literature requires too high a level of literacy and that written in the 1980s often appeared hectoring.

*Supplementary Table 2: Elicitation on % reduction in HCV-infected donations by ever-IDUs across England in 1986 (versus 1985 as a baseline) that was achieved by self-deferral advice and HIV antibody testing. ** Seventh assessor judged that, on account of venous access, the number of current-IDUs who would have attended as blood donors would be very low – even before HIV antibody testing was introduced. There would have been a further 50% (with 50% confidence).*

| | **Six assessments (A), please see footnote** | | | | | | | | | | | | | | | | | | | **Total** |
|---|---|---|---|---|---|---|---|---|---|---|---|---|---|---|---|---|---|---|---|---|
| | | | | | | | | | | | | 1 | 4 | 10 | 4 | 1 | | | | 20 |
| | 1 | 1 | 1 | 1 | 1 | 1 | 1 | 1 | 1 | 1 | 1 | 1 | 1 | 1 | 1 | 1 | 1 | 1 | 1 | 19 |
| **Betting** | | | | 6 | | 10 | | 4 | | | | | | | | | | | | 20 |
| **Counters** | 1 | 1 | 1 | 1 | 1 | 1 | 2 | 3 | 2 | 1 | 1 | 1 | 1 | 1 | 1 | 1 | | | | 20 |
| | | | | | | | | | | 4 | 4 | 4 | 4 | 4 | | | | | | 20 |
| | | | | | | | | | | | | | 4 | 6 | 6 | 4 | | | | 20 |
| | | | | | | | | | | | | | | | | | | | | ** |
| **TOTALS** | **2** | **2** | **2** | **8** | **2** | **12** | **3** | **8** | **3** | **6** | **6** | **7** | **14** | **22** | **12** | **7** | **1** | **1** | **1** | **119** |
| **%** reduction | 5% | 10% | 15% | 20% | 25% | 30% | 35% | 40% | 45% | 50% | 55% | 60% | 65% | 70% | 75% | 80% | 85% | 90% | 95% | |
| From baseline 100 to | 95 | 90 | 85 | 80 | 75 | 70 | 65 | 60 | 55 | 50 | 55 | 40 | 35 | 30 | 25 | 20 | 15 | 10 | 5 | |

Six respondents offered an opinion on the % of HCV-infected past-IDUs who were dissuaded from donation by self-deferral leaflets, HIV antibody testing or other awareness about the risk of blood-borne infection. See Supplementary Table 3.

The resultant distribution is again bimodal. Ignoring outlying votes, the lower mode has accrued 48/113 votes. These 48 votes support a mean % reduction of 24% (1160/48; most likely 20%). The upper mode accrued 65/113 votes. These 65 votes support a mean % reduction of 67% (4330/65; most likely 65%). Consistent with the elicited bimodal distribution, sensitivity analysis for the simulation model has used 67% and 33% reductions for England's chronically infected past-IDUs.

*Supplementary Table 3: Elicitation on % reduction in HCV-infected donations by past-IDUs across England in 1986 (versus 1985 as a baseline) that was achieved by self-deferral advice and HIV antibody testing. Until 1987, leaflets asked drug addicts or drug abusers to refrain from donating: they did not specifically encompass past users. Someone who used intravenous drugs occasionally, decades previously might not consider themselves a "drug abuser". After 1987, leaflets are specific that this applies to an ever-user in the previous decade. Hence, 70 to 80% decrease amongst past users in response (certainty level: 70-80%). **Seventh assessor judged that, on account of venous access, the number of current-IDUs who would have attended as blood donors would be very low – even before HIV antibody testing was introduced. There would have been a further 50% (with 50% confidence).*

| | **Six assessments (A)** | | | | | | | | | | | | | | | | | | | **Total** |
|---|---|---|---|---|---|---|---|---|---|---|---|---|---|---|---|---|---|---|---|---|
| | | | | | | | | | | | 1 | 4 | 10 | 4 | 1 | | | | | 20 |
| **Betting** | 1 | 1 | 1 | 1 | 1 | 1 | 1 | 1 | 1 | 1 | 1 | 1 | 1 | 1 | 1 | 1 | 1 | 1 | 1 | **19** |
| | | 6 | | 10 | | 4 | | | | | | | | | | | | | | 20 |
| | | | | | | | | | | | | | | | | | | | | ** |
| | | | | 4 | 4 | 4 | 4 | 4 | | | | | | | | | | | | 20 |
| | | | | | | | | | | | 4 | 6 | 6 | 4 | | | | | | 20 |
| | | | | | | | | | | | | 2 | 2 | 5 | 5 | 5 | 1 | | | 20 |
| **Counters TOTALS** | 1 | 7 | 1 | 15 | 5 | 9 | 5 | 5 | 1 | 1 | 6 | 13 | 19 | 14 | 7 | 6 | 2 | 1 | 1 | **119** |
| **% reduction** | 5 % | 10 % | 15 % | 20 % | 25 % | 30 % | 35 % | 40 % | 45 % | 50 % | 55 % | 60 % | 65 % | 70 % | 75 % | 80 % | 85 % | 90 % | 95 % | |
| From baseline 100 to | 95 | 90 | 85 | 80 | 75 | 70 | 65 | 60 | 55 | 50 | 55 | 40 | 35 | 30 | 25 | 20 | 15 | 10 | 5 | |

### Task 2: How many blood components were transfused?

Supplementary Table 4 presents the stages in estimating the number of units transfused, with data from England taken for illustration purposes.

*Supplementary Table 4: Stages in estimating the number of units transfused in England, 1970 – August 1991 – results reported to 3 significant figures. The estimates displayed in columns 2, 3 and 5 for 1991 are for January to August 1991 and not the full 12-month year. *Indicates estimates from extrapolation and interpolation from data from 1975 – 1976 and 1978 – 1979. **Indicates estimates from extrapolation from data from 1978 – 1990. *** Indicates estimated by extrapolating from known data for 1982 – 1988.*

| Year | Recorded donations in England and Wales | Estimated number of donations in England | Estimated number of units transfused per donation | Estimated number of units transfused in England |
|---|---|---|---|---|
| 1970 | 1,460,000* | 1,380,000 | 0.80*** | 1,100,000 |
| 1971 | 1,530,000* | 1,440,000 | 0.82*** | 1,180,000 |
| 1972 | 1,600,000* | 1,510,000 | 0.83*** | 1,250,000 |
| 1973 | 1,670,000* | 1,580,000 | 0.85*** | 1,340,000 |
| 1974 | 1,750,000* | 1,650,000 | 0.86*** | 1,420,000 |
| 1975 | 1,780,000 | 1,680,000 | 0.88*** | 1,480,000 |
| 1976 | 1,960,000 | 1,850,000 | 0.89*** | 1,660,000 |
| 1977 | 2,000,000* | 1,880,000 | 0.91*** | 1,720,000 |
| 1978 | 2,120,000 | 2,000,000 | 0.93*** | 1,860,000 |
| 1979 | 2,140,000 | 2,020,000 | 0.94*** | 1,910,000 |
| 1980 | 2,220,000 | 2,090,000 | 0.96*** | 2,010,000 |
| 1981 | 2,070,000 | 1,950,000 | 0.97*** | 1,900,000 |
| 1982 | 2,060,000 | 1,940,000 | 0.99 | 1,930,000 |
| 1983 | 2,140,000 | 2,020,000 | 1.03 | 2,070,000 |
| 1984 | 2,160,000 | 2,040,000 | 1.07 | 2,170,000 |
| 1985 | 2,120,000 | 2,000,000 | 1.10 | 2,210,000 |
| 1986 | 2,130,000 | 2,010,000 | 1.14 | 2,290,000 |
| 1987 | 2,090,000 | 1,980,000 | 1.18 | 2,330,000 |
| 1988 | 2,140,000 | 2,020,000 | 1.22 | 2,460,000 |
| 1989 | 2,230,000 | 2,100,000 | 1.23*** | 2,590,000 |
| 1990 | 2,180,000 | 2,060,000 | 1.25*** | 2,570,000 |
| 1991 | 1,440,000** | 1,360,000 | 1.26*** | 1,720,000 |

#### *Task 2.1 How many blood donations were there annually between January 1970 and August 1991?*

Data on the number of donations per year in England and Wales were observed for 1975 – 1976 and 1978 – 1990 (1,2). Estimates for the missing years were modelled using two Poisson regression models to account for the distinct difference in the number of observations from 1978 compared to that observed in 1975 – 1976 (Supplementary Figure 1). In each case, the model is described as:

$$\log(\widehat{Number\ of\ donations}) = \hat{\beta}_0 + \hat{\beta}_1 Year$$

Specifically, we first produced a model fitted to the observed data from 1975 – 1976 and 1978 – 1979 and used this to estimate values for 1970 – 1974 and 1977 (Supplementary Figure 1A). We then fitted a model to data from 1978 – 1990 and used this to estimate the number of donations in 1991 (Supplementary Figure 1B). As our study period goes to August 1991, we then scale the estimated value for 1991 by 0.67 (8 months of the year). The prediction intervals plotted in Supplementary Figure 1 reflect a compromise between the optimistic assumption of Poisson variation and the observed overdispersion in the data available.

As noted in the main text, the number of donations for England and Wales were taken from scaling these values in accordance with the population size of each country. Across 1970 – 1991, on average England accounted for 94.3% of the population of England and Wales, and thus 94.3% of donations were attributed to England, with the remaining 5.7% attributed to Wales (3).

As we were unable to obtain specific information on the number of donations in Northern Ireland during this period, analogous to the case of England and Wales, we assumed the number of donations could be approximated by assuming these values were proportional to Northern Ireland's population in comparison to England. Across 1970 – 1991, on average Northern Ireland's population was 3.3% that of England's and so the number of donations in Northern Ireland was assumed to be 3.3% the number of those observed in England (3).

The estimates for England and Wales are presented in Supplementary Figure 2, with the estimates for all four nations presented in Supplementary Table 5.

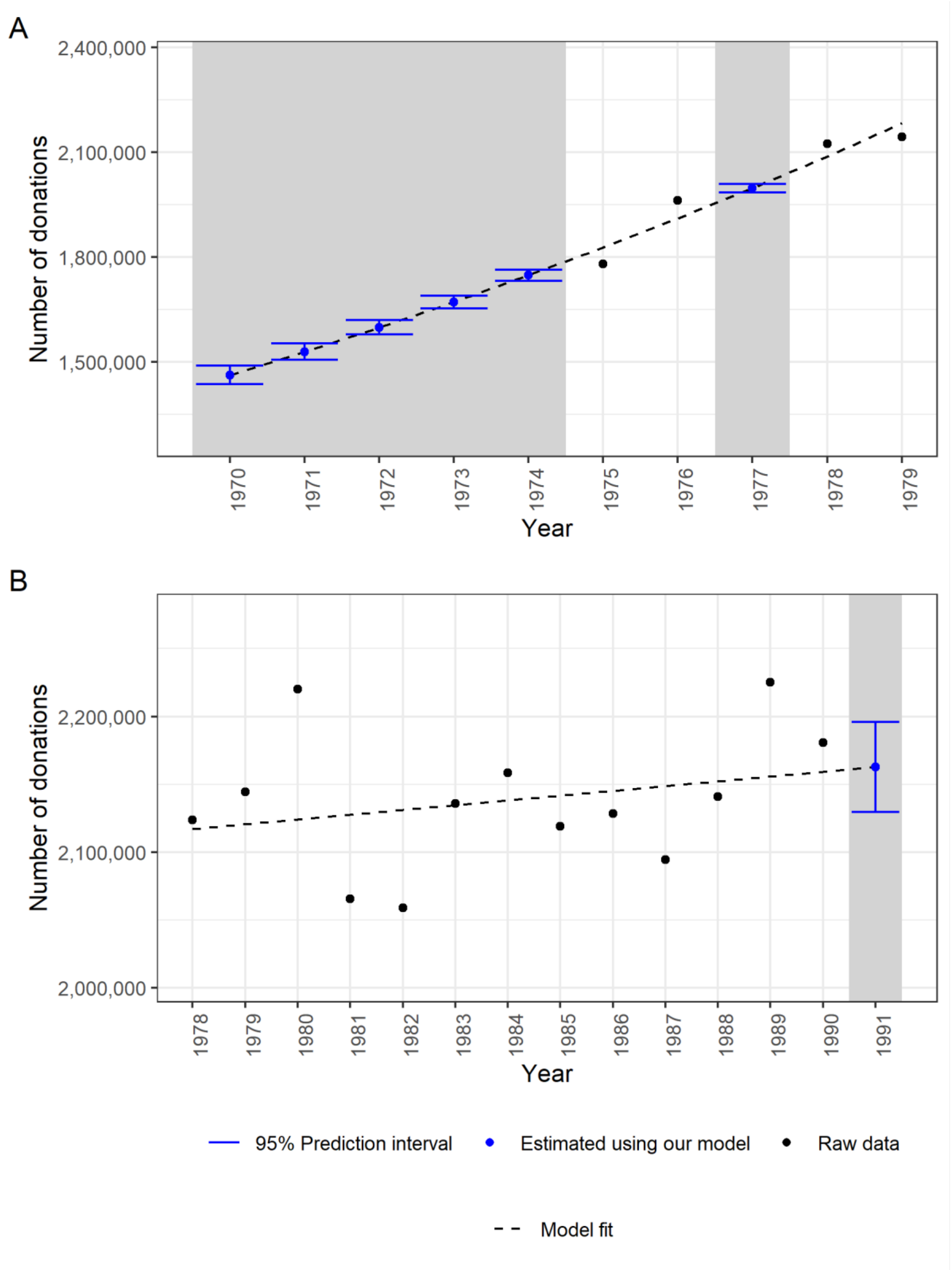

*Supplementary Figure 1: Estimating the number of donations in England and Wales in years without data using Poisson regression. (A) Missing years are interpolated using a model fit to 1975 – 1976 and 1978 – 1979. (B) Missing year is interpolated using a model fit to 1978 – 1990.*

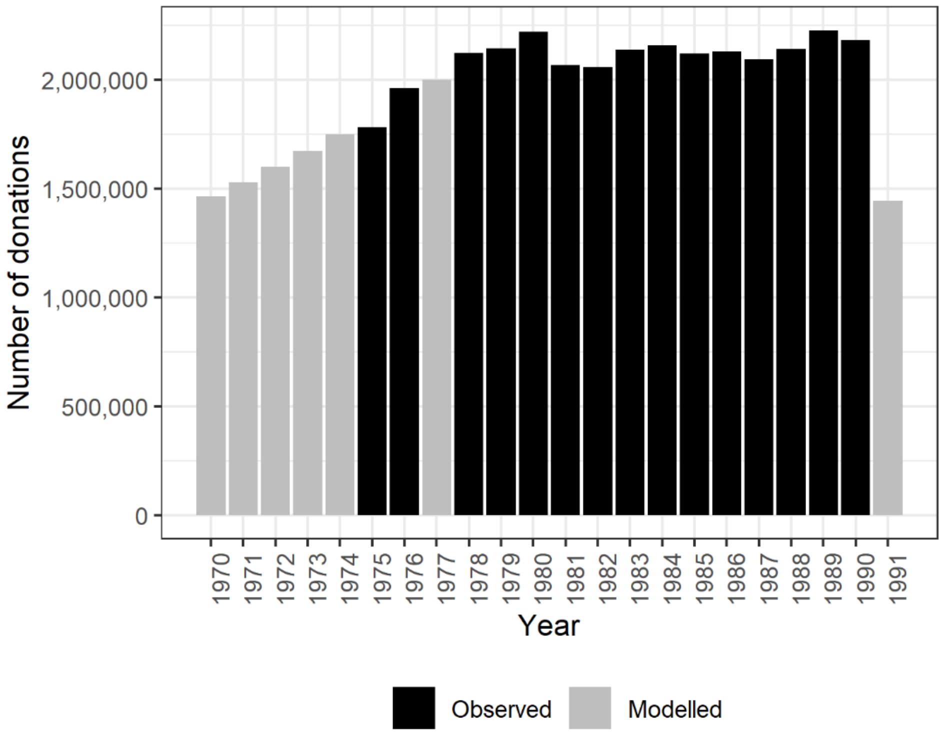

*Supplementary Figure 2: Number of blood donations in England and Wales, January 1970 – August 1991. The estimate displayed for 1991 is substantially lower than the estimate obtained for 1991 in Supplementary Figure 1 because the former is for January to August 1991 and the latter is for the full 12-month year.*

*Supplementary Table 5: The number of blood donations in each nation of the UK across 1970 – August 1991. The estimates displayed for 1991 are for January to August 1991 and not the full 12-month year. *Indicates estimates from extrapolation from data from 1975 – 1976 and 1978 – 1979. **Indicates estimates from extrapolation from data from 1978 – 1990.*

| Year | Recorded donations in England and Wales | Estimated number of donations in England | Estimated number of donations in Wales | Estimated number of donations in Northern Ireland | Estimated number of donations in Scotland |
|---|---|---|---|---|---|
| 1970 | 1,462,577* | 1,380,032 | 82,545 | 45,584 | 244,463 |
| 1971 | 1,529,182* | 1,442,878 | 86,304 | 47,660 | 248,216 |
| 1972 | 1,598,820* | 1,506,586 | 90,234 | 49,831 | 252,026 |
| 1973 | 1,671,629* | 1,577,286 | 94,343 | 52,100 | 255,895 |
| 1974 | 1,747,754* | 1,649,114 | 98,640 | 54,472 | 259,823 |
| 1975 | 1,780,000 | 1,679,540 | 100,460 | 55,477 | 248,558 |
| 1976 | 1,962,000 | 1,851,269 | 110,731 | 61,150 | 262,549 |
| 1977 | 1,997,568* | 1,884,829 | 112,739 | 62,258 | 277,772 |
| 1978 | 2,123,607 | 2,003,755 | 119,852 | 66,187 | 283,306 |
| 1979 | 2,144,484 | 2,023,454 | 121,030 | 66,837 | 290,078 |
| 1980 | 2,220,036 | 2,094,742 | 125,294 | 69,192 | 289,324 |
| 1981 | 2,065,428 | 1,948,859 | 116,569 | 64,373 | 293,501 |
| 1982 | 2,058,994 | 1,942,788 | 116,206 | 64,173 | 297,851 |
| 1983 | 2,135,840 | 2,015,297 | 120,543 | 66,568 | 302,233 |
| 1984 | 2,158,626 | 2,036,797 | 121,829 | 67,278 | 308,617 |
| 1985 | 2,119,060 | 1,999,464 | 119,596 | 66,045 | 304,914 |
| 1986 | 2,128,450 | 2,008,325 | 120,125 | 66,338 | 309,748 |
| 1987 | 2,094,316 | 1,976,117 | 118,199 | 65,274 | 289,006 |
| 1988 | 2,140,810 | 2,019,987 | 120,823 | 66,723 | 310,785 |
| 1989 | 2,225,009 | 2,099,434 | 125,575 | 69,347 | 321,588 |
| 1990 | 2,180,858 | 2,057,775 | 123,083 | 67,971 | 331,979 |
| 1991 | 1,441,838** | 1,360,464 | 81,374 | 44,938 | 238,979 |

*Task 2.2: Per donation, on average how many units were transfused?*

Data on the number of units transfused per donation were obtained annually for 1982 - 1988 from the National Blood Transfusion Service Statistics (NBTSS) for England and Wales (Supplementary Table 6) (4).

We sought to extrapolate the trend observed during this period to 1970 - 1981 and to 1989 - 1991 using a linear model as follows:

$$Units\ per\ \widehat{donation} = \widehat{\beta_0} + \widehat{\beta_1}Year$$

However, there is a steep increase observed in the number of units transfused per donation between 1982 - 1988 which is unlikely to hold throughout the entire period 1970 - 1991. Therefore, we scaled the gradient down to 25% of its estimated values for the periods without data and took the values expected under this model (Supplementary Figure 3).

The resulting number of units transfused per year in each nation is shown in Supplementary Table 7.

*Supplementary Table 6: Data from the National Blood Transfusion Service Statistics (NBTSS) (4) from which the number of units transfused per donation was approximated. RBC: red blood cells.*

| Year | (a) Number of donations | (b) Units issued (whole +RBC) | (c) Units returned unused | (d) % returned (c/b) | (e) Cryoprecipitate | (f) Donations from which plasma retained | (g) platelets | (h) total units available for use (b+e+f+g) | (i) Available units per donation (h/a) | (j) not used (assuming % returned) (d x h) | (k) usage per unit (1 - j/h) | (l) total units used (h - j) | (m) units used per donation (l/a) |
|---|---|---|---|---|---|---|---|---|---|---|---|---|---|
| 1978 | 2124 | 1663 | 239 | 14% | 164 | | | | | | | | |
| 1979 | 2145 | 1705 | 226 | 13% | 145 | | | | | | | | |
| 1980 | 2220 | 1793 | 234 | 13% | 127 | | | | | | | | |
| 1981 | 2065 | 1837 | 234 | 13% | 103 | | | | | | | | |
| 1982 | 2059 | 1785 | 197 | 11% | 89 | 98 | 329 | 2301 | 1.12 | 254 | 0.89 | 2047 | 0.99 |
| 1983 | 2136 | 1830 | 172 | 9% | 94 | 122 | 364 | 2410 | 1.13 | 227 | 0.91 | 2183 | 1.02 |
| 1984 | 2159 | 1873 | 149 | 8% | 96 | 135 | 404 | 2508 | 1.16 | 200 | 0.92 | 2308 | 1.07 |
| 1985 | 2119 | 1844 | 143 | 8% | 81 | 157 | 462 | 2544 | 1.20 | 197 | 0.92 | 2347 | 1.11 |
| 1986 | 2128 | 1861 | 144 | 8% | 73 | 172 | 547 | 2653 | 1.25 | 205 | 0.92 | 2448 | 1.15 |
| 1987 | 2094 | 1833 | 148 | 8% | 75 | 188 | 613 | 2709 | 1.29 | 219 | 0.92 | 2490 | 1.19 |
| 1988 | 2141 | 1867 | 128 | 7% | 63 | 208 | 631 | 2769 | 1.29 | 190 | 0.93 | 2579 | 1.20 |

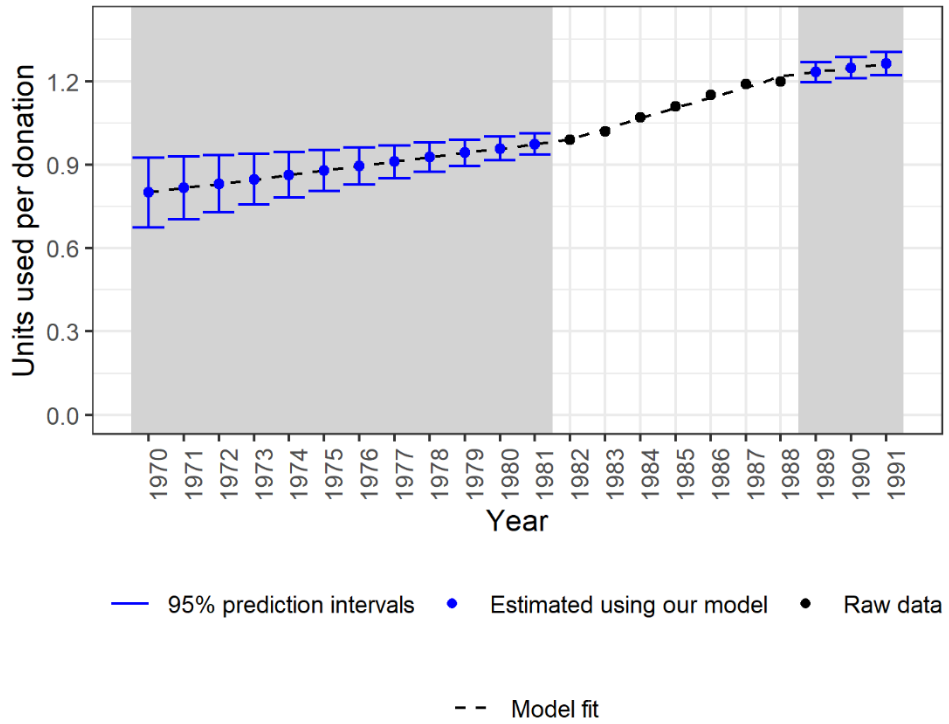

*Supplementary Figure 3: Extrapolation of units per donation from observed data in 1982 to 1988.*

*Supplementary Table 7: The estimated number of units transfused in each nation. The estimated numbers of units transfused displayed for 1991 are for January to August 1991 and not the full 12-month year. *** Indicates estimates from extrapolation.*

| Year | Estimated proportion of units transfused per donation | Estimated number of units transfused in England | Estimated number of units transfused in Northern Ireland | Estimated number of units transfused in Scotland | Estimated number of units transfused in Wales |
|---|---|---|---|---|---|
| 1970 | 0.80*** | 1,104,023 | 36,467 | 195,570 | 66,036 |
| 1971 | 0.82*** | 1,177,145 | 38,883 | 202,502 | 70,409 |
| 1972 | 0.83*** | 1,254,637 | 41,443 | 209,601 | 75,044 |
| 1973 | 0.85*** | 1,336,746 | 44,155 | 216,870 | 79,955 |
| 1974 | 0.86*** | 1,423,732 | 47,027 | 224,313 | 85,159 |
| 1975 | 0.88*** | 1,476,592 | 48,773 | 218,523 | 88,321 |
| 1976 | 0.89*** | 1,656,882 | 54,729 | 234,981 | 99,104 |
| 1977 | 0.91*** | 1,716,761 | 56,707 | 253,003 | 102,686 |
| 1978 | 0.93*** | 1,856,809 | 61,333 | 262,530 | 111,063 |
| 1979 | 0.94*** | 1,907,101 | 62,994 | 273,398 | 114,071 |
| 1980 | 0.96*** | 2,007,457 | 66,309 | 277,268 | 120,073 |
| 1981 | 0.97*** | 1,898,509 | 62,710 | 285,918 | 113,557 |
| 1982 | 0.99 | 1,926,829 | 63,646 | 295,404 | 115,251 |
| 1983 | 1.03 | 2,074,316 | 68,517 | 311,084 | 124,073 |
| 1984 | 1.07 | 2,172,826 | 71,771 | 329,228 | 129,965 |
| 1985 | 1.10 | 2,207,980 | 72,933 | 336,712 | 132,068 |
| 1986 | 1.14 | 2,293,077 | 75,744 | 353,666 | 137,157 |
| 1987 | 1.18 | 2,330,407 | 76,977 | 340,821 | 139,390 |
| 1988 | 1.22 | 2,457,891 | 81,188 | 379,158 | 147,016 |
| 1989 | 1.23*** | 2,587,802 | 85,478 | 396,395 | 154,786 |
| 1990 | 1.25*** | 2,569,034 | 84,859 | 414,460 | 153,663 |
| 1991 | 1.26*** | 1,720,015 | 56,814 | 302,138 | 102,880 |

### Task 3: How many transfusion recipients were infected with HCV?

Supplementary Table 8, Supplementary Table 9, Supplementary Table 10 and Supplementary Table 11 present the estimated number of individuals infected with HCV by transfusion in England, Northern Ireland, Scotland and Wales, respectively.

Our results show 33% of the total estimated HCV infections occurring in the period of 1970 – 1979 in England, but this was just 22% in Scotland for the same period.

*Supplementary Table 8: Estimation of the annual number of individuals infected with HCV by transfusion in England, 1970 – 1991 (totals are the rounded sums of the unrounded values). The estimated numbers displayed in columns 3 and 4 for 1991 are for January to August 1991 and not the full 12-month year.*

| Year | Estimated proportion of donations that were infectious with HCV in England (Task 1) | Estimated number of units transfused in England (Task 2) | Estimated number of individuals infected by HCV by transfusion in England (Task 3) |
|---|---|---|---|
| 1970 | 0.034% | 1,100,000 | 380 (270 - 620) |
| 1971 | 0.036% | 1,180,000 | 430 (310 - 700) |
| 1972 | 0.039% | 1,250,000 | 490 (350 - 780) |
| 1973 | 0.041% | 1,340,000 | 550 (400 - 900) |
| 1974 | 0.044% | 1,420,000 | 630 (450 - 1,000) |
| 1975 | 0.047% | 1,480,000 | 700 (500 - 1,100) |
| 1976 | 0.050% | 1,660,000 | 840 (600 - 1,400) |
| 1977 | 0.054% | 1,720,000 | 940 (660 - 1,600) |
| 1978 | 0.058% | 1,860,000 | 1,100 (770 - 1,800) |
| 1979 | 0.062% | 1,910,000 | 1,200 (850 - 2,000) |
| 1980 | 0.067% | 2,010,000 | 1,300 (950 - 2,300) |
| 1981 | 0.072% | 1,900,000 | 1,300 (940 - 2,300) |
| 1982 | 0.077% | 1,930,000 | 1,400 (1,000 - 2,500) |
| 1983 | 0.082% | 2,070,000 | 1,600 (1,200 - 2,800) |
| 1984 | 0.088% | 2,170,000 | 1,800 (1,300 - 3,200) |
| 1985 | 0.039% | 2,210,000 | 830 (710 - 990) |
| 1986 | 0.041% | 2,290,000 | 910 (770 - 1,100) |
| 1987 | 0.043% | 2,340,000 | 980 (830 - 1,200) |
| 1988 | 0.044% | 2,460,000 | 1,100 (920 - 1,300) |
| 1989 | 0.046% | 2,590,000 | 1,200 (1,000 - 1,400) |
| 1990 | 0.047% | 2,570,000 | 1,200 (1,000 - 1,400) |
| 1991 | 0.049% | 1,720,000 | 840 (730 - 950) |
| Total | | **41,100,000** | **22,000 (17,000 - 32,000)** |
| Total for 1970 – 1979 | | 17,000,000 | 7,300 (5,300 - 12,000) |
| Total for 1980 – August 1991 | | 23,600,000 | 15,000 (12,000 - 20,000) |

*Supplementary Table 9: Estimation of the annual number of individuals infected with HCV by transfusion in Northern Ireland, 1970 – 1991 (totals are the rounded sums of the unrounded values). The estimated numbers displayed in columns 3 and 4 for 1991 are for January to August 1991 and not the full 12-month year.*

| Year | Estimated proportion of donations that were infectious with HCV in Northern Ireland (Task 1) | Estimated number of units transfused in Northern Ireland (Task 2) | Estimated number of individuals infected by HCV by transfusion in Northern Ireland (Task 3) |
|---|---|---|---|
| 1970 | 0.034% | 36,467 | 13 (6 - 23) |
| 1971 | 0.036% | 38,883 | 14 (7 - 26) |
| 1972 | 0.039% | 41,443 | 16 (8 - 29) |
| 1973 | 0.041% | 44,155 | 18 (10 - 33) |
| 1974 | 0.044% | 47,027 | 21 (11 - 37) |
| 1975 | 0.047% | 48,773 | 23 (13 - 41) |
| 1976 | 0.050% | 54,729 | 28 (16 - 49) |
| 1977 | 0.054% | 56,707 | 31 (18 - 55) |
| 1978 | 0.058% | 61,333 | 36 (21 - 65) |
| 1979 | 0.062% | 62,994 | 40 (24 - 71) |
| 1980 | 0.067% | 66,309 | 45 (27 - 80) |
| 1981 | 0.072% | 62,710 | 45 (27 - 79) |
| 1982 | 0.077% | 63,646 | 48 (29 - 86) |
| 1983 | 0.082% | 68,517 | 54 (33 - 99) |
| 1984 | 0.088% | 71,771 | 61 (37 - 110) |
| 1985 | 0.039% | 72,933 | 27 (17 - 40) |
| 1986 | 0.041% | 75,744 | 30 (19 - 44) |
| 1987 | 0.043% | 76,977 | 32 (20 - 46) |
| 1988 | 0.044% | 81,188 | 36 (23 - 51) |
| 1989 | 0.046% | 85,478 | 40 (26 - 56) |
| 1990 | 0.047% | 84,859 | 40 (26 - 57) |
| 1991 | 0.049% | 56,814 | 27 (17 - 40) |
| **Total** | | **1,359,456** | **730 (570 - 1,100)** |
| Total for 1970 – 1979 | | 492,510 | 240 (170 - 400) |
| Total for 1980 – August 1991 | | 866,945 | 490 (390 - 680) |

*Supplementary Table 10: Estimation of the annual number of individuals infected with HCV by transfusion in Scotland, 1970 – 1991 (totals are the rounded sums of the unrounded values). The estimated numbers displayed in columns 3 and 4 for 1991 are for January to August 1991 and not the full 12-month year.*

| **Year** | **Estimated proportion of donations that were infectious with HCV in Scotland (Task 1)** | **Estimated number of units transfused in Scotland (Task 2)** | **Estimated number of individuals infected by HCV by transfusion in Scotland (Task 3)** |
|---|---:|---:|---:|
| 1970 | 0.021% | 195,570 | 41 (27 - 58) |
| 1971 | 0.021% | 202,502 | 44 (29 - 62) |
| 1972 | 0.022% | 209,601 | 47 (32 - 66) |
| 1973 | 0.023% | 216,870 | 50 (34 - 70) |
| 1974 | 0.023% | 224,313 | 53 (36 - 75) |
| 1975 | 0.024% | 218,523 | 53 (36 - 76) |
| 1976 | 0.025% | 234,981 | 59 (41 - 84) |
| 1977 | 0.027% | 253,003 | 70 (49 - 100) |
| 1978 | 0.030% | 262,530 | 81 (57 - 120) |
| 1979 | 0.034% | 273,398 | 95 (66 - 140) |
| 1980 | 0.038% | 277,268 | 110 (75 - 170) |
| 1981 | 0.045% | 285,918 | 130 (90 - 200) |
| 1982 | 0.054% | 295,404 | 160 (110 - 260) |
| 1983 | 0.066% | 311,084 | 210 (140 - 340) |
| 1984 | 0.038% | 329,228 | 120 (95 - 160) |
| 1985 | 0.044% | 336,712 | 150 (110 - 180) |
| 1986 | 0.049% | 353,666 | 170 (140 - 220) |
| 1987 | 0.053% | 340,821 | 180 (140 - 220) |
| 1988 | 0.056% | 379,158 | 210 (170 - 260) |
| 1989 | 0.060% | 396,395 | 240 (190 - 290) |
| 1990 | 0.063% | 414,460 | 260 (210 - 320) |
| 1991 | 0.065% | 302,138 | 200 (160 - 240) |
| **Total** | | **6,312,546** | **2,700 (2,200 - 3,400)** |
| Total for 1970 – 1979 | | 2,291,292 | 590 (470 - 800) |
| Total for 1980 – August 1991 | | 4,021,254 | 2,100 (1,800 - 2,700) |

*Supplementary Table 11: Estimation of the annual number of individuals infected with HCV by transfusion in Wales, 1970 – 1991 (totals are the rounded sums of the unrounded values). The estimated numbers displayed in columns 3 and 4 for 1991 are for January to August 1991 and not the full 12-month year.*

| Year | Estimated proportion of donations that were infectious with HCV in Wales (Task 1) | Estimated number of units transfused in Wales (Task 2) | Estimated number of individuals infected by HCV by transfusion in Wales (Task 3) |
|---|---|---|---|
| 1970 | 0.034% | 66,036 | 23 (13 - 40) |
| 1971 | 0.036% | 70,409 | 26 (15 - 45) |
| 1972 | 0.039% | 75,044 | 29 (17 - 50) |
| 1973 | 0.041% | 79,955 | 33 (20 - 57) |
| 1974 | 0.044% | 85,159 | 38 (23 - 66) |
| 1975 | 0.047% | 88,321 | 42 (26 - 72) |
| 1976 | 0.050% | 99,104 | 51 (32 - 89) |
| 1977 | 0.054% | 102,686 | 56 (35 - 98) |
| 1978 | 0.058% | 111,063 | 65 (42 - 120) |
| 1979 | 0.062% | 114,071 | 73 (46 - 130) |
| 1980 | 0.067% | 120,073 | 81 (52 - 140) |
| 1981 | 0.072% | 113,557 | 81 (52 - 140) |
| 1982 | 0.077% | 115,251 | 87 (56 - 150) |
| 1983 | 0.082% | 124,073 | 99 (63 - 170) |
| 1984 | 0.088% | 129,965 | 110 (71 - 200) |
| 1985 | 0.039% | 132,068 | 50 (34 - 67) |
| 1986 | 0.041% | 137,157 | 54 (38 - 73) |
| 1987 | 0.043% | 139,390 | 58 (41 - 78) |
| 1988 | 0.044% | 147,016 | 65 (46 - 87) |
| 1989 | 0.046% | 154,786 | 72 (52 - 95) |
| 1990 | 0.047% | 153,663 | 73 (53 - 97) |
| 1991 | 0.049% | 102,880 | 50 (34 - 67) |
| **Total** | | **2,461,730** | **1,300 (1,000 - 2,000)** |
| Total for 1970 – 1979 | | 891,848 | 430 (310 - 730) |
| Total for 1980 – August 1991 | | 1,569,881 | 808 (710 - 1,200) |

*Age-sex distribution of those infected by transfusions*

We used the age-sex distribution as observed by Wallis et al. (5) in the north of England in 1994. At the time of the development of the Schnier and Goldberg model (6), these were the only data available. However, we were able to compare this to the age-sex distribution of transfusion recipients within the Scotland's National Blood Transfusion Service (SNBTS) record-linkage study in 1999. Age bands (in years) 0 and 1 - 9 years from SNBTS were pooled to be 0 - 9 and age bands 80 - 89 and 90+ from Wallis et al. were pooled to be 80+ in order to allow for direct comparison. This showed close agreement of the age-sex distribution between these two independent sources as shown in Supplementary Figure 4, giving confidence to the application of this across different settings.

Supplementary Table 12 presents the age-sex distribution of those infected in each of the four nations.

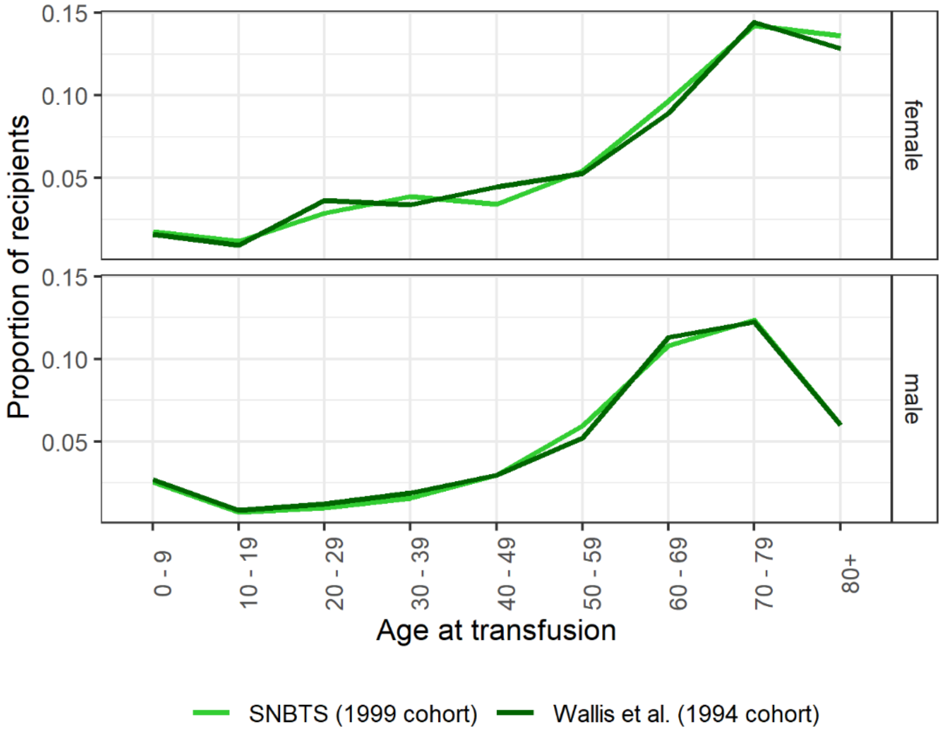

*Supplementary Figure 4: Age-sex distribution of two independent transfusion cohorts: Wallis et al. (2004) 1994 cohort from North West England, and Scottish National Blood Transfusion Service (SNBTS) 1999 cohort. Age is given in years.*

*Supplementary Table 12: Estimated age-sex distribution of those infected with HCV through transfusion in each of the four nations, 1970 – August 1991.*

| Age at transfusion (in completed years) | Proportion | | Estimated number HCV-infected | | | | | | | |
|---|---|---|---|---|---|---|---|---|---|---|
| | | | England | | Northern Ireland | | Scotland | | Wales | |
| | Females | Males | Females | Males | Females | Males | Females | Males | Females | Males |
| 0 – 9 | 0.016 (0.012 - 0.021) | 0.027 (0.021 - 0.033) | 350 (230 - 550) | 590 (420 - 910) | 12 (5 - 22) | 20 (10 - 34) | 43 (26 - 66) | 74 (49 - 110) | 21 (11 - 37) | 36 (21 - 59) |
| 10 – 19 | 0.0092 (0.0062 - 0.013) | 0.0082 (0.0054 - 0.012) | 200 (120 - 340) | 180 (110 - 310) | 7 (2 - 14) | 6 (1 - 13) | 25 (13 - 43) | 22 (11 - 38) | 12 (5 - 24) | 11 (4 - 22) |
| 20 – 29 | 0.037 (0.030 - 0.044) | 0.012 (0.0088 - 0.017) | 810 (580 - 1,200) | 280 (170 - 440) | 27 (15 - 45) | 9 (3 - 18) | 100 (70 - 140) | 34 (19-55) | 49 (30 - 78) | 16 (8 - 30) |
| 30 – 39 | 0.034 (0.028 - 0.041) | 0.019 (0.014 - 0.024) | 750 (540 - 1,100) | 420 (280 - 650) | 25 (14 - 41) | 14 (6 - 25) | 93 (64 - 130) | 52 (33 - 78) | 45 (28 - 73) | 25 (13 - 43) |
| 40 – 49 | 0.045 (0.037 - 0.052) | 0.030 (0.024 - 0.036) | 980 (730- 1,500) | 660 (470 - 1000) | 33 (19 - 53) | 22 (12 - 37) | 120 (87 - 170) | 81 (55 - 120) | 59 (38 - 93) | 39 (24 - 65) |
| 50 – 59 | 0.053 (0.045 - 0.061) | 0.052 (0.044- 0.061) | 1,200 (870 - 1,700) | 1,200 (860 - 1,700) | 39 (24 - 63) | 38 (23 - 61) | 140 (110 - 200) | 140 (100 - 190) | 70 (46 - 110) | 69 (46 - 110) |
| 60 – 69 | 0.089 (0.079 - 0.10) | 0.110 (0.100- 0.130) | 2,000 (1,500 - 2,900) | 2,500 (1,900 - 3,700) | 65 (44 - 100) | 83 (57 - 130) | 240 (190 - 320) | 310 (240 - 400) | 120 (83 - 180) | 150 (110 - 220) |
| 70 – 79 | 0.140 (0.130 - 0.160) | 0.120 (0.110 - 0.130) | 3,200 (2,500 - 4,700) | 2,700 (2,100 - 4,000) | 100 (75 - 160) | 89 (62 - 140) | 400 (310 - 510) | 340 (260 - 430) | 190 (140 - 290) | 160 (120 - 240) |
| 80 – 89 | 0.110 (0.097 - 0.120) | 0.055 (0.047 - 0.064) | 2,400 (1,800 - 3,500) | 1,200 (910 - 1,800) | 79 (54 - 120) | 40 (25 - 65) | 300 (230 - 390) | 150 (110 - 200) | 140 (100 - 220) | 74 (49 - 110) |
| 90+ | 0.020 (0.015 - 0.025) | 0.0044 (0.0024 - 0.0072) | 440 (300 - 690) | 97 (47 - 180) | 15 (7 - 26) | 3 (0 - 8) | 55 (35 - 81) | 12 (4 - 24) | 26 (14 - 45) | 6 (1 - 14) |
| Total | 0.56 (0.54 - 0.57) | 0.44 (0.43 - 0.46) | 12,000 (9,700 - 18,000) | 9,800 (7,700 - 14,000) | 400 (310 - 600) | 320 (240 - 480) | 1,500 (1,200 - 1,900) | 1,200 (990 - 1,500) | 730 (570 - 1,100) | 590 (450 - 870) |

### Task 4: How many chronically HCV-infected recipients survived 10 years after transfusion?

#### *Extending survival from 5 to 10 years post-transfusion*

The Cox proportional hazard model is based on assuming a survival function $S_i(t)$ (the probability of surviving beyond $t$ for $i$th band) of the form:

$$S_i(t) = S_0(t)^{\exp(\beta_i)},$$

where $S_0(t)$ is the survival function of a 'baseline' patient (with factors all at 0), and $\beta_i$ is the coefficient (log hazard ratio) associated with the $i$th band.

So for band $i$,

$$logS_i(t) = logS_0(t) \times e^{\beta_i},$$

and

$$\frac{logS_i(t)}{logS_0(t)} = e^{\beta_i}$$

is the hazard ratio.

So for times $t = 5{,}10$,

$$\frac{logS_i(5)}{logS_0(5)} = \frac{logS_i(10)}{logS_0(10)}$$

or

$$\frac{logS_i(10)}{logS_i(5)} = \frac{logS_0(10)}{logS_0(5)}.$$

So for all bands, $\frac{logS_i(10)}{logS_i(5)}$ should be a constant, depending on $i$.

We estimate $r_i = \frac{logS_i(10)}{logS_i(5)}$ for each age-band in the EASTR study (7) and apply this to the appropriate age-band in Wallis et al. (5).

If $S_W(5)$ is the 5-year survival in the $i$th age-band in Wallis et al., we estimate $S_W(10)$ as follows.

Assume the change from 5 to 10 years matches that in EASTR, and so:

$$r_j = \frac{logS_W(10)}{logS_W(5)}$$

or

$$S_W(10) = S_W(5)^{r_j}.$$

Estimates of $r_j$ can be found in Supplementary Table 13.

*Supplementary Table 13: Estimates of $r_i = \frac{logS_i(10)}{logS_i(5)}$ for each age-band in the EASTR study, and the age-bands within Wallis et al. that each value corresponds to.*

| Age-band (in years) in EASTR study | $r_i = \frac{logS_i(10)}{logS_i(5)}$ | | Age-bands (in years) to apply to in Wallis et al. |
|---|---|---|---|
| 16 – 24 | 1.157 | Average 1.161 | 0 – 9<br>10 – 19<br>20 – 29<br>30 – 39 |
| 25 – 39 | 1.164 | | |
| 40 – 59 | 1.237 | | 40 – 49<br>50 – 59 |
| 60 – 74 | 1.368 | | 60 – 69<br>70 – 79 |
| 75+ | 1.670 | | 80 – 89<br>90+ |

### *Comparison of Wallis et al. and EASTR*

The probability of surviving to 1, 5 and 7 years post-transfusion, independent of age-band, was broadly similar within the Wallis and EASTR cohorts, as shown in Supplementary Table 14 and Supplementary Table 15. In particular, similar values were observed for Wallis compared to recipients of red blood cells (RBC) in the EASTR study (the majority of participants in this study, 9142 RBC out of 16,958 total). We also compared the aforementioned survival probabilities across the Wallis cohort and EASTR RBC cohort using the EASTR age-bands in both settings. Again, there was broad agreement between the two sources, with the age categories 40 - 59 and 60 - 74 years appearing to have the greatest differences.

*Supplementary Table 14: Percentage of transfusion recipients surviving to 1, 5, and 7 years post-transfusion using data from the Wallis study and different blood products from the EASTR study without age stratification. RBC: red blood cells; FFP: fresh frozen plasma; PLT: platelets.*

| Data | 1-year survival (%) | 5-year survival (%) | 7-year survival (%) |
|---|---|---|---|
| Wallis – all | 67.5 | 46.8 | 41.3 |
| EASTR – RBC | 66.0 | 47.0 | 41.5 |
| EASTR – FFP | 55.0 | 41.0 | 35.3 |
| EASTR – PLT | 53.0 | 38.0 | 34.2 |

*Supplementary Table 15: Percentage of transfusion recipients surviving to 1, 5, and 7 years post-transfusion using data from the Wallis study and the EASTR study (RBC only) using the age bands (in years) in EASTR. RBC: red blood cells.*

| Age band | Sample size | | 1-year survival (%) | | 5-year survival (%) | | 7-year survival (%) | |
|---|---|---|---|---|---|---|---|---|
| | EASTR (RBC) | Wallis | EASTR (RBC) | Wallis | EASTR (RBC) | Wallis | EASTR (RBC) | Wallis |
| 16 – 24 | 288 | 91 | 93.1 | 93.4 | 89.8 | 88.9 | 88.9 | 85.4 |
| 25 – 39 | 841 | 241 | 89.4 | 89.4 | 83.7 | 82.8 | 82.7 | 80.2 |

| | | | | | | | | |
|---|---|---|---|---|---|---|---|---|
| 40 – 59 | 1,625 | 537 | 75.1 | 71.6 | 60.3 | 54.6 | 57.3 | 51.8 |
| 60 – 74 | 2,666 | 1,032 | 66.2 | 64.5 | 47.1 | 44.1 | 42.0 | 36.9 |
| 75+ | 3,255 | 846 | 54.1 | 56.9 | 27.5 | 24.4 | 20.4 | 17.7 |

Supplementary Figure 5 presents the resulting 5- and 10-year post-transfusion survival probabilities by age (years)-sex band, with the data underlying this figure presented in Supplementary Table 16.

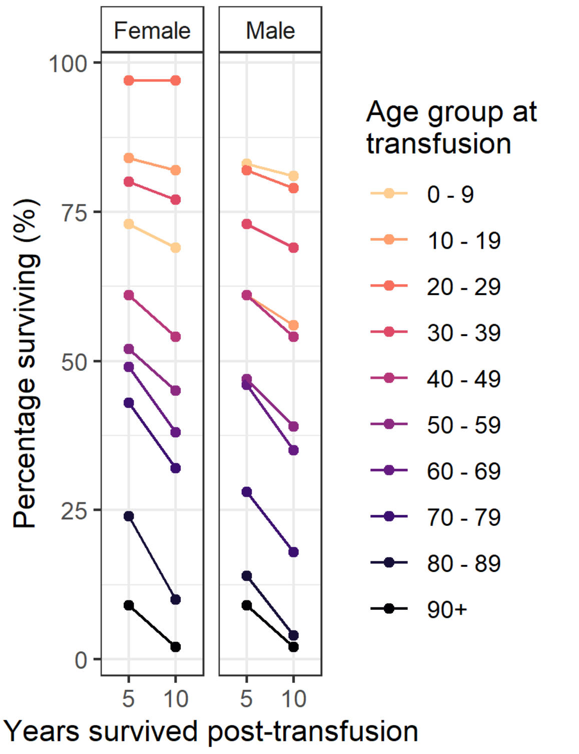

*Supplementary Figure 5: The probability of surviving to 5- and 10-years post-transfusion by age-sex band.*

*Supplementary Table 16: Estimated percentage of individuals surviving 5- and 10-years post-transfusion.*

| **Age (years)** | **5-year post-transfusion survival (%)** | | **10-year post-transfusion survival (%)** | |
|---|---|---|---|---|
| | **Female** | **Male** | **Female** | **Male** |
| 0 – 9 | 73 | 83 | 69 | 81 |
| 10 – 19 | 84 | 61 | 82 | 56 |
| 20 – 29 | 97 | 82 | 97 | 79 |
| 30 – 39 | 80 | 73 | 77 | 69 |
| 40 – 49 | 61 | 61 | 54 | 54 |
| 50 – 59 | 52 | 47 | 45 | 39 |
| 60 – 69 | 49 | 46 | 38 | 35 |
| 70 – 79 | 43 | 28 | 32 | 18 |
| 80 – 89 | 24 | 14 | 10 | 4 |
| 90+ | 9 | 9 | 2 | 2 |

Supplementary Table 17 presents the resulting number of chronically-HCV-infected persons surviving to 10-years post-transfusion.

*Supplementary Table 17: Estimated annual number of individuals with chronic HCV infection by transfusion in each nation 1970 – August 1991, and surviving 10-years post transfusion. The number of chronically infected is obtained by applying the estimated clearance rate to the number of people infected. The number surviving 10-years post transfusion is obtained by applying the survival rates in Supplementary Table 16 to the age-sex profiles in Supplementary Table 12. The estimated numbers displayed for 1991 are for January to August 1991 and not the full 12-month year.*

| Year | England | | Northern Ireland | | Scotland | | Wales | |
|---|---|---|---|---|---|---|---|---|
| | Chronically infected | Chronically infected and surviving 10-years post transfusion | Chronically infected | Chronically infected and surviving 10-years post transfusion | Chronically infected | Chronically infected and surviving 10-years post transfusion | Chronically infected | Chronically infected and surviving 10-years post transfusion |
| 1970 | 310 (220 - 500) | 120 (78 - 190) | 10 (4 - 19) | 4 (1 - 9) | 33 (21 - 49) | 12 (6 - 21) | 19 (10 - 34) | 7 (2 - 14) |
| 1971 | 360 (250 - 570) | 130 (89 - 210) | 12 (5 - 22) | 4 (1 - 10) | 36 (23 - 52) | 13 (6 - 22) | 21 (12 - 38) | 8 (3 - 16) |
| 1972 | 400 (280 - 650) | 150 (100 - 240) | 13 (6 - 25) | 5 (1 - 11) | 38 (25 - 56) | 14 (7 - 23) | 24 (14 - 42) | 9 (3 - 17) |
| 1973 | 450 (320 - 740) | 170 (120 - 280) | 15 (7 - 28) | 5 (1 - 12) | 41 (27 - 58) | 15 (8 - 24) | 27 (15 - 47) | 10 (4 - 20) |
| 1974 | 510 (360 - 840) | 190 (130 - 320) | 17 (9 - 31) | 6 (2 - 13) | 43 (29 - 63) | 16 (8 - 26) | 31 (18 - 55) | 11 (5 - 22) |
| 1975 | 570 (400 - 940) | 210 (140 - 350) | 19 (10 - 34) | 7 (2 - 14) | 43 (29 - 64) | 16 (8 - 26) | 34 (20 - 60) | 13 (6 - 24) |
| 1976 | 690 (490 - 1,100) | 260 (180 - 430) | 23 (12 - 41) | 8 (3 - 17) | 48 (32 - 70) | 18 (9 - 29) | 42 (25 - 73) | 15 (7 - 29) |
| 1977 | 770 (540 - 1,300) | 280 (190 - 480) | 25 (14 - 46) | 9 (4 - 19) | 57 (39 - 85) | 21 (12 - 34) | 46 (28 - 81) | 17 (8 - 32) |
| 1978 | 890 (630 - 1,500) | 330 (230 - 560) | 30 (17 - 54) | 11 (5 - 21) | 66 (45 - 98) | 24 (14 - 39) | 54 (34 - 95) | 20 (10 - 37) |
| 1979 | 990 (690 - 1,700) | 370 (250 - 630) | 33 (19 - 59) | 12 (5 - 24) | 78 (53 - 120) | 29 (17 -46) | 60 (37 - 100) | 22 (11 - 41) |
| 1980 | 1,100 (770 - 1,900) | 410 (280 - 690) | 37 (21 - 66) | 14 (6 - 26) | 88 (60 - 140) | 33 (20 - 54) | 66 (42 - 120) | 25 (13 - 45) |
| 1981 | 1,100 (770 - 1,900) | 410 (280 - 700) | 37 (21 - 66) | 14 (6 - 26) | 100 (72 - 170) | 39 (24 - 64) | 66 (42 - 120) | 25 (13 - 46) |
| 1982 | 1,200 (820 - 2,000) | 440 (300 - 750) | 39 (23 - 71) | 15 (6 - 28) | 130 (90 - 220) | 49 (30 - 82) | 71 (45 - 130) | 26 (14 - 49) |
| 1983 | 1,400 (940 - 2,300) | 500 (340 - 870) | 45 (27 - 81) | 17 (8 - 32) | 170 (120 - 280) | 63 (39 - 110) | 81 (51 - 140) | 30 (16 - 56) |
| 1984 | 1,500 (1,000 - 2,600) | 560 (380 - 960) | 50 (29 - 92) | 19 (9 - 36) | 100 (76 - 130) | 37 (24 - 52) | 90 (57 - 160) | 33 (19 - 62) |
| 1985 | 680 (570 - 820) | 250 (200 - 310) | 22 (13 - 34) | 8 (3 - 15) | 120 (91 - 150) | 44 (30 - 61) | 41 (27 - 57) | 15 (8 - 24) |
| 1986 | 750 (620 - 890) | 280 (220 - 340) | 25 (15 - 36) | 9 (4 - 16) | 140 (110 - 180) | 53 (36 - 72) | 45 (30 - 62) | 16 (9 - 26) |
| 1987 | 800 (670 - 960) | 300 (240 - 360) | 26 (16 - 39) | 10 (4 - 17) | 150 (110 - 190) | 54 (38 - 74) | 48 (32 - 66) | 18 (9 - 28) |
| 1988 | 890 (740 - 1,100) | 330 (270 - 400) | 29 (18 - 43) | 11 (5 - 18) | 180 (140 - 220) | 64 (46 - 86) | 53 (36 - 73) | 20 (11 - 30) |

| | | | | | | | | |
|---|---|---|---|---|---|---|---|---|
| 1989 | 990 (820 - 1,200) | 370 (300 - 450) | 32 (20 - 47) | 12 (6 - 20) | 200 (150 - 240) | 72 (51 - 96) | 59 (41 - 80) | 22 (12 - 33) |
| 1990 | 1,000 (830 - 1,200) | 370 (300 - 450) | 33 (21 - 47) | 12 (6 - 20) | 210 (170 - 270) | 79 (58 - 100) | 60 (42 - 81) | 22 (13 - 33) |
| 1991 | 690 (590 - 790) | 250 (210 - 300) | 22 (13 - 34) | 8 (3 - 15) | 160 (120 - 200) | 59 (42 - 80) | 41 (27 - 57) | 15 (8 - 24) |
| **Total** | **18,000 (14,000 - 26,000)** | **6,700 (5,200 - 9,800)** | **600 (460 - 890)** | **220 (160 - 330)** | **2,200 (1,800 - 2,800)** | **830 (660 - 1,100)** | **1,100 (830 - 1,600)** | **400 (300 - 600)** |

### Task 5: How many chronic HCV 10-year-survivors would have survived to the end of 2019, assuming no excess risk from HCV?

#### *Estimating long-term survival without additional transfusion-risk*

Long-term post-transfusion survival requires the annual hazards for each individual surviving from 10 years-post transfusion until the end of 2019. (Note that 2019 can be substituted for preceding years of interest during the study period using this methodology). Accounting for the 10 age-bands at transfusion, two sexes, 21 transfusion years and all possible years between 1980 (10 years after the first transfusion year) and 2019, 12,980 hazards were required to be extracted from 38 life-tables (in three-year intervals from 1980 - 1982 until 2017 - 2019) for each nation individually. Note that for the year 1980, the lifetable used was 1980 - 1982 rather than 1979 - 1981 which was not available online. Similarly, for 2019 the life-table used was 2017 - 2019 rather than 2018 - 2020 which was likely obscured by changes in mortality caused, directly and indirectly, by the COVID-19 pandemic. For each nation, the methodology was applied to the national life-tables as published by the Office for National Statistics (ONS).

An illustrative example is now provided. Consider a female aged 40 - 49 transfused in England in 1985. This female's age can be represented as 45 (years) (the mean and median of this age band). The likelihood of surviving 10 years post-transfusion was already accounted for, and so 10 years later in 1995, this female is 55. There are 24 years (2019 - 1995) that this female has to survive in order to still be alive in 2019, with this hazard changing annually both due to their ageing and also to revisions in life-expectancy within life-tables. Supplementary Table 18 illustrates the hazards to be extracted relevant to this female.

By denoting $s_x = 1 - q_x$ as the probability of surviving each year, the probability of surviving to the end of 2019 is the product of all $s_x$ for the number of years between transfusion and 2019. In the example above, this is 24 years and thus 24 values of $s_x$ contributing to overall probability of surviving to the end of 2019.

The resulting survival probabilities can be seen in Supplementary Table 19 - Supplementary Table 26.

*Supplementary Table 18: An illustrative example of the hazards required to estimate the probability of survival to 2019 for a female aged 40 – 49 at transfusion in 1985.*

| **Years since transfusion** | **Calendar year between 10-years post transfusion and 2019** | **Age (year) in this calendar year** | **Life-table to extract from** | **Hazard ($q_x$)** |
|---|---|---|---|---|
| 10 | 1995 | 55 | 1994 – 1996 | 0.004461 |
| 11 | 1996 | 56 | 1995 – 1997 | 0.004757 |
| 12 | 1997 | 57 | 1996 – 1998 | 0.005295 |
| … | … | … | … | … |
| 32 | 2017 | 77 | 2016 – 2018 | 0.039053 |
| 33 | 2018 | 78 | 2017 – 2019 | 0.042587 |
| 34 | 2019 | 79 | 2017 – 2019 | 0.046928 |

*Supplementary Table 19: Probability of a female in England in each age-band and transfusion year surviving to the end of 2019 without assuming excess post-transfusion hazard beyond the first 10 years.*

| **Age band at transfusion (years)** | | **0 - 9** | **10 - 19** | **20 - 29** | **30 - 39** | **40 - 49** | **50 - 59** | **60 - 69** | **70 - 79** | **80 - 89** | **90+** |
|---|---|---|---|---|---|---|---|---|---|---|---|
| **Midpoint age (years)** | | 5 | 15 | 25 | 35 | 45 | 55 | 65 | 75 | 85 | 95 |
| **Midpoint age after surviving 10 years (years)** | | 15 | 25 | 35 | 45 | 55 | 65 | 75 | 85 | 95 | 105 |
| ***Transfusion year*** | **1970** | 0.96 | 0.91 | 0.80 | 0.52 | 0.11 | 0.00 | 0.00 | 0.00 | 0.00 | 0.00 |
| | **1971** | 0.97 | 0.92 | 0.82 | 0.56 | 0.14 | 0.00 | 0.00 | 0.00 | 0.00 | 0.00 |
| | **1972** | 0.97 | 0.93 | 0.84 | 0.60 | 0.17 | 0.00 | 0.00 | 0.00 | 0.00 | 0.00 |
| | **1973** | 0.97 | 0.93 | 0.85 | 0.63 | 0.21 | 0.00 | 0.00 | 0.00 | 0.00 | 0.00 |
| | **1974** | 0.97 | 0.94 | 0.86 | 0.66 | 0.25 | 0.01 | 0.00 | 0.00 | 0.00 | 0.00 |
| | **1975** | 0.98 | 0.95 | 0.87 | 0.69 | 0.30 | 0.02 | 0.00 | 0.00 | 0.00 | 0.00 |
| | **1976** | 0.98 | 0.95 | 0.88 | 0.72 | 0.35 | 0.03 | 0.00 | 0.00 | 0.00 | 0.00 |
| | **1977** | 0.98 | 0.96 | 0.89 | 0.74 | 0.40 | 0.05 | 0.00 | 0.00 | 0.00 | 0.00 |
| | **1978** | 0.98 | 0.96 | 0.90 | 0.77 | 0.44 | 0.06 | 0.00 | 0.00 | 0.00 | 0.00 |
| | **1979** | 0.98 | 0.96 | 0.91 | 0.79 | 0.49 | 0.09 | 0.00 | 0.00 | 0.00 | 0.00 |
| | **1980** | 0.98 | 0.97 | 0.92 | 0.81 | 0.53 | 0.12 | 0.00 | 0.00 | 0.00 | 0.00 |
| | **1981** | 0.99 | 0.97 | 0.93 | 0.83 | 0.58 | 0.15 | 0.00 | 0.00 | 0.00 | 0.00 |
| | **1982** | 0.99 | 0.97 | 0.93 | 0.84 | 0.62 | 0.19 | 0.00 | 0.00 | 0.00 | 0.00 |
| | **1983** | 0.99 | 0.97 | 0.94 | 0.86 | 0.65 | 0.23 | 0.00 | 0.00 | 0.00 | 0.00 |
| | **1984** | 0.99 | 0.98 | 0.95 | 0.87 | 0.68 | 0.28 | 0.02 | 0.00 | 0.00 | 0.00 |
| | **1985** | 0.99 | 0.98 | 0.95 | 0.88 | 0.71 | 0.33 | 0.03 | 0.00 | 0.00 | 0.00 |
| | **1986** | 0.99 | 0.98 | 0.96 | 0.89 | 0.74 | 0.38 | 0.04 | 0.00 | 0.00 | 0.00 |
| | **1987** | 0.99 | 0.98 | 0.96 | 0.90 | 0.77 | 0.43 | 0.06 | 0.00 | 0.00 | 0.00 |
| | **1988** | 0.99 | 0.98 | 0.96 | 0.91 | 0.79 | 0.48 | 0.08 | 0.00 | 0.00 | 0.00 |
| | **1989** | 0.99 | 0.99 | 0.97 | 0.92 | 0.81 | 0.53 | 0.11 | 0.00 | 0.00 | 0.00 |
| | **1990** | 0.99 | 0.99 | 0.97 | 0.93 | 0.83 | 0.57 | 0.14 | 0.00 | 0.00 | 0.00 |
| | **1991** | 0.99 | 0.99 | 0.97 | 0.94 | 0.85 | 0.62 | 0.18 | 0.00 | 0.00 | 0.00 |

*Supplementary Table 20: Probability of a male in England in each age-band and transfusion year surviving to the end of 2019 without assuming excess post-transfusion hazard beyond the first 10 years.*

| **Age band at transfusion (years)** | | **0 - 9** | **10 - 19** | **20 - 29** | **30 - 39** | **40 - 49** | **50 - 59** | **60 - 69** | **70 - 79** | **80 - 89** | **90+** |
|---|---|---|---|---|---|---|---|---|---|---|---|
| **Midpoint age (years)** | | 5 | 15 | 25 | 35 | 45 | 55 | 65 | 75 | 85 | 95 |
| **Midpoint age after surviving 10 years (years)** | | 15 | 25 | 35 | 45 | 55 | 65 | 75 | 85 | 95 | 105 |
| **Transfusion year** | **1970** | 0.94 | 0.87 | 0.71 | 0.38 | 0.05 | 0.00 | 0.00 | 0.00 | 0.00 | 0.00 |
| | **1971** | 0.94 | 0.88 | 0.74 | 0.42 | 0.07 | 0.00 | 0.00 | 0.00 | 0.00 | 0.00 |
| | **1972** | 0.95 | 0.89 | 0.76 | 0.46 | 0.09 | 0.00 | 0.00 | 0.00 | 0.00 | 0.00 |
| | **1973** | 0.95 | 0.90 | 0.78 | 0.50 | 0.11 | 0.00 | 0.00 | 0.00 | 0.00 | 0.00 |
| | **1974** | 0.95 | 0.91 | 0.80 | 0.53 | 0.15 | 0.00 | 0.00 | 0.00 | 0.00 | 0.00 |
| | **1975** | 0.96 | 0.92 | 0.81 | 0.56 | 0.18 | 0.01 | 0.00 | 0.00 | 0.00 | 0.00 |
| | **1976** | 0.96 | 0.92 | 0.83 | 0.60 | 0.22 | 0.01 | 0.00 | 0.00 | 0.00 | 0.00 |
| | **1977** | 0.96 | 0.93 | 0.84 | 0.63 | 0.26 | 0.02 | 0.00 | 0.00 | 0.00 | 0.00 |
| | **1978** | 0.96 | 0.94 | 0.85 | 0.67 | 0.30 | 0.03 | 0.00 | 0.00 | 0.00 | 0.00 |
| | **1979** | 0.97 | 0.94 | 0.87 | 0.70 | 0.35 | 0.04 | 0.00 | 0.00 | 0.00 | 0.00 |
| | **1980** | 0.97 | 0.94 | 0.88 | 0.72 | 0.40 | 0.06 | 0.00 | 0.00 | 0.00 | 0.00 |
| | **1981** | 0.97 | 0.95 | 0.89 | 0.75 | 0.44 | 0.08 | 0.00 | 0.00 | 0.00 | 0.00 |
| | **1982** | 0.97 | 0.95 | 0.90 | 0.77 | 0.48 | 0.10 | 0.00 | 0.00 | 0.00 | 0.00 |
| | **1983** | 0.98 | 0.96 | 0.91 | 0.79 | 0.52 | 0.13 | 0.00 | 0.00 | 0.00 | 0.00 |
| | **1984** | 0.98 | 0.96 | 0.92 | 0.81 | 0.56 | 0.17 | 0.01 | 0.00 | 0.00 | 0.00 |
| | **1985** | 0.98 | 0.96 | 0.93 | 0.82 | 0.59 | 0.21 | 0.01 | 0.00 | 0.00 | 0.00 |
| | **1986** | 0.98 | 0.97 | 0.93 | 0.84 | 0.63 | 0.25 | 0.02 | 0.00 | 0.00 | 0.00 |
| | **1987** | 0.98 | 0.97 | 0.94 | 0.85 | 0.66 | 0.30 | 0.03 | 0.00 | 0.00 | 0.00 |
| | **1988** | 0.98 | 0.97 | 0.95 | 0.87 | 0.70 | 0.35 | 0.04 | 0.00 | 0.00 | 0.00 |
| | **1989** | 0.99 | 0.97 | 0.95 | 0.88 | 0.73 | 0.40 | 0.06 | 0.00 | 0.00 | 0.00 |
| | **1990** | 0.99 | 0.98 | 0.95 | 0.89 | 0.75 | 0.45 | 0.08 | 0.00 | 0.00 | 0.00 |
| | **1991** | 0.99 | 0.98 | 0.96 | 0.90 | 0.78 | 0.49 | 0.11 | 0.00 | 0.00 | 0.00 |

*Supplementary Table 21: Probability of a female in Northern Ireland in each age-band and transfusion year surviving to the end of 2019 without assuming excess post-transfusion hazard beyond the first 10 years.*

| **Age band at transfusion (years)** | | **0 - 9** | **10 - 19** | **20 - 29** | **30 - 39** | **40 - 49** | **50 - 59** | **60 - 69** | **70 - 79** | **80 - 89** | **90+** |
|---|---|---|---|---|---|---|---|---|---|---|---|
| **Midpoint age (years)** | | 5 | 15 | 25 | 35 | 45 | 55 | 65 | 75 | 85 | 95 |
| **Midpoint age after surviving 10 years (years)** | | 15 | 25 | 35 | 45 | 55 | 65 | 75 | 85 | 95 | 105 |
| **Transfusion year** | **1970** | 0.96 | 0.90 | 0.79 | 0.50 | 0.09 | 0.00 | 0.00 | 0.00 | 0.00 | 0.00 |
| | **1971** | 0.97 | 0.91 | 0.81 | 0.54 | 0.13 | 0.00 | 0.00 | 0.00 | 0.00 | 0.00 |
| | **1972** | 0.97 | 0.92 | 0.82 | 0.57 | 0.16 | 0.00 | 0.00 | 0.00 | 0.00 | 0.00 |
| | **1973** | 0.97 | 0.93 | 0.84 | 0.61 | 0.20 | 0.00 | 0.00 | 0.00 | 0.00 | 0.00 |
| | **1974** | 0.97 | 0.94 | 0.85 | 0.64 | 0.24 | 0.01 | 0.00 | 0.00 | 0.00 | 0.00 |
| | **1975** | 0.97 | 0.94 | 0.86 | 0.68 | 0.28 | 0.02 | 0.00 | 0.00 | 0.00 | 0.00 |
| | **1976** | 0.98 | 0.95 | 0.88 | 0.71 | 0.32 | 0.03 | 0.00 | 0.00 | 0.00 | 0.00 |
| | **1977** | 0.98 | 0.95 | 0.89 | 0.73 | 0.37 | 0.04 | 0.00 | 0.00 | 0.00 | 0.00 |
| | **1978** | 0.98 | 0.96 | 0.89 | 0.75 | 0.42 | 0.06 | 0.00 | 0.00 | 0.00 | 0.00 |
| | **1979** | 0.98 | 0.96 | 0.90 | 0.77 | 0.47 | 0.08 | 0.00 | 0.00 | 0.00 | 0.00 |
| | **1980** | 0.99 | 0.97 | 0.91 | 0.80 | 0.52 | 0.10 | 0.00 | 0.00 | 0.00 | 0.00 |
| | **1981** | 0.99 | 0.97 | 0.92 | 0.82 | 0.56 | 0.14 | 0.00 | 0.00 | 0.00 | 0.00 |
| | **1982** | 0.99 | 0.97 | 0.93 | 0.83 | 0.59 | 0.17 | 0.00 | 0.00 | 0.00 | 0.00 |
| | **1983** | 0.99 | 0.97 | 0.93 | 0.85 | 0.63 | 0.22 | 0.00 | 0.00 | 0.00 | 0.00 |
| | **1984** | 0.99 | 0.98 | 0.94 | 0.86 | 0.66 | 0.26 | 0.02 | 0.00 | 0.00 | 0.00 |
| | **1985** | 0.99 | 0.98 | 0.94 | 0.87 | 0.70 | 0.31 | 0.02 | 0.00 | 0.00 | 0.00 |
| | **1986** | 0.99 | 0.98 | 0.95 | 0.88 | 0.73 | 0.36 | 0.03 | 0.00 | 0.00 | 0.00 |
| | **1987** | 0.99 | 0.98 | 0.96 | 0.90 | 0.75 | 0.41 | 0.05 | 0.00 | 0.00 | 0.00 |
| | **1988** | 0.99 | 0.98 | 0.96 | 0.90 | 0.77 | 0.46 | 0.07 | 0.00 | 0.00 | 0.00 |
| | **1989** | 0.99 | 0.99 | 0.97 | 0.91 | 0.80 | 0.51 | 0.10 | 0.00 | 0.00 | 0.00 |
| | **1990** | 0.99 | 0.99 | 0.97 | 0.92 | 0.82 | 0.56 | 0.13 | 0.00 | 0.00 | 0.00 |
| | **1991** | 0.99 | 0.99 | 0.97 | 0.93 | 0.84 | 0.60 | 0.17 | 0.00 | 0.00 | 0.00 |

*Supplementary Table 22: Probability of a male in Northern Ireland in each age-band and transfusion year surviving to the end of 2019 without assuming excess post-transfusion hazard beyond the first 10 years.*

| **Age band at transfusion (years)** | | **0 - 9** | **10 - 19** | **20 - 29** | **30 - 39** | **40 - 49** | **50 - 59** | **60 - 69** | **70 - 79** | **80 - 89** | **90+** |
|---|---|---|---|---|---|---|---|---|---|---|---|
| **Midpoint age (years)** | | 5 | 15 | 25 | 35 | 45 | 55 | 65 | 75 | 85 | 95 |
| **Midpoint age after surviving 10 years (years)** | | 15 | 25 | 35 | 45 | 55 | 65 | 75 | 85 | 95 | 105 |
| **Transfusion year** | **1970** | 0.93 | 0.86 | 0.69 | 0.34 | 0.04 | 0.00 | 0.00 | 0.00 | 0.00 | 0.00 |
| | **1971** | 0.93 | 0.87 | 0.72 | 0.39 | 0.05 | 0.00 | 0.00 | 0.00 | 0.00 | 0.00 |
| | **1972** | 0.94 | 0.89 | 0.74 | 0.43 | 0.07 | 0.00 | 0.00 | 0.00 | 0.00 | 0.00 |
| | **1973** | 0.94 | 0.89 | 0.76 | 0.47 | 0.10 | 0.00 | 0.00 | 0.00 | 0.00 | 0.00 |
| | **1974** | 0.95 | 0.90 | 0.79 | 0.51 | 0.12 | 0.00 | 0.00 | 0.00 | 0.00 | 0.00 |
| | **1975** | 0.95 | 0.91 | 0.80 | 0.55 | 0.16 | 0.01 | 0.00 | 0.00 | 0.00 | 0.00 |
| | **1976** | 0.95 | 0.92 | 0.82 | 0.58 | 0.19 | 0.01 | 0.00 | 0.00 | 0.00 | 0.00 |
| | **1977** | 0.96 | 0.92 | 0.83 | 0.62 | 0.24 | 0.01 | 0.00 | 0.00 | 0.00 | 0.00 |
| | **1978** | 0.96 | 0.93 | 0.85 | 0.64 | 0.28 | 0.02 | 0.00 | 0.00 | 0.00 | 0.00 |
| | **1979** | 0.96 | 0.94 | 0.86 | 0.67 | 0.32 | 0.03 | 0.00 | 0.00 | 0.00 | 0.00 |
| | **1980** | 0.96 | 0.94 | 0.87 | 0.70 | 0.36 | 0.05 | 0.00 | 0.00 | 0.00 | 0.00 |
| | **1981** | 0.97 | 0.94 | 0.88 | 0.73 | 0.41 | 0.06 | 0.00 | 0.00 | 0.00 | 0.00 |
| | **1982** | 0.97 | 0.95 | 0.90 | 0.76 | 0.45 | 0.09 | 0.00 | 0.00 | 0.00 | 0.00 |
| | **1983** | 0.97 | 0.95 | 0.91 | 0.78 | 0.50 | 0.12 | 0.00 | 0.00 | 0.00 | 0.00 |
| | **1984** | 0.97 | 0.96 | 0.91 | 0.80 | 0.54 | 0.15 | 0.01 | 0.00 | 0.00 | 0.00 |
| | **1985** | 0.97 | 0.96 | 0.92 | 0.82 | 0.58 | 0.18 | 0.01 | 0.00 | 0.00 | 0.00 |
| | **1986** | 0.98 | 0.96 | 0.93 | 0.83 | 0.61 | 0.22 | 0.01 | 0.00 | 0.00 | 0.00 |
| | **1987** | 0.98 | 0.97 | 0.93 | 0.85 | 0.65 | 0.28 | 0.02 | 0.00 | 0.00 | 0.00 |
| | **1988** | 0.98 | 0.97 | 0.94 | 0.86 | 0.67 | 0.32 | 0.03 | 0.00 | 0.00 | 0.00 |
| | **1989** | 0.98 | 0.97 | 0.95 | 0.87 | 0.70 | 0.37 | 0.05 | 0.00 | 0.00 | 0.00 |
| | **1990** | 0.98 | 0.97 | 0.95 | 0.89 | 0.73 | 0.42 | 0.07 | 0.00 | 0.00 | 0.00 |
| | **1991** | 0.98 | 0.98 | 0.95 | 0.90 | 0.77 | 0.47 | 0.09 | 0.00 | 0.00 | 0.00 |

*Supplementary Table 23: Probability of a female in Scotland in each age-band and transfusion year surviving to the end of 2019 without assuming excess post-transfusion hazard beyond the first 10 years.*

| **Age band at transfusion (years)** | | **0 - 9** | **10 - 19** | **20 - 29** | **30 - 39** | **40 - 49** | **50 - 59** | **60 - 69** | **70 - 79** | **80 - 89** | **90+** |
|---|---|---|---|---|---|---|---|---|---|---|---|
| **Midpoint age (years)** | | 5 | 15 | 25 | 35 | 45 | 55 | 65 | 75 | 85 | 95 |
| **Midpoint age after surviving 10 years (years)** | | 15 | 25 | 35 | 45 | 55 | 65 | 75 | 85 | 95 | 105 |
| **Transfusion year** | **1970** | 0.95 | 0.89 | 0.75 | 0.44 | 0.08 | 0.00 | 0.00 | 0.00 | 0.00 | 0.00 |
| | **1971** | 0.96 | 0.90 | 0.77 | 0.48 | 0.10 | 0.00 | 0.00 | 0.00 | 0.00 | 0.00 |
| | **1972** | 0.96 | 0.91 | 0.79 | 0.52 | 0.13 | 0.00 | 0.00 | 0.00 | 0.00 | 0.00 |
| | **1973** | 0.96 | 0.92 | 0.81 | 0.55 | 0.16 | 0.00 | 0.00 | 0.00 | 0.00 | 0.00 |
| | **1974** | 0.97 | 0.93 | 0.83 | 0.59 | 0.20 | 0.01 | 0.00 | 0.00 | 0.00 | 0.00 |
| | **1975** | 0.97 | 0.93 | 0.84 | 0.62 | 0.24 | 0.02 | 0.00 | 0.00 | 0.00 | 0.00 |
| | **1976** | 0.97 | 0.94 | 0.85 | 0.65 | 0.28 | 0.02 | 0.00 | 0.00 | 0.00 | 0.00 |
| | **1977** | 0.97 | 0.94 | 0.87 | 0.68 | 0.33 | 0.03 | 0.00 | 0.00 | 0.00 | 0.00 |
| | **1978** | 0.98 | 0.95 | 0.88 | 0.71 | 0.37 | 0.05 | 0.00 | 0.00 | 0.00 | 0.00 |
| | **1979** | 0.98 | 0.95 | 0.89 | 0.74 | 0.42 | 0.06 | 0.00 | 0.00 | 0.00 | 0.00 |
| | **1980** | 0.98 | 0.96 | 0.90 | 0.76 | 0.46 | 0.09 | 0.00 | 0.00 | 0.00 | 0.00 |
| | **1981** | 0.98 | 0.96 | 0.91 | 0.79 | 0.50 | 0.11 | 0.00 | 0.00 | 0.00 | 0.00 |
| | **1982** | 0.98 | 0.96 | 0.91 | 0.81 | 0.54 | 0.14 | 0.00 | 0.00 | 0.00 | 0.00 |
| | **1983** | 0.98 | 0.97 | 0.92 | 0.82 | 0.58 | 0.18 | 0.00 | 0.00 | 0.00 | 0.00 |
| | **1984** | 0.98 | 0.97 | 0.93 | 0.84 | 0.61 | 0.22 | 0.01 | 0.00 | 0.00 | 0.00 |
| | **1985** | 0.99 | 0.97 | 0.94 | 0.85 | 0.64 | 0.26 | 0.02 | 0.00 | 0.00 | 0.00 |
| | **1986** | 0.99 | 0.97 | 0.94 | 0.87 | 0.68 | 0.31 | 0.03 | 0.00 | 0.00 | 0.00 |
| | **1987** | 0.99 | 0.98 | 0.95 | 0.88 | 0.71 | 0.36 | 0.04 | 0.00 | 0.00 | 0.00 |
| | **1988** | 0.99 | 0.98 | 0.95 | 0.89 | 0.74 | 0.41 | 0.06 | 0.00 | 0.00 | 0.00 |
| | **1989** | 0.99 | 0.98 | 0.96 | 0.90 | 0.76 | 0.46 | 0.08 | 0.00 | 0.00 | 0.00 |
| | **1990** | 0.99 | 0.98 | 0.96 | 0.91 | 0.79 | 0.51 | 0.11 | 0.00 | 0.00 | 0.00 |
| | **1991** | 0.99 | 0.98 | 0.97 | 0.92 | 0.81 | 0.55 | 0.15 | 0.00 | 0.00 | 0.00 |

*Supplementary Table 24: Probability of a male in Scotland in each age-band and transfusion year surviving to the end of 2019 without assuming excess post-transfusion hazard beyond the first 10 years.*

| **Age band at transfusion (years)** | | **0 - 9** | **10 - 19** | **20 - 29** | **30 - 39** | **40 - 49** | **50 - 59** | **60 - 69** | **70 - 79** | **80 - 89** | **90+** |
|---|---|---|---|---|---|---|---|---|---|---|---|
| **Midpoint age (years)** | | 5 | 15 | 25 | 35 | 45 | 55 | 65 | 75 | 85 | 95 |
| **Midpoint age after surviving 10 years (years)** | | 15 | 25 | 35 | 45 | 55 | 65 | 75 | 85 | 95 | 105 |
| **Transfusion year** | **1970** | 0.92 | 0.84 | 0.64 | 0.30 | 0.03 | 0.00 | 0.00 | 0.00 | 0.00 | 0.00 |
| | **1971** | 0.92 | 0.85 | 0.68 | 0.34 | 0.05 | 0.00 | 0.00 | 0.00 | 0.00 | 0.00 |
| | **1972** | 0.92 | 0.86 | 0.70 | 0.38 | 0.06 | 0.00 | 0.00 | 0.00 | 0.00 | 0.00 |
| | **1973** | 0.93 | 0.87 | 0.73 | 0.41 | 0.08 | 0.00 | 0.00 | 0.00 | 0.00 | 0.00 |
| | **1974** | 0.93 | 0.88 | 0.75 | 0.45 | 0.11 | 0.00 | 0.00 | 0.00 | 0.00 | 0.00 |
| | **1975** | 0.94 | 0.89 | 0.77 | 0.48 | 0.14 | 0.01 | 0.00 | 0.00 | 0.00 | 0.00 |
| | **1976** | 0.94 | 0.90 | 0.79 | 0.52 | 0.17 | 0.01 | 0.00 | 0.00 | 0.00 | 0.00 |
| | **1977** | 0.94 | 0.91 | 0.80 | 0.55 | 0.21 | 0.01 | 0.00 | 0.00 | 0.00 | 0.00 |
| | **1978** | 0.95 | 0.91 | 0.82 | 0.59 | 0.24 | 0.02 | 0.00 | 0.00 | 0.00 | 0.00 |
| | **1979** | 0.95 | 0.92 | 0.83 | 0.62 | 0.28 | 0.03 | 0.00 | 0.00 | 0.00 | 0.00 |
| | **1980** | 0.95 | 0.92 | 0.85 | 0.66 | 0.32 | 0.04 | 0.00 | 0.00 | 0.00 | 0.00 |
| | **1981** | 0.95 | 0.93 | 0.86 | 0.69 | 0.36 | 0.06 | 0.00 | 0.00 | 0.00 | 0.00 |
| | **1982** | 0.96 | 0.93 | 0.87 | 0.72 | 0.40 | 0.07 | 0.00 | 0.00 | 0.00 | 0.00 |
| | **1983** | 0.96 | 0.94 | 0.88 | 0.75 | 0.44 | 0.10 | 0.00 | 0.00 | 0.00 | 0.00 |
| | **1984** | 0.96 | 0.94 | 0.89 | 0.76 | 0.48 | 0.13 | 0.01 | 0.00 | 0.00 | 0.00 |
| | **1985** | 0.97 | 0.94 | 0.90 | 0.78 | 0.51 | 0.16 | 0.01 | 0.00 | 0.00 | 0.00 |
| | **1986** | 0.97 | 0.95 | 0.91 | 0.80 | 0.55 | 0.20 | 0.01 | 0.00 | 0.00 | 0.00 |
| | **1987** | 0.97 | 0.95 | 0.92 | 0.82 | 0.59 | 0.25 | 0.02 | 0.00 | 0.00 | 0.00 |
| | **1988** | 0.97 | 0.95 | 0.93 | 0.84 | 0.63 | 0.29 | 0.03 | 0.00 | 0.00 | 0.00 |
| | **1989** | 0.98 | 0.96 | 0.93 | 0.85 | 0.66 | 0.33 | 0.05 | 0.00 | 0.00 | 0.00 |
| | **1990** | 0.98 | 0.96 | 0.94 | 0.87 | 0.70 | 0.38 | 0.06 | 0.00 | 0.00 | 0.00 |
| | **1991** | 0.98 | 0.96 | 0.94 | 0.88 | 0.73 | 0.42 | 0.09 | 0.00 | 0.00 | 0.00 |

*Supplementary Table 25: Probability of a female in Wales in each age-band and transfusion year surviving to the end of 2019 without assuming excess post-transfusion hazard beyond the first 10 years.*

| **Age band at transfusion (years)** | | **0 - 9** | **10 - 19** | **20 - 29** | **30 - 39** | **40 - 49** | **50 - 59** | **60 - 69** | **70 - 79** | **80 - 89** | **90+** |
|---|---|---|---|---|---|---|---|---|---|---|---|
| **Midpoint age (years)** | | 5 | 15 | 25 | 35 | 45 | 55 | 65 | 75 | 85 | 95 |
| **Midpoint age after surviving 10 years (years)** | | 15 | 25 | 35 | 45 | 55 | 65 | 75 | 85 | 95 | 105 |
| **Transfusion year** | **1970** | 0.96 | 0.90 | 0.78 | 0.49 | 0.10 | 0.00 | 0.00 | 0.00 | 0.00 | 0.00 |
| | **1971** | 0.96 | 0.91 | 0.80 | 0.53 | 0.12 | 0.00 | 0.00 | 0.00 | 0.00 | 0.00 |
| | **1972** | 0.97 | 0.92 | 0.82 | 0.57 | 0.16 | 0.00 | 0.00 | 0.00 | 0.00 | 0.00 |
| | **1973** | 0.97 | 0.93 | 0.84 | 0.61 | 0.19 | 0.00 | 0.00 | 0.00 | 0.00 | 0.00 |
| | **1974** | 0.97 | 0.93 | 0.85 | 0.64 | 0.23 | 0.01 | 0.00 | 0.00 | 0.00 | 0.00 |
| | **1975** | 0.97 | 0.94 | 0.86 | 0.67 | 0.28 | 0.02 | 0.00 | 0.00 | 0.00 | 0.00 |
| | **1976** | 0.98 | 0.95 | 0.87 | 0.70 | 0.33 | 0.03 | 0.00 | 0.00 | 0.00 | 0.00 |
| | **1977** | 0.98 | 0.95 | 0.88 | 0.73 | 0.37 | 0.04 | 0.00 | 0.00 | 0.00 | 0.00 |
| | **1978** | 0.98 | 0.95 | 0.89 | 0.75 | 0.42 | 0.06 | 0.00 | 0.00 | 0.00 | 0.00 |
| | **1979** | 0.98 | 0.96 | 0.90 | 0.77 | 0.47 | 0.08 | 0.00 | 0.00 | 0.00 | 0.00 |
| | **1980** | 0.98 | 0.96 | 0.91 | 0.79 | 0.51 | 0.11 | 0.00 | 0.00 | 0.00 | 0.00 |
| | **1981** | 0.98 | 0.97 | 0.92 | 0.81 | 0.55 | 0.14 | 0.00 | 0.00 | 0.00 | 0.00 |
| | **1982** | 0.98 | 0.97 | 0.92 | 0.83 | 0.59 | 0.17 | 0.00 | 0.00 | 0.00 | 0.00 |
| | **1983** | 0.99 | 0.97 | 0.93 | 0.85 | 0.63 | 0.21 | 0.00 | 0.00 | 0.00 | 0.00 |
| | **1984** | 0.99 | 0.97 | 0.94 | 0.86 | 0.66 | 0.26 | 0.02 | 0.00 | 0.00 | 0.00 |
| | **1985** | 0.99 | 0.98 | 0.94 | 0.87 | 0.69 | 0.30 | 0.02 | 0.00 | 0.00 | 0.00 |
| | **1986** | 0.99 | 0.98 | 0.95 | 0.88 | 0.72 | 0.36 | 0.04 | 0.00 | 0.00 | 0.00 |
| | **1987** | 0.99 | 0.98 | 0.95 | 0.89 | 0.75 | 0.41 | 0.05 | 0.00 | 0.00 | 0.00 |
| | **1988** | 0.99 | 0.98 | 0.96 | 0.90 | 0.77 | 0.46 | 0.07 | 0.00 | 0.00 | 0.00 |
| | **1989** | 0.99 | 0.98 | 0.96 | 0.91 | 0.79 | 0.51 | 0.10 | 0.00 | 0.00 | 0.00 |
| | **1990** | 0.99 | 0.99 | 0.97 | 0.92 | 0.81 | 0.55 | 0.13 | 0.00 | 0.00 | 0.00 |
| | **1991** | 0.99 | 0.99 | 0.97 | 0.93 | 0.84 | 0.60 | 0.17 | 0.00 | 0.00 | 0.00 |

*Supplementary Table 26: Probability of a male in Wales in each age-band and transfusion year surviving to the end of 2019 without assuming excess post-transfusion hazard beyond the first 10 years.*

| **Age band at transfusion (years)** | | **0 - 9** | **10 - 19** | **20 - 29** | **30 - 39** | **40 - 49** | **50 - 59** | **60 - 69** | **70 - 79** | **80 - 89** | **90+** |
|---|---|---|---|---|---|---|---|---|---|---|---|
| **Midpoint age (years)** | | 5 | 15 | 25 | 35 | 45 | 55 | 65 | 75 | 85 | 95 |
| **Midpoint age after surviving 10 years (years)** | | 15 | 25 | 35 | 45 | 55 | 65 | 75 | 85 | 95 | 105 |
| **Transfusion year** | **1970** | 0.93 | 0.86 | 0.69 | 0.35 | 0.04 | 0.00 | 0.00 | 0.00 | 0.00 | 0.00 |
| | **1971** | 0.93 | 0.87 | 0.72 | 0.39 | 0.06 | 0.00 | 0.00 | 0.00 | 0.00 | 0.00 |
| | **1972** | 0.94 | 0.88 | 0.75 | 0.43 | 0.08 | 0.00 | 0.00 | 0.00 | 0.00 | 0.00 |
| | **1973** | 0.94 | 0.90 | 0.77 | 0.47 | 0.10 | 0.00 | 0.00 | 0.00 | 0.00 | 0.00 |
| | **1974** | 0.95 | 0.90 | 0.78 | 0.51 | 0.13 | 0.00 | 0.00 | 0.00 | 0.00 | 0.00 |
| | **1975** | 0.95 | 0.91 | 0.80 | 0.54 | 0.16 | 0.01 | 0.00 | 0.00 | 0.00 | 0.00 |
| | **1976** | 0.95 | 0.92 | 0.82 | 0.58 | 0.20 | 0.01 | 0.00 | 0.00 | 0.00 | 0.00 |
| | **1977** | 0.95 | 0.92 | 0.83 | 0.62 | 0.24 | 0.01 | 0.00 | 0.00 | 0.00 | 0.00 |
| | **1978** | 0.96 | 0.93 | 0.85 | 0.65 | 0.28 | 0.02 | 0.00 | 0.00 | 0.00 | 0.00 |
| | **1979** | 0.96 | 0.93 | 0.86 | 0.68 | 0.33 | 0.03 | 0.00 | 0.00 | 0.00 | 0.00 |
| | **1980** | 0.96 | 0.94 | 0.87 | 0.70 | 0.37 | 0.05 | 0.00 | 0.00 | 0.00 | 0.00 |
| | **1981** | 0.96 | 0.94 | 0.88 | 0.74 | 0.41 | 0.07 | 0.00 | 0.00 | 0.00 | 0.00 |
| | **1982** | 0.97 | 0.95 | 0.89 | 0.76 | 0.45 | 0.09 | 0.00 | 0.00 | 0.00 | 0.00 |
| | **1983** | 0.97 | 0.95 | 0.90 | 0.78 | 0.49 | 0.12 | 0.00 | 0.00 | 0.00 | 0.00 |
| | **1984** | 0.97 | 0.95 | 0.91 | 0.80 | 0.53 | 0.15 | 0.01 | 0.00 | 0.00 | 0.00 |
| | **1985** | 0.97 | 0.96 | 0.92 | 0.81 | 0.57 | 0.19 | 0.01 | 0.00 | 0.00 | 0.00 |
| | **1986** | 0.98 | 0.96 | 0.93 | 0.83 | 0.61 | 0.23 | 0.01 | 0.00 | 0.00 | 0.00 |
| | **1987** | 0.98 | 0.96 | 0.93 | 0.84 | 0.64 | 0.28 | 0.02 | 0.00 | 0.00 | 0.00 |
| | **1988** | 0.98 | 0.96 | 0.94 | 0.86 | 0.68 | 0.32 | 0.04 | 0.00 | 0.00 | 0.00 |
| | **1989** | 0.98 | 0.97 | 0.94 | 0.87 | 0.71 | 0.37 | 0.05 | 0.00 | 0.00 | 0.00 |
| | **1990** | 0.98 | 0.97 | 0.95 | 0.88 | 0.73 | 0.42 | 0.07 | 0.00 | 0.00 | 0.00 |
| | **1991** | 0.99 | 0.97 | 0.95 | 0.89 | 0.77 | 0.47 | 0.10 | 0.00 | 0.00 | 0.00 |

*Scotland's National Blood Transfusion Service (SNBTS) record-linkage study*

To assist the Infected Blood Inquiry, the SNBTS initiated a record-linkage study which followed up for mortality four 5-yearly cohorts of transfusion recipients, who joined their respective cohort at the date of their first transfusion in the cohort-year (8). The SNBTS database's coverage is not Scotland-wide but accounts for around 40% of Scotland's transfused patients. The cohort data were extracted from the SNBTS blood bank Laboratory Information Management System, eTraceline. The 1999 cohort, for example, represented three large teaching hospitals across Scotland and one large general hospital. The cohort-years are 1999 (1999-cohort); 2004 (2004-cohort); 2009 (2009-cohort); and 2014 (2014-cohort) to allow calendar-year trends in the deployment of units to, and survival outcome for, recipients of RBC transfusions to be monitored up to 20 years post-transfusion.

The SNBTS's suite of record-linkage studies was approved by Scotland's Public Benefit and Privacy Panel for Health and Social Care (PBPP-HSC) in 2021. Approval was necessary because individual consent for establishing survival status was not being sought. Minimal information about cohort-members includes age in completed years at 1st RBC-transfusion in their cohort-year, sex and International Classification of Diseases 2010 (ICD-10) disease-chapter for the underlying condition at hospital-discharge that aligned with the patient's RBC-transfusion-date.

The 1999-RBC-cohort comprised 13,260 persons with known sex (7,431 females; 5,829 males) and age-band (2,064 aged under 40 years (15.6%) of whom 1,286 (62%) were female; but 8,836 aged 60 years or older (66.6%) at their first RBC-transfusion in 2004, of whom 4,970 (56%) were female).

The 2004-RBC-cohort comprised 13,274 persons with known sex (7,438 females; 5,836 males) and age-band (1,565 aged under 40 years (11.8%) of whom 970 (62%) were female; but 9,520 aged 60 years or older (66.6%) at their first RBC-transfusion in 1999, of whom 5,345 (56%) were female).

Follow-up for mortality was to 31 December 2019. The vast majority of first transfusion recipients in any cohort-year has received RBC and so we focus on 1999-RBC-cohort, for whom we have 20 years of follow-up post-transfusion; and on the 2004-RBC-cohort, for whom we know survival status for 15 years post-transfusion.

Using the appropriate Scottish life-table for each year between 10 years post-transfusion and 2019 (e.g. 1999 – 2001 for the year 2000), annual hazards were extracted per sex and age-band at transfusion (using, for example, age 25 for those aged 20-29 years at transfusion), and updated to reflect the cohort's age in the year in question. These determined the expected number of deaths in the SNBTS cohorts. However, in this instance, there were only 5 relevant hazards contributing to the probability of surviving the epoch, one per year of the epoch under consideration. The expected number of survivors was estimated as the estimated number of people susceptible at the beginning of the epoch (e.g. alive at the end of the previous epoch or in the case of the first epoch, the number of people transfused) multiplied by the probability of surviving that epoch. The number of people susceptible at the beginning of each epoch was taken as the observed number of individuals surviving to that time minus the observed number of emigrations. The expected deaths were then the differences between the number of non-emigres who began the epoch and expected survivors. It is possible that some people who emigrated earlier in the follow-up then returned to Scotland to die - which was not accounted for here. However, this applied to a very small number of individuals and thus is unlikely to have any substantial effect on the resulting post-transfusion hazards.

The additional risks from 10 years post-transfusion were determined via the ratio of observed to expected deaths from the SNBTS's record-linkage study. Two scenarios were considered. First, hazards were pooled over age-bands and sex to give one estimated hazard ratio for 11 - 15 years post-transfusion and another for 16 - 20 years post-transfusion (with the exception of the 70 - 79 age-band which showed an anomalous reduced risk 16 - 20 years post-transfusion). In the case of 11 - 15 years post-transfusion, data were pooled over the 1999 and 2004 cohorts. This was not possible for the 16 - 20 years post-transfusion epoch due to lack of time elapsed since 2004, and so the hazard ratio for this epoch comes only from the 1999 cohort. The results are shown in Supplementary Table 27.

Second, the hazards were then stratified by age-band per epoch by pooling across sex and cohort years as applicable. Values for age-bands 0 and 1 - 9 years in the study were pooled as the 0 - 9 years band in accordance with our model structure. Similarly, the study's final age band is 80+, and so the hazards for this category were applied to both the 80 - 89 and 90+ age bands in the model. The results are shown in Supplementary Table 28.

The likelihood of surviving 10 years post-transfusion has already been covered in Task 4. Transfusion-adjusted hazard rates were obtained by multiplying the relevant hazards from the nation-specific life-tables for 11-15 years and 16-20 years post-transfusion by the observed/expected ratios calculated previously. The product of one minus the hazards across each year per age-band and sex was used to estimate the probability of survival to 2019, with and without the additional transfusion risk (Supplementary Figure 6).

These transfusion-adjusted hazard rates were applied to the output from Task 4 to give the estimated number of survivors to the end of 2019, both without and with allowance for the excess mortality of being post-transfusion.

The resulting survival probabilities can be seen Supplementary Table 29 - Supplementary Table 36, and the estimated number of infected individuals surviving to 2019 can be seen in Supplementary Table 37.

*Supplementary Table 27: Additional hazards from transfusion for 11 – 15 and 16 – 20 years post-transfusion by pooling across age band, sex and, where applicable, cohort year.*

| **Additional hazard 11 – 15 years post-transfusion** | **Additional hazard 16 – 20 years post-transfusion** |
|---|---|
| 1.72 | 1.32 |

*Supplementary Table 28: Additional hazards from transfusion by age band (in years) for 11 – 15 and 16 – 20 years post-transfusion by pooling across sex and, where applicable, cohort year.*

| **Age band at transfusion (years)** | **Additional hazard 11 – 15 years post-transfusion** | **Additional hazard 16 – 20 years post-transfusion** |
|---|---|---|
| 0 – 9 | 9.30 | 5.20 |
| 10 – 19 | 5.80 | 6.30 |
| 20 – 29 | 4.10 | 2.00 |
| 30 – 39 | 3.10 | 1.20 |
| 40 – 49 | 3.40 | 2.30 |
| 50 – 59 | 2.00 | 1.50 |
| 60 – 69 | 1.42 | 1.13 |
| 70 – 79 | 1.04 | 0.81 |
| 80 – 89 | 1.31 | 1.08 |
| 90+ | 1.31 | 1.08 |

*Supplementary Table 29: Probability of a female in England in each age-band and transfusion year surviving to the end of 2019 assuming excess age-stratified post-transfusion hazard beyond 10 years.*

| **Age band at transfusion (years)** | | **0 - 9** | **10 - 19** | **20 - 29** | **30 - 39** | **40 - 49** | **50 - 59** | **60 - 69** | **70 - 79** | **80 - 89** | **90+** |
|---|---|---|---|---|---|---|---|---|---|---|---|
| **Midpoint age (years)** | | 5 | 15 | 25 | 35 | 45 | 55 | 65 | 75 | 85 | 95 |
| **Midpoint age after surviving 10 years (years)** | | 15 | 25 | 35 | 45 | 55 | 65 | 75 | 85 | 95 | 105 |
| **Transfusion year** | **1970** | 0.95 | 0.89 | 0.78 | 0.50 | 0.09 | 0.00 | 0.00 | 0.00 | 0.00 | 0.00 |
| | **1971** | 0.95 | 0.90 | 0.80 | 0.54 | 0.12 | 0.00 | 0.00 | 0.00 | 0.00 | 0.00 |
| | **1972** | 0.95 | 0.91 | 0.82 | 0.58 | 0.14 | 0.00 | 0.00 | 0.00 | 0.00 | 0.00 |
| | **1973** | 0.95 | 0.91 | 0.83 | 0.61 | 0.18 | 0.00 | 0.00 | 0.00 | 0.00 | 0.00 |
| | **1974** | 0.95 | 0.92 | 0.84 | 0.64 | 0.22 | 0.01 | 0.00 | 0.00 | 0.00 | 0.00 |
| | **1975** | 0.96 | 0.92 | 0.85 | 0.67 | 0.26 | 0.02 | 0.00 | 0.00 | 0.00 | 0.00 |
| | **1976** | 0.96 | 0.93 | 0.86 | 0.70 | 0.30 | 0.03 | 0.00 | 0.00 | 0.00 | 0.00 |
| | **1977** | 0.96 | 0.93 | 0.87 | 0.72 | 0.34 | 0.04 | 0.00 | 0.00 | 0.00 | 0.00 |
| | **1978** | 0.96 | 0.94 | 0.88 | 0.75 | 0.38 | 0.06 | 0.00 | 0.00 | 0.00 | 0.00 |
| | **1979** | 0.96 | 0.94 | 0.89 | 0.77 | 0.43 | 0.08 | 0.00 | 0.00 | 0.00 | 0.00 |
| | **1980** | 0.97 | 0.94 | 0.90 | 0.79 | 0.47 | 0.10 | 0.00 | 0.00 | 0.00 | 0.00 |
| | **1981** | 0.97 | 0.95 | 0.91 | 0.81 | 0.51 | 0.13 | 0.00 | 0.00 | 0.00 | 0.00 |
| | **1982** | 0.97 | 0.95 | 0.91 | 0.82 | 0.54 | 0.16 | 0.00 | 0.00 | 0.00 | 0.00 |
| | **1983** | 0.97 | 0.95 | 0.92 | 0.84 | 0.57 | 0.20 | 0.00 | 0.00 | 0.00 | 0.00 |
| | **1984** | 0.97 | 0.95 | 0.93 | 0.85 | 0.60 | 0.24 | 0.02 | 0.00 | 0.00 | 0.00 |
| | **1985** | 0.97 | 0.96 | 0.93 | 0.86 | 0.63 | 0.28 | 0.02 | 0.00 | 0.00 | 0.00 |
| | **1986** | 0.97 | 0.96 | 0.94 | 0.87 | 0.66 | 0.33 | 0.04 | 0.00 | 0.00 | 0.00 |
| | **1987** | 0.97 | 0.96 | 0.94 | 0.88 | 0.68 | 0.38 | 0.05 | 0.00 | 0.00 | 0.00 |
| | **1988** | 0.98 | 0.96 | 0.95 | 0.89 | 0.71 | 0.42 | 0.07 | 0.00 | 0.00 | 0.00 |
| | **1989** | 0.98 | 0.96 | 0.95 | 0.90 | 0.73 | 0.47 | 0.10 | 0.00 | 0.00 | 0.00 |
| | **1990** | 0.98 | 0.97 | 0.95 | 0.90 | 0.75 | 0.51 | 0.13 | 0.00 | 0.00 | 0.00 |
| | **1991** | 0.98 | 0.97 | 0.96 | 0.91 | 0.77 | 0.55 | 0.16 | 0.00 | 0.00 | 0.00 |

*Supplementary Table 30: Probability of a male in England in each age-band and transfusion year surviving to the end of 2019 assuming excess age-stratified post-transfusion hazard beyond 10 years.*

| **Age band at transfusion (years)** | | **0 - 9** | **10 - 19** | **20 - 29** | **30 - 39** | **40 - 49** | **50 - 59** | **60 - 69** | **70 - 79** | **80 - 89** | **90+** |
|---|---|---|---|---|---|---|---|---|---|---|---|
| **Midpoint age (years)** | | 5 | 15 | 25 | 35 | 45 | 55 | 65 | 75 | 85 | 95 |
| **Midpoint age after surviving 10 years (years)** | | 15 | 25 | 35 | 45 | 55 | 65 | 75 | 85 | 95 | 105 |
| **Transfusion year** | **1970** | 0.89 | 0.83 | 0.69 | 0.36 | 0.03 | 0.00 | 0.00 | 0.00 | 0.00 | 0.00 |
| | **1971** | 0.89 | 0.84 | 0.72 | 0.40 | 0.05 | 0.00 | 0.00 | 0.00 | 0.00 | 0.00 |
| | **1972** | 0.90 | 0.85 | 0.74 | 0.44 | 0.07 | 0.00 | 0.00 | 0.00 | 0.00 | 0.00 |
| | **1973** | 0.90 | 0.86 | 0.76 | 0.47 | 0.09 | 0.00 | 0.00 | 0.00 | 0.00 | 0.00 |
| | **1974** | 0.90 | 0.87 | 0.77 | 0.51 | 0.11 | 0.00 | 0.00 | 0.00 | 0.00 | 0.00 |
| | **1975** | 0.91 | 0.87 | 0.79 | 0.54 | 0.14 | 0.01 | 0.00 | 0.00 | 0.00 | 0.00 |
| | **1976** | 0.91 | 0.88 | 0.80 | 0.57 | 0.17 | 0.01 | 0.00 | 0.00 | 0.00 | 0.00 |
| | **1977** | 0.91 | 0.89 | 0.81 | 0.61 | 0.20 | 0.01 | 0.00 | 0.00 | 0.00 | 0.00 |
| | **1978** | 0.92 | 0.89 | 0.82 | 0.64 | 0.24 | 0.02 | 0.00 | 0.00 | 0.00 | 0.00 |
| | **1979** | 0.92 | 0.89 | 0.84 | 0.67 | 0.28 | 0.03 | 0.00 | 0.00 | 0.00 | 0.00 |
| | **1980** | 0.92 | 0.90 | 0.85 | 0.69 | 0.31 | 0.04 | 0.00 | 0.00 | 0.00 | 0.00 |
| | **1981** | 0.93 | 0.90 | 0.86 | 0.72 | 0.35 | 0.06 | 0.00 | 0.00 | 0.00 | 0.00 |
| | **1982** | 0.93 | 0.90 | 0.87 | 0.75 | 0.39 | 0.08 | 0.00 | 0.00 | 0.00 | 0.00 |
| | **1983** | 0.93 | 0.91 | 0.88 | 0.76 | 0.42 | 0.10 | 0.00 | 0.00 | 0.00 | 0.00 |
| | **1984** | 0.94 | 0.91 | 0.89 | 0.78 | 0.45 | 0.13 | 0.01 | 0.00 | 0.00 | 0.00 |
| | **1985** | 0.94 | 0.92 | 0.90 | 0.79 | 0.48 | 0.17 | 0.01 | 0.00 | 0.00 | 0.00 |
| | **1986** | 0.94 | 0.92 | 0.90 | 0.81 | 0.52 | 0.20 | 0.01 | 0.00 | 0.00 | 0.00 |
| | **1987** | 0.94 | 0.92 | 0.91 | 0.82 | 0.55 | 0.24 | 0.02 | 0.00 | 0.00 | 0.00 |
| | **1988** | 0.94 | 0.92 | 0.92 | 0.83 | 0.58 | 0.28 | 0.03 | 0.00 | 0.00 | 0.00 |
| | **1989** | 0.95 | 0.93 | 0.92 | 0.85 | 0.61 | 0.33 | 0.05 | 0.00 | 0.00 | 0.00 |
| | **1990** | 0.95 | 0.93 | 0.93 | 0.86 | 0.63 | 0.37 | 0.07 | 0.00 | 0.00 | 0.00 |
| | **1991** | 0.95 | 0.94 | 0.93 | 0.87 | 0.66 | 0.41 | 0.09 | 0.00 | 0.00 | 0.00 |

*Supplementary Table 31: Probability of a female in Northern Ireland in each age-band and transfusion year surviving to the end of 2019 assuming excess age-stratified post-transfusion hazard beyond 10 years.*

| **Age band at transfusion (years)** | | **0 - 9** | **10 - 19** | **20 - 29** | **30 - 39** | **40 - 49** | **50 - 59** | **60 - 69** | **70 - 79** | **80 - 89** | **90+** |
|---|---|---|---|---|---|---|---|---|---|---|---|
| **Midpoint age (years)** | | 5 | 15 | 25 | 35 | 45 | 55 | 65 | 75 | 85 | 95 |
| **Midpoint age after surviving 10 years (years)** | | 15 | 25 | 35 | 45 | 55 | 65 | 75 | 85 | 95 | 105 |
| **Transfusion year** | **1970** | 0.95 | 0.88 | 0.77 | 0.48 | 0.08 | 0.00 | 0.00 | 0.00 | 0.00 | 0.00 |
| | **1971** | 0.95 | 0.89 | 0.79 | 0.52 | 0.10 | 0.00 | 0.00 | 0.00 | 0.00 | 0.00 |
| | **1972** | 0.95 | 0.90 | 0.80 | 0.55 | 0.13 | 0.00 | 0.00 | 0.00 | 0.00 | 0.00 |
| | **1973** | 0.95 | 0.90 | 0.82 | 0.59 | 0.16 | 0.00 | 0.00 | 0.00 | 0.00 | 0.00 |
| | **1974** | 0.95 | 0.91 | 0.83 | 0.62 | 0.20 | 0.01 | 0.00 | 0.00 | 0.00 | 0.00 |
| | **1975** | 0.96 | 0.92 | 0.84 | 0.65 | 0.23 | 0.02 | 0.00 | 0.00 | 0.00 | 0.00 |
| | **1976** | 0.96 | 0.93 | 0.86 | 0.68 | 0.27 | 0.02 | 0.00 | 0.00 | 0.00 | 0.00 |
| | **1977** | 0.96 | 0.93 | 0.87 | 0.71 | 0.32 | 0.03 | 0.00 | 0.00 | 0.00 | 0.00 |
| | **1978** | 0.97 | 0.93 | 0.88 | 0.73 | 0.36 | 0.05 | 0.00 | 0.00 | 0.00 | 0.00 |
| | **1979** | 0.97 | 0.94 | 0.88 | 0.75 | 0.40 | 0.07 | 0.00 | 0.00 | 0.00 | 0.00 |
| | **1980** | 0.97 | 0.95 | 0.89 | 0.77 | 0.44 | 0.09 | 0.00 | 0.00 | 0.00 | 0.00 |
| | **1981** | 0.97 | 0.95 | 0.90 | 0.79 | 0.48 | 0.12 | 0.00 | 0.00 | 0.00 | 0.00 |
| | **1982** | 0.97 | 0.95 | 0.91 | 0.81 | 0.52 | 0.15 | 0.00 | 0.00 | 0.00 | 0.00 |
| | **1983** | 0.97 | 0.95 | 0.91 | 0.82 | 0.55 | 0.18 | 0.00 | 0.00 | 0.00 | 0.00 |
| | **1984** | 0.97 | 0.96 | 0.92 | 0.84 | 0.58 | 0.22 | 0.01 | 0.00 | 0.00 | 0.00 |
| | **1985** | 0.97 | 0.96 | 0.93 | 0.85 | 0.61 | 0.27 | 0.02 | 0.00 | 0.00 | 0.00 |
| | **1986** | 0.97 | 0.96 | 0.93 | 0.86 | 0.64 | 0.31 | 0.03 | 0.00 | 0.00 | 0.00 |
| | **1987** | 0.97 | 0.96 | 0.94 | 0.87 | 0.67 | 0.35 | 0.04 | 0.00 | 0.00 | 0.00 |
| | **1988** | 0.97 | 0.96 | 0.94 | 0.88 | 0.68 | 0.40 | 0.06 | 0.00 | 0.00 | 0.00 |
| | **1989** | 0.98 | 0.96 | 0.95 | 0.89 | 0.71 | 0.45 | 0.09 | 0.00 | 0.00 | 0.00 |
| | **1990** | 0.98 | 0.97 | 0.95 | 0.89 | 0.73 | 0.49 | 0.11 | 0.00 | 0.00 | 0.00 |
| | **1991** | 0.98 | 0.97 | 0.96 | 0.90 | 0.75 | 0.53 | 0.15 | 0.00 | 0.00 | 0.00 |

*Supplementary Table 32: Probability of a male in Northern Ireland in each age-band and transfusion year surviving to the end of 2019 assuming excess age-stratified post-transfusion hazard beyond 10 years.*

| **Age band at transfusion (years)** | | **0 - 9** | **10 - 19** | **20 - 29** | **30 - 39** | **40 - 49** | **50 - 59** | **60 - 69** | **70 - 79** | **80 - 89** | **90+** |
|---|---|---|---|---|---|---|---|---|---|---|---|
| **Midpoint age (years)** | | 5 | 15 | 25 | 35 | 45 | 55 | 65 | 75 | 85 | 95 |
| **Midpoint age after surviving 10 years (years)** | | 15 | 25 | 35 | 45 | 55 | 65 | 75 | 85 | 95 | 105 |
| **Transfusion year** | **1970** | 0.86 | 0.81 | 0.66 | 0.32 | 0.03 | 0.00 | 0.00 | 0.00 | 0.00 | 0.00 |
| | **1971** | 0.87 | 0.82 | 0.69 | 0.36 | 0.04 | 0.00 | 0.00 | 0.00 | 0.00 | 0.00 |
| | **1972** | 0.87 | 0.84 | 0.71 | 0.40 | 0.05 | 0.00 | 0.00 | 0.00 | 0.00 | 0.00 |
| | **1973** | 0.88 | 0.84 | 0.74 | 0.45 | 0.07 | 0.00 | 0.00 | 0.00 | 0.00 | 0.00 |
| | **1974** | 0.88 | 0.85 | 0.76 | 0.48 | 0.09 | 0.00 | 0.00 | 0.00 | 0.00 | 0.00 |
| | **1975** | 0.89 | 0.86 | 0.78 | 0.52 | 0.11 | 0.00 | 0.00 | 0.00 | 0.00 | 0.00 |
| | **1976** | 0.89 | 0.86 | 0.79 | 0.55 | 0.14 | 0.01 | 0.00 | 0.00 | 0.00 | 0.00 |
| | **1977** | 0.90 | 0.87 | 0.80 | 0.58 | 0.18 | 0.01 | 0.00 | 0.00 | 0.00 | 0.00 |
| | **1978** | 0.90 | 0.88 | 0.82 | 0.61 | 0.21 | 0.02 | 0.00 | 0.00 | 0.00 | 0.00 |
| | **1979** | 0.90 | 0.88 | 0.83 | 0.64 | 0.24 | 0.02 | 0.00 | 0.00 | 0.00 | 0.00 |
| | **1980** | 0.91 | 0.89 | 0.84 | 0.67 | 0.28 | 0.03 | 0.00 | 0.00 | 0.00 | 0.00 |
| | **1981** | 0.91 | 0.90 | 0.86 | 0.70 | 0.32 | 0.05 | 0.00 | 0.00 | 0.00 | 0.00 |
| | **1982** | 0.91 | 0.90 | 0.87 | 0.72 | 0.36 | 0.06 | 0.00 | 0.00 | 0.00 | 0.00 |
| | **1983** | 0.91 | 0.90 | 0.88 | 0.74 | 0.40 | 0.09 | 0.00 | 0.00 | 0.00 | 0.00 |
| | **1984** | 0.91 | 0.91 | 0.88 | 0.76 | 0.43 | 0.11 | 0.00 | 0.00 | 0.00 | 0.00 |
| | **1985** | 0.92 | 0.92 | 0.89 | 0.79 | 0.46 | 0.14 | 0.01 | 0.00 | 0.00 | 0.00 |
| | **1986** | 0.92 | 0.92 | 0.90 | 0.80 | 0.49 | 0.18 | 0.01 | 0.00 | 0.00 | 0.00 |
| | **1987** | 0.92 | 0.92 | 0.90 | 0.81 | 0.52 | 0.22 | 0.02 | 0.00 | 0.00 | 0.00 |
| | **1988** | 0.92 | 0.92 | 0.91 | 0.83 | 0.55 | 0.26 | 0.03 | 0.00 | 0.00 | 0.00 |
| | **1989** | 0.92 | 0.92 | 0.91 | 0.84 | 0.58 | 0.30 | 0.04 | 0.00 | 0.00 | 0.00 |
| | **1990** | 0.93 | 0.92 | 0.92 | 0.85 | 0.61 | 0.34 | 0.05 | 0.00 | 0.00 | 0.00 |
| | **1991** | 0.93 | 0.92 | 0.92 | 0.86 | 0.64 | 0.38 | 0.07 | 0.00 | 0.00 | 0.00 |

*Supplementary Table 33: Probability of a female in Scotland in each age-band and transfusion year surviving to the end of 2019 assuming excess age-stratified post-transfusion hazard beyond 10 years.*

| **Age band at transfusion (years)** | | **0 - 9** | **10 - 19** | **20 - 29** | **30 - 39** | **40 - 49** | **50 - 59** | **60 - 69** | **70 - 79** | **80 - 89** | **90+** |
|---|---|---|---|---|---|---|---|---|---|---|---|
| **Midpoint age (years)** | | 5 | 15 | 25 | 35 | 45 | 55 | 65 | 75 | 85 | 95 |
| **Midpoint age after surviving 10 years (years)** | | 15 | 25 | 35 | 45 | 55 | 65 | 75 | 85 | 95 | 105 |
| **Transfusion year** | **1970** | 0.93 | 0.87 | 0.73 | 0.42 | 0.06 | 0.00 | 0.00 | 0.00 | 0.00 | 0.00 |
| | **1971** | 0.94 | 0.88 | 0.75 | 0.46 | 0.08 | 0.00 | 0.00 | 0.00 | 0.00 | 0.00 |
| | **1972** | 0.94 | 0.88 | 0.77 | 0.50 | 0.10 | 0.00 | 0.00 | 0.00 | 0.00 | 0.00 |
| | **1973** | 0.94 | 0.89 | 0.79 | 0.53 | 0.13 | 0.00 | 0.00 | 0.00 | 0.00 | 0.00 |
| | **1974** | 0.95 | 0.90 | 0.81 | 0.57 | 0.16 | 0.01 | 0.00 | 0.00 | 0.00 | 0.00 |
| | **1975** | 0.95 | 0.91 | 0.82 | 0.60 | 0.19 | 0.01 | 0.00 | 0.00 | 0.00 | 0.00 |
| | **1976** | 0.95 | 0.91 | 0.83 | 0.63 | 0.23 | 0.02 | 0.00 | 0.00 | 0.00 | 0.00 |
| | **1977** | 0.95 | 0.91 | 0.84 | 0.65 | 0.27 | 0.03 | 0.00 | 0.00 | 0.00 | 0.00 |
| | **1978** | 0.95 | 0.92 | 0.85 | 0.68 | 0.31 | 0.04 | 0.00 | 0.00 | 0.00 | 0.00 |
| | **1979** | 0.96 | 0.92 | 0.87 | 0.71 | 0.35 | 0.05 | 0.00 | 0.00 | 0.00 | 0.00 |
| | **1980** | 0.96 | 0.93 | 0.88 | 0.73 | 0.39 | 0.07 | 0.00 | 0.00 | 0.00 | 0.00 |
| | **1981** | 0.96 | 0.93 | 0.88 | 0.76 | 0.42 | 0.09 | 0.00 | 0.00 | 0.00 | 0.00 |
| | **1982** | 0.96 | 0.94 | 0.89 | 0.78 | 0.46 | 0.12 | 0.00 | 0.00 | 0.00 | 0.00 |
| | **1983** | 0.96 | 0.94 | 0.90 | 0.80 | 0.49 | 0.15 | 0.00 | 0.00 | 0.00 | 0.00 |
| | **1984** | 0.96 | 0.94 | 0.91 | 0.81 | 0.52 | 0.18 | 0.01 | 0.00 | 0.00 | 0.00 |
| | **1985** | 0.96 | 0.95 | 0.91 | 0.82 | 0.55 | 0.22 | 0.02 | 0.00 | 0.00 | 0.00 |
| | **1986** | 0.96 | 0.95 | 0.92 | 0.84 | 0.57 | 0.26 | 0.03 | 0.00 | 0.00 | 0.00 |
| | **1987** | 0.96 | 0.95 | 0.93 | 0.85 | 0.60 | 0.31 | 0.04 | 0.00 | 0.00 | 0.00 |
| | **1988** | 0.97 | 0.95 | 0.93 | 0.86 | 0.63 | 0.35 | 0.05 | 0.00 | 0.00 | 0.00 |
| | **1989** | 0.97 | 0.95 | 0.94 | 0.87 | 0.66 | 0.40 | 0.07 | 0.00 | 0.00 | 0.00 |
| | **1990** | 0.97 | 0.95 | 0.94 | 0.88 | 0.68 | 0.44 | 0.10 | 0.00 | 0.00 | 0.00 |
| | **1991** | 0.97 | 0.95 | 0.94 | 0.89 | 0.71 | 0.48 | 0.13 | 0.00 | 0.00 | 0.00 |

*Supplementary Table 34: Probability of a male in Scotland in each age-band and transfusion year surviving to the end of 2019 assuming excess age-stratified post-transfusion hazard beyond 10 years.*

| **Age band at transfusion (years)** | | **0 - 9** | **10 - 19** | **20 - 29** | **30 - 39** | **40 - 49** | **50 - 59** | **60 - 69** | **70 - 79** | **80 - 89** | **90+** |
|---|---|---|---|---|---|---|---|---|---|---|---|
| **Midpoint age (years)** | | 5 | 15 | 25 | 35 | 45 | 55 | 65 | 75 | 85 | 95 |
| **Midpoint age after surviving 10 years (years)** | | 15 | 25 | 35 | 45 | 55 | 65 | 75 | 85 | 95 | 105 |
| **Transfusion year** | **1970** | 0.86 | 0.79 | 0.61 | 0.28 | 0.02 | 0.00 | 0.00 | 0.00 | 0.00 | 0.00 |
| | **1971** | 0.87 | 0.80 | 0.65 | 0.31 | 0.03 | 0.00 | 0.00 | 0.00 | 0.00 | 0.00 |
| | **1972** | 0.87 | 0.81 | 0.67 | 0.35 | 0.04 | 0.00 | 0.00 | 0.00 | 0.00 | 0.00 |
| | **1973** | 0.87 | 0.82 | 0.70 | 0.39 | 0.06 | 0.00 | 0.00 | 0.00 | 0.00 | 0.00 |
| | **1974** | 0.87 | 0.83 | 0.71 | 0.42 | 0.07 | 0.00 | 0.00 | 0.00 | 0.00 | 0.00 |
| | **1975** | 0.88 | 0.84 | 0.73 | 0.45 | 0.09 | 0.00 | 0.00 | 0.00 | 0.00 | 0.00 |
| | **1976** | 0.88 | 0.85 | 0.75 | 0.48 | 0.12 | 0.01 | 0.00 | 0.00 | 0.00 | 0.00 |
| | **1977** | 0.89 | 0.85 | 0.77 | 0.52 | 0.15 | 0.01 | 0.00 | 0.00 | 0.00 | 0.00 |
| | **1978** | 0.89 | 0.85 | 0.79 | 0.55 | 0.17 | 0.01 | 0.00 | 0.00 | 0.00 | 0.00 |
| | **1979** | 0.89 | 0.86 | 0.80 | 0.59 | 0.21 | 0.02 | 0.00 | 0.00 | 0.00 | 0.00 |
| | **1980** | 0.89 | 0.86 | 0.81 | 0.62 | 0.24 | 0.03 | 0.00 | 0.00 | 0.00 | 0.00 |
| | **1981** | 0.89 | 0.86 | 0.82 | 0.66 | 0.27 | 0.04 | 0.00 | 0.00 | 0.00 | 0.00 |
| | **1982** | 0.89 | 0.87 | 0.84 | 0.68 | 0.30 | 0.05 | 0.00 | 0.00 | 0.00 | 0.00 |
| | **1983** | 0.90 | 0.87 | 0.85 | 0.71 | 0.33 | 0.07 | 0.00 | 0.00 | 0.00 | 0.00 |
| | **1984** | 0.90 | 0.87 | 0.86 | 0.72 | 0.36 | 0.09 | 0.00 | 0.00 | 0.00 | 0.00 |
| | **1985** | 0.91 | 0.87 | 0.86 | 0.74 | 0.39 | 0.12 | 0.01 | 0.00 | 0.00 | 0.00 |
| | **1986** | 0.91 | 0.88 | 0.87 | 0.76 | 0.42 | 0.15 | 0.01 | 0.00 | 0.00 | 0.00 |
| | **1987** | 0.91 | 0.88 | 0.88 | 0.78 | 0.45 | 0.19 | 0.02 | 0.00 | 0.00 | 0.00 |
| | **1988** | 0.92 | 0.88 | 0.88 | 0.79 | 0.48 | 0.22 | 0.02 | 0.00 | 0.00 | 0.00 |
| | **1989** | 0.92 | 0.88 | 0.89 | 0.81 | 0.52 | 0.26 | 0.04 | 0.00 | 0.00 | 0.00 |
| | **1990** | 0.92 | 0.88 | 0.89 | 0.82 | 0.55 | 0.30 | 0.05 | 0.00 | 0.00 | 0.00 |
| | **1991** | 0.93 | 0.89 | 0.90 | 0.83 | 0.59 | 0.33 | 0.07 | 0.00 | 0.00 | 0.00 |

*Supplementary Table 35: Probability of a female in Wales in each age-band and transfusion year surviving to the end of 2019 assuming excess age-stratified post-transfusion hazard beyond 10 years.*

| **Age band at transfusion (years)** | | **0 - 9** | **10 - 19** | **20 - 29** | **30 - 39** | **40 - 49** | **50 - 59** | **60 - 69** | **70 - 79** | **80 - 89** | **90+** |
|---|---|---|---|---|---|---|---|---|---|---|---|
| **Midpoint age (years)** | | 5 | 15 | 25 | 35 | 45 | 55 | 65 | 75 | 85 | 95 |
| **Midpoint age after surviving 10 years (years)** | | 15 | 25 | 35 | 45 | 55 | 65 | 75 | 85 | 95 | 105 |
| **Transfusion year** | **1970** | 0.94 | 0.88 | 0.76 | 0.48 | 0.08 | 0.00 | 0.00 | 0.00 | 0.00 | 0.00 |
| | **1971** | 0.95 | 0.89 | 0.78 | 0.51 | 0.10 | 0.00 | 0.00 | 0.00 | 0.00 | 0.00 |
| | **1972** | 0.95 | 0.90 | 0.80 | 0.55 | 0.13 | 0.00 | 0.00 | 0.00 | 0.00 | 0.00 |
| | **1973** | 0.95 | 0.91 | 0.82 | 0.59 | 0.16 | 0.00 | 0.00 | 0.00 | 0.00 | 0.00 |
| | **1974** | 0.95 | 0.91 | 0.83 | 0.62 | 0.20 | 0.01 | 0.00 | 0.00 | 0.00 | 0.00 |
| | **1975** | 0.95 | 0.92 | 0.84 | 0.65 | 0.23 | 0.02 | 0.00 | 0.00 | 0.00 | 0.00 |
| | **1976** | 0.96 | 0.92 | 0.86 | 0.68 | 0.28 | 0.02 | 0.00 | 0.00 | 0.00 | 0.00 |
| | **1977** | 0.96 | 0.93 | 0.87 | 0.71 | 0.32 | 0.03 | 0.00 | 0.00 | 0.00 | 0.00 |
| | **1978** | 0.96 | 0.93 | 0.87 | 0.73 | 0.36 | 0.05 | 0.00 | 0.00 | 0.00 | 0.00 |
| | **1979** | 0.96 | 0.93 | 0.88 | 0.75 | 0.40 | 0.07 | 0.00 | 0.00 | 0.00 | 0.00 |
| | **1980** | 0.96 | 0.94 | 0.89 | 0.77 | 0.44 | 0.09 | 0.00 | 0.00 | 0.00 | 0.00 |
| | **1981** | 0.96 | 0.94 | 0.89 | 0.79 | 0.48 | 0.12 | 0.00 | 0.00 | 0.00 | 0.00 |
| | **1982** | 0.96 | 0.95 | 0.90 | 0.81 | 0.51 | 0.15 | 0.00 | 0.00 | 0.00 | 0.00 |
| | **1983** | 0.97 | 0.95 | 0.91 | 0.82 | 0.55 | 0.18 | 0.00 | 0.00 | 0.00 | 0.00 |
| | **1984** | 0.97 | 0.95 | 0.92 | 0.83 | 0.58 | 0.22 | 0.01 | 0.00 | 0.00 | 0.00 |
| | **1985** | 0.97 | 0.95 | 0.92 | 0.85 | 0.60 | 0.26 | 0.02 | 0.00 | 0.00 | 0.00 |
| | **1986** | 0.97 | 0.95 | 0.93 | 0.86 | 0.63 | 0.31 | 0.03 | 0.00 | 0.00 | 0.00 |
| | **1987** | 0.97 | 0.96 | 0.93 | 0.87 | 0.66 | 0.35 | 0.04 | 0.00 | 0.00 | 0.00 |
| | **1988** | 0.98 | 0.96 | 0.94 | 0.88 | 0.68 | 0.40 | 0.06 | 0.00 | 0.00 | 0.00 |
| | **1989** | 0.98 | 0.96 | 0.94 | 0.89 | 0.70 | 0.44 | 0.09 | 0.00 | 0.00 | 0.00 |
| | **1990** | 0.98 | 0.96 | 0.95 | 0.89 | 0.72 | 0.48 | 0.12 | 0.00 | 0.00 | 0.00 |
| | **1991** | 0.98 | 0.96 | 0.95 | 0.90 | 0.74 | 0.53 | 0.15 | 0.00 | 0.00 | 0.00 |

*Supplementary Table 36: Probability of a male in Wales in each age-band and transfusion year surviving to the end of 2019 assuming excess age-stratified post-transfusion hazard beyond 10 years.*

| **Age band at transfusion (years)** | | **0 - 9** | **10 - 19** | **20 - 29** | **30 - 39** | **40 - 49** | **50 - 59** | **60 - 69** | **70 - 79** | **80 - 89** | **90+** |
|---|---|---|---|---|---|---|---|---|---|---|---|
| **Midpoint age (years)** | | 5 | 15 | 25 | 35 | 45 | 55 | 65 | 75 | 85 | 95 |
| **Midpoint age after surviving 10 years (years)** | | 15 | 25 | 35 | 45 | 55 | 65 | 75 | 85 | 95 | 105 |
| **Transfusion year** | **1970** | 0.88 | 0.82 | 0.67 | 0.33 | 0.03 | 0.00 | 0.00 | 0.00 | 0.00 | 0.00 |
| | **1971** | 0.88 | 0.83 | 0.70 | 0.37 | 0.04 | 0.00 | 0.00 | 0.00 | 0.00 | 0.00 |
| | **1972** | 0.89 | 0.84 | 0.72 | 0.41 | 0.05 | 0.00 | 0.00 | 0.00 | 0.00 | 0.00 |
| | **1973** | 0.89 | 0.86 | 0.74 | 0.44 | 0.07 | 0.00 | 0.00 | 0.00 | 0.00 | 0.00 |
| | **1974** | 0.90 | 0.86 | 0.76 | 0.48 | 0.10 | 0.00 | 0.00 | 0.00 | 0.00 | 0.00 |
| | **1975** | 0.90 | 0.87 | 0.77 | 0.52 | 0.12 | 0.00 | 0.00 | 0.00 | 0.00 | 0.00 |
| | **1976** | 0.90 | 0.87 | 0.79 | 0.55 | 0.15 | 0.01 | 0.00 | 0.00 | 0.00 | 0.00 |
| | **1977** | 0.90 | 0.88 | 0.80 | 0.59 | 0.18 | 0.01 | 0.00 | 0.00 | 0.00 | 0.00 |
| | **1978** | 0.90 | 0.88 | 0.82 | 0.62 | 0.22 | 0.02 | 0.00 | 0.00 | 0.00 | 0.00 |
| | **1979** | 0.90 | 0.88 | 0.83 | 0.65 | 0.25 | 0.02 | 0.00 | 0.00 | 0.00 | 0.00 |
| | **1980** | 0.90 | 0.88 | 0.84 | 0.67 | 0.29 | 0.03 | 0.00 | 0.00 | 0.00 | 0.00 |
| | **1981** | 0.91 | 0.89 | 0.85 | 0.70 | 0.32 | 0.05 | 0.00 | 0.00 | 0.00 | 0.00 |
| | **1982** | 0.91 | 0.89 | 0.86 | 0.73 | 0.36 | 0.07 | 0.00 | 0.00 | 0.00 | 0.00 |
| | **1983** | 0.91 | 0.89 | 0.87 | 0.75 | 0.39 | 0.09 | 0.00 | 0.00 | 0.00 | 0.00 |
| | **1984** | 0.91 | 0.90 | 0.88 | 0.77 | 0.43 | 0.12 | 0.00 | 0.00 | 0.00 | 0.00 |
| | **1985** | 0.92 | 0.90 | 0.89 | 0.78 | 0.46 | 0.15 | 0.01 | 0.00 | 0.00 | 0.00 |
| | **1986** | 0.93 | 0.90 | 0.89 | 0.79 | 0.49 | 0.18 | 0.01 | 0.00 | 0.00 | 0.00 |
| | **1987** | 0.93 | 0.91 | 0.90 | 0.81 | 0.53 | 0.22 | 0.02 | 0.00 | 0.00 | 0.00 |
| | **1988** | 0.93 | 0.90 | 0.91 | 0.82 | 0.56 | 0.26 | 0.03 | 0.00 | 0.00 | 0.00 |
| | **1989** | 0.94 | 0.91 | 0.91 | 0.84 | 0.59 | 0.30 | 0.04 | 0.00 | 0.00 | 0.00 |
| | **1990** | 0.94 | 0.91 | 0.92 | 0.85 | 0.61 | 0.34 | 0.06 | 0.00 | 0.00 | 0.00 |
| | **1991** | 0.94 | 0.91 | 0.92 | 0.86 | 0.64 | 0.38 | 0.08 | 0.00 | 0.00 | 0.00 |

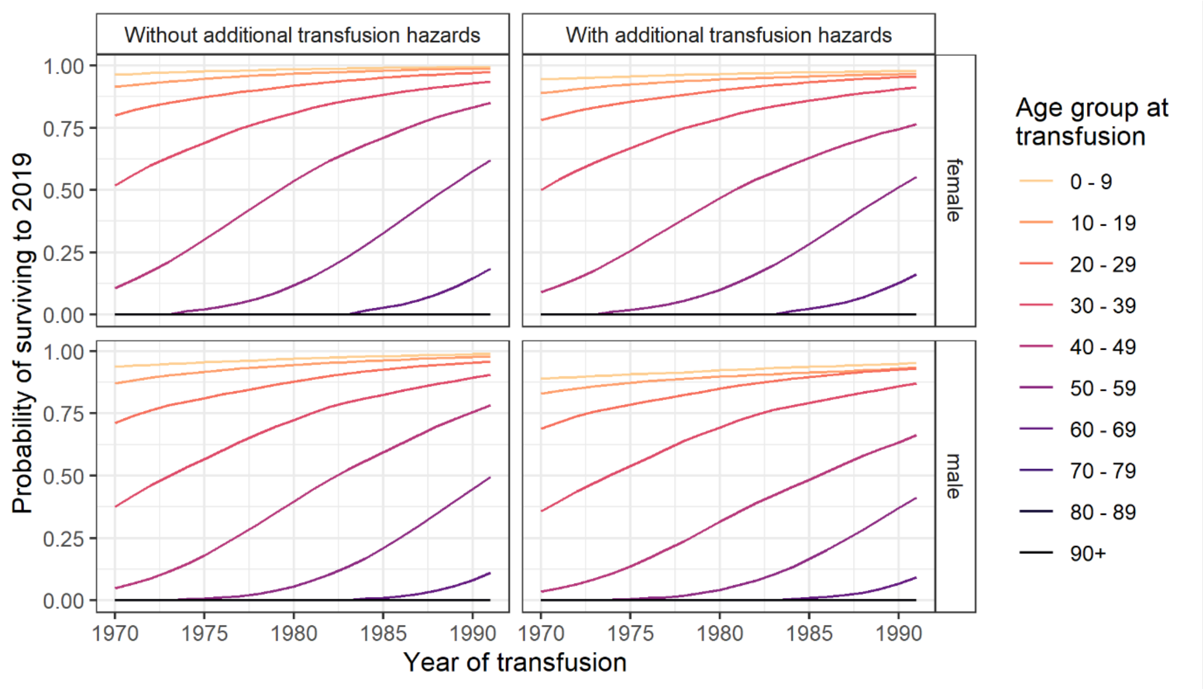

*Supplementary Figure 6: Estimated probability of surviving until 31 December 2019 post-transfusion in England, given age band (in years), sex and year of transfusion and assuming excess age-stratified post-transfusion hazard.*

*Supplementary Table 37: Estimated age-sex distribution of numbers who were chronically HCV-infected surviving to the end of 2019 in each of the four nations, up until 2019. Estimates are provided as median and 95% uncertainty intervals.*

| **Age at transfusion (in completed years)** | **Proportion** | | **Number surviving to the end of 2019** | | | | | | | |
|---|---|---|---|---|---|---|---|---|---|---|
| | | | **England** | | **Northern Ireland** | | **Scotland** | | **Wales** | |
| | **Females** | **Males** | **Females** | **Males** | **Females** | **Males** | **Females** | **Males** | **Females** | **Males** |
| 0 – 9 | 0.016 (0.012 - 0.021) | 0.027 (0.021 - 0.033) | 190 (120 - 300) | 350 (240 - 540) | 6 (2 - 14) | 13 (6 - 23) | 24 (12 - 40) | 44 (28 - 64) | 12 (5 - 22) | 23 (12 - 38) |
| 10 – 19 | 0.0092 (0.0062 - 0.013) | 0.0082 (0.0054 - 0.012) | 120 (75 - 220) | 72 (37 - 130) | 4 (1 - 11) | 3 (0 - 7) | 17 (7 - 30) | 10 (3 - 19) | 8 (2 - 17) | 5 (1 - 12) |
| 20 – 29 | 0.037 (0.030 - 0.044) | 0.012 (0.0088 - 0.017) | 550 (390 - 830) | 140 (86 - 230) | 20 (11 - 33) | 6 (1 - 12) | 68 (47 - 96) | 19 (10 - 30) | 35 (22 - 55) | 10 (4 - 18) |
| 30 – 39 | 0.034 (0.028 - 0.041) | 0.019 (0.014 - 0.024) | 330 (230 - 510) | 140 (89 - 230) | 12 (6 - 20) | 5 (1 - 10) | 40 (25 - 60) | 16 (9 - 28) | 20 (12 - 33) | 9 (3 - 16) |
| 40 – 49 | 0.045 (0.037 - 0.052) | 0.030 (0.024 - 0.036) | 160 (100 - 250) | 70 (41 - 120) | 5 (1 - 10) | 1 (0 - 4) | 18 (10 - 30) | 7 (2 - 14) | 8 (3 - 16) | 2 (0 - 7) |
| 50 – 59 | 0.053 (0.045 - 0.061) | 0.052 (0.044- 0.061) | 47 (26 - 80) | 20 (9 - 39) | 0 (0 - 2) | 0 (0 - 1) | 5 (2 - 11) | 2 (0 - 5) | 1 (0 - 4) | 0 (0 - 2) |
| 60 – 69 | 0.089 (0.079 - 0.100) | 0.110 (0.100 - 0.130) | 3 (0 - 11) | 1 (0 - 5) | 0 (0 - 0) | 0 (0 - 0) | 0 (0 - 1) | 0 (0 - 0) | 0 (0 - 0) | 0 (0 - 0) |
| 70 – 79 | 0.140 (0.130 - 0.160) | 0.120 (0.110 - 0.130) | 0 (0 - 0) | 0 (0 - 0) | 0 (0 - 0) | 0 (0 - 0) | 0 (0 - 0) | 0 (0 - 0) | 0 (0 - 0) | 0 (0 - 0) |
| 80 – 89 | 0.110 (0.097 - 0.12) | 0.055 (0.047 - 0.064) | 0 (0 - 0) | 0 (0 - 0 | 0 (0 - 0) | 0 (0 - 0) | 0 (0 - 0) | 0 (0 - 0) | 0 (0 - 0) | 0 (0 - 0) |
| 90+ | 0.020 (0.015 - 0.025) | 0.0044 (0.0024 - 0.0072) | 0 (0 - 0) | 0 (0 - 0) | 0 (0 - 0) | 0 (0 - 0) | 0 (0 - 0) | 0 (0 - 0) | 0 (0 - 0) | 0 (0 - 0) |
| Total | 0.56 (0.54 - 0.57) | 0.44 (0.43 - 0.46) | **1,400** (1,100 - 2,100 | **800** (590 - 1,200) | **49** (32 - 74) | **28** (16 - 44) | **170** (130 - 230) | **99** (70 - 130) | **86** (60 - 130) | **50** (32 - 76) |

### Task 6: Of those chronically HCV-infected through transfusion between January 1970 and August 1991, how many died of HCV-related causes by the end of 2019?

Supplementary Table 38 to Supplementary Table 45 present the transfusion- and chronic HCV-adjusted probabilities of surviving to the end of 2019.

Supplementary Figure 7 presents the estimated number of chronically HCV-infected people surviving to each year up to the end of 2019 under the three different survival scenarios: without any additional hazards; with age-stratified transfusion hazards; and with age-stratified transfusion hazards and additional hazard from having chronic HCV.

Supplementary Table 46 to Supplementary Table 49 present the number of survivors to the end of 2019 under the three aforementioned survival scenarios stratified by year of transfusion.

Supplementary Table 50 presents the estimated number of people alive in 2019 according to their age in 2019 under both age-stratified transfusion hazards and additional hazard from having chronic HCV in each of the four nations.

*Supplementary Table 38: Probability of a female in England in each age-band and transfusion year surviving to the end of 2019 assuming excess chronic HCV infection and excess post-transfusion hazard beyond 10 years.*

| **Age band at transfusion (years)** | | **0 - 9** | **10 - 19** | **20 - 29** | **30 - 39** | **40 - 49** | **50 - 59** | **60 - 69** | **70 - 79** | **80 - 89** | **90+** |
|---|---|---|---|---|---|---|---|---|---|---|---|
| **Midpoint age (years)** | | 5 | 15 | 25 | 35 | 45 | 55 | 65 | 75 | 85 | 95 |
| **Midpoint age after surviving 10 years (years)** | | 15 | 25 | 35 | 45 | 55 | 65 | 75 | 85 | 95 | 105 |
| **Transfusion year** | **1970** | 0.92 | 0.84 | 0.68 | 0.34 | 0.02 | 0.00 | 0.00 | 0.00 | 0.00 | 0.00 |
| | **1971** | 0.92 | 0.85 | 0.71 | 0.39 | 0.03 | 0.00 | 0.00 | 0.00 | 0.00 | 0.00 |
| | **1972** | 0.92 | 0.86 | 0.73 | 0.43 | 0.05 | 0.00 | 0.00 | 0.00 | 0.00 | 0.00 |
| | **1973** | 0.93 | 0.87 | 0.76 | 0.47 | 0.07 | 0.00 | 0.00 | 0.00 | 0.00 | 0.00 |
| | **1974** | 0.93 | 0.88 | 0.77 | 0.51 | 0.09 | 0.00 | 0.00 | 0.00 | 0.00 | 0.00 |
| | **1975** | 0.94 | 0.89 | 0.79 | 0.54 | 0.12 | 0.00 | 0.00 | 0.00 | 0.00 | 0.00 |
| | **1976** | 0.94 | 0.89 | 0.80 | 0.57 | 0.15 | 0.00 | 0.00 | 0.00 | 0.00 | 0.00 |
| | **1977** | 0.94 | 0.90 | 0.81 | 0.61 | 0.19 | 0.01 | 0.00 | 0.00 | 0.00 | 0.00 |
| | **1978** | 0.94 | 0.91 | 0.83 | 0.64 | 0.23 | 0.01 | 0.00 | 0.00 | 0.00 | 0.00 |
| | **1979** | 0.95 | 0.91 | 0.84 | 0.67 | 0.27 | 0.02 | 0.00 | 0.00 | 0.00 | 0.00 |
| | **1980** | 0.95 | 0.92 | 0.85 | 0.69 | 0.31 | 0.03 | 0.00 | 0.00 | 0.00 | 0.00 |
| | **1981** | 0.95 | 0.92 | 0.86 | 0.72 | 0.35 | 0.04 | 0.00 | 0.00 | 0.00 | 0.00 |
| | **1982** | 0.95 | 0.92 | 0.87 | 0.74 | 0.39 | 0.06 | 0.00 | 0.00 | 0.00 | 0.00 |
| | **1983** | 0.95 | 0.93 | 0.88 | 0.76 | 0.42 | 0.08 | 0.00 | 0.00 | 0.00 | 0.00 |
| | **1984** | 0.96 | 0.93 | 0.89 | 0.78 | 0.46 | 0.11 | 0.00 | 0.00 | 0.00 | 0.00 |
| | **1985** | 0.96 | 0.93 | 0.90 | 0.79 | 0.49 | 0.14 | 0.00 | 0.00 | 0.00 | 0.00 |
| | **1986** | 0.96 | 0.94 | 0.91 | 0.81 | 0.52 | 0.18 | 0.00 | 0.00 | 0.00 | 0.00 |
| | **1987** | 0.96 | 0.94 | 0.91 | 0.82 | 0.56 | 0.22 | 0.01 | 0.00 | 0.00 | 0.00 |
| | **1988** | 0.96 | 0.94 | 0.92 | 0.83 | 0.59 | 0.26 | 0.01 | 0.00 | 0.00 | 0.00 |
| | **1989** | 0.96 | 0.95 | 0.92 | 0.85 | 0.61 | 0.31 | 0.02 | 0.00 | 0.00 | 0.00 |
| | **1990** | 0.97 | 0.95 | 0.93 | 0.86 | 0.64 | 0.36 | 0.04 | 0.00 | 0.00 | 0.00 |
| | **1991** | 0.97 | 0.95 | 0.93 | 0.87 | 0.66 | 0.40 | 0.06 | 0.00 | 0.00 | 0.00 |

*Supplementary Table 39: Probability of a male in England in each age-band and transfusion year surviving to the end of 2019 assuming excess chronic HCV infection and excess post-transfusion hazard beyond 10 years.*

| **Age band at transfusion (years)** | | **0 - 9** | **10 - 19** | **20 - 29** | **30 - 39** | **40 - 49** | **50 - 59** | **60 - 69** | **70 - 79** | **80 - 89** | **90+** |
|---|---|---|---|---|---|---|---|---|---|---|---|
| **Midpoint age (years)** | | 5 | 15 | 25 | 35 | 45 | 55 | 65 | 75 | 85 | 95 |
| **Midpoint age after surviving 10 years (years)** | | 15 | 25 | 35 | 45 | 55 | 65 | 75 | 85 | 95 | 105 |
| **Transfusion year** | **1970** | 0.83 | 0.75 | 0.56 | 0.20 | 0.00 | 0.00 | 0.00 | 0.00 | 0.00 | 0.00 |
| | **1971** | 0.84 | 0.77 | 0.60 | 0.24 | 0.01 | 0.00 | 0.00 | 0.00 | 0.00 | 0.00 |
| | **1972** | 0.85 | 0.78 | 0.63 | 0.28 | 0.01 | 0.00 | 0.00 | 0.00 | 0.00 | 0.00 |
| | **1973** | 0.85 | 0.79 | 0.65 | 0.32 | 0.02 | 0.00 | 0.00 | 0.00 | 0.00 | 0.00 |
| | **1974** | 0.86 | 0.80 | 0.67 | 0.35 | 0.03 | 0.00 | 0.00 | 0.00 | 0.00 | 0.00 |
| | **1975** | 0.86 | 0.81 | 0.69 | 0.39 | 0.04 | 0.00 | 0.00 | 0.00 | 0.00 | 0.00 |
| | **1976** | 0.87 | 0.82 | 0.71 | 0.42 | 0.06 | 0.00 | 0.00 | 0.00 | 0.00 | 0.00 |
| | **1977** | 0.87 | 0.83 | 0.73 | 0.46 | 0.08 | 0.00 | 0.00 | 0.00 | 0.00 | 0.00 |
| | **1978** | 0.87 | 0.84 | 0.74 | 0.50 | 0.11 | 0.00 | 0.00 | 0.00 | 0.00 | 0.00 |
| | **1979** | 0.88 | 0.84 | 0.76 | 0.54 | 0.14 | 0.00 | 0.00 | 0.00 | 0.00 | 0.00 |
| | **1980** | 0.88 | 0.85 | 0.78 | 0.57 | 0.17 | 0.01 | 0.00 | 0.00 | 0.00 | 0.00 |
| | **1981** | 0.89 | 0.85 | 0.79 | 0.61 | 0.20 | 0.01 | 0.00 | 0.00 | 0.00 | 0.00 |
| | **1982** | 0.89 | 0.86 | 0.81 | 0.64 | 0.23 | 0.02 | 0.00 | 0.00 | 0.00 | 0.00 |
| | **1983** | 0.90 | 0.86 | 0.82 | 0.66 | 0.26 | 0.03 | 0.00 | 0.00 | 0.00 | 0.00 |
| | **1984** | 0.90 | 0.87 | 0.84 | 0.68 | 0.30 | 0.04 | 0.00 | 0.00 | 0.00 | 0.00 |
| | **1985** | 0.91 | 0.87 | 0.85 | 0.70 | 0.33 | 0.06 | 0.00 | 0.00 | 0.00 | 0.00 |
| | **1986** | 0.91 | 0.88 | 0.86 | 0.72 | 0.36 | 0.08 | 0.00 | 0.00 | 0.00 | 0.00 |
| | **1987** | 0.91 | 0.88 | 0.87 | 0.74 | 0.40 | 0.11 | 0.00 | 0.00 | 0.00 | 0.00 |
| | **1988** | 0.92 | 0.88 | 0.87 | 0.76 | 0.43 | 0.14 | 0.00 | 0.00 | 0.00 | 0.00 |
| | **1989** | 0.92 | 0.89 | 0.88 | 0.77 | 0.47 | 0.18 | 0.01 | 0.00 | 0.00 | 0.00 |
| | **1990** | 0.92 | 0.90 | 0.89 | 0.79 | 0.50 | 0.21 | 0.01 | 0.00 | 0.00 | 0.00 |
| | **1991** | 0.93 | 0.90 | 0.89 | 0.81 | 0.53 | 0.25 | 0.02 | 0.00 | 0.00 | 0.00 |

*Supplementary Table 40: Probability of a female in Northern Ireland in each age-band and transfusion year surviving to the end of 2019 assuming excess chronic HCV infection and excess post-transfusion hazard beyond 10 years.*

| **Age band at transfusion (years)** | | **0 - 9** | **10 - 19** | **20 - 29** | **30 - 39** | **40 - 49** | **50 - 59** | **60 - 69** | **70 - 79** | **80 - 89** | **90+** |
|---|---|---|---|---|---|---|---|---|---|---|---|
| **Midpoint age (years)** | | 5 | 15 | 25 | 35 | 45 | 55 | 65 | 75 | 85 | 95 |
| **Midpoint age after surviving 10 years (years)** | | 15 | 25 | 35 | 45 | 55 | 65 | 75 | 85 | 95 | 105 |
| **Transfusion year** | **1970** | 0.92 | 0.82 | 0.67 | 0.32 | 0.02 | 0.00 | 0.00 | 0.00 | 0.00 | 0.00 |
| | **1971** | 0.92 | 0.84 | 0.69 | 0.37 | 0.03 | 0.00 | 0.00 | 0.00 | 0.00 | 0.00 |
| | **1972** | 0.92 | 0.85 | 0.71 | 0.40 | 0.04 | 0.00 | 0.00 | 0.00 | 0.00 | 0.00 |
| | **1973** | 0.92 | 0.86 | 0.73 | 0.44 | 0.06 | 0.00 | 0.00 | 0.00 | 0.00 | 0.00 |
| | **1974** | 0.93 | 0.87 | 0.75 | 0.48 | 0.08 | 0.00 | 0.00 | 0.00 | 0.00 | 0.00 |
| | **1975** | 0.93 | 0.88 | 0.77 | 0.52 | 0.10 | 0.00 | 0.00 | 0.00 | 0.00 | 0.00 |
| | **1976** | 0.94 | 0.89 | 0.79 | 0.56 | 0.13 | 0.00 | 0.00 | 0.00 | 0.00 | 0.00 |
| | **1977** | 0.94 | 0.89 | 0.81 | 0.58 | 0.17 | 0.00 | 0.00 | 0.00 | 0.00 | 0.00 |
| | **1978** | 0.95 | 0.90 | 0.82 | 0.61 | 0.21 | 0.01 | 0.00 | 0.00 | 0.00 | 0.00 |
| | **1979** | 0.95 | 0.91 | 0.83 | 0.64 | 0.25 | 0.01 | 0.00 | 0.00 | 0.00 | 0.00 |
| | **1980** | 0.95 | 0.92 | 0.83 | 0.67 | 0.29 | 0.02 | 0.00 | 0.00 | 0.00 | 0.00 |
| | **1981** | 0.95 | 0.92 | 0.85 | 0.70 | 0.33 | 0.03 | 0.00 | 0.00 | 0.00 | 0.00 |
| | **1982** | 0.96 | 0.92 | 0.86 | 0.72 | 0.37 | 0.05 | 0.00 | 0.00 | 0.00 | 0.00 |
| | **1983** | 0.95 | 0.93 | 0.87 | 0.74 | 0.40 | 0.07 | 0.00 | 0.00 | 0.00 | 0.00 |
| | **1984** | 0.95 | 0.93 | 0.88 | 0.76 | 0.43 | 0.10 | 0.00 | 0.00 | 0.00 | 0.00 |
| | **1985** | 0.95 | 0.93 | 0.89 | 0.78 | 0.47 | 0.13 | 0.00 | 0.00 | 0.00 | 0.00 |
| | **1986** | 0.96 | 0.93 | 0.90 | 0.79 | 0.51 | 0.16 | 0.00 | 0.00 | 0.00 | 0.00 |
| | **1987** | 0.96 | 0.94 | 0.91 | 0.81 | 0.54 | 0.20 | 0.01 | 0.00 | 0.00 | 0.00 |
| | **1988** | 0.96 | 0.94 | 0.91 | 0.83 | 0.56 | 0.24 | 0.01 | 0.00 | 0.00 | 0.00 |
| | **1989** | 0.96 | 0.94 | 0.92 | 0.84 | 0.59 | 0.29 | 0.02 | 0.00 | 0.00 | 0.00 |
| | **1990** | 0.97 | 0.95 | 0.93 | 0.84 | 0.62 | 0.34 | 0.03 | 0.00 | 0.00 | 0.00 |
| | **1991** | 0.97 | 0.95 | 0.93 | 0.86 | 0.65 | 0.38 | 0.05 | 0.00 | 0.00 | 0.00 |

*Supplementary Table 41: Probability of a male in Northern Ireland in each age-band and transfusion year surviving to the end of 2019 assuming excess chronic HCV infection and excess post-transfusion hazard beyond 10 years.*

| **Age band at transfusion (years)** | | **0 - 9** | **10 - 19** | **20 - 29** | **30 - 39** | **40 - 49** | **50 - 59** | **60 - 69** | **70 - 79** | **80 - 89** | **90+** |
|---|---|---|---|---|---|---|---|---|---|---|---|
| **Midpoint age (years)** | | 5 | 15 | 25 | 35 | 45 | 55 | 65 | 75 | 85 | 95 |
| **Midpoint age after surviving 10 years (years)** | | 15 | 25 | 35 | 45 | 55 | 65 | 75 | 85 | 95 | 105 |
| **Transfusion year** | **1970** | 0.80 | 0.72 | 0.53 | 0.17 | 0.00 | 0.00 | 0.00 | 0.00 | 0.00 | 0.00 |
| | **1971** | 0.81 | 0.74 | 0.57 | 0.21 | 0.01 | 0.00 | 0.00 | 0.00 | 0.00 | 0.00 |
| | **1972** | 0.81 | 0.76 | 0.60 | 0.25 | 0.01 | 0.00 | 0.00 | 0.00 | 0.00 | 0.00 |
| | **1973** | 0.82 | 0.77 | 0.62 | 0.29 | 0.01 | 0.00 | 0.00 | 0.00 | 0.00 | 0.00 |
| | **1974** | 0.83 | 0.78 | 0.65 | 0.33 | 0.02 | 0.00 | 0.00 | 0.00 | 0.00 | 0.00 |
| | **1975** | 0.84 | 0.79 | 0.68 | 0.37 | 0.03 | 0.00 | 0.00 | 0.00 | 0.00 | 0.00 |
| | **1976** | 0.84 | 0.80 | 0.70 | 0.40 | 0.05 | 0.00 | 0.00 | 0.00 | 0.00 | 0.00 |
| | **1977** | 0.85 | 0.81 | 0.71 | 0.44 | 0.07 | 0.00 | 0.00 | 0.00 | 0.00 | 0.00 |
| | **1978** | 0.86 | 0.82 | 0.73 | 0.46 | 0.09 | 0.00 | 0.00 | 0.00 | 0.00 | 0.00 |
| | **1979** | 0.85 | 0.83 | 0.75 | 0.50 | 0.11 | 0.00 | 0.00 | 0.00 | 0.00 | 0.00 |
| | **1980** | 0.86 | 0.83 | 0.77 | 0.54 | 0.14 | 0.00 | 0.00 | 0.00 | 0.00 | 0.00 |
| | **1981** | 0.86 | 0.84 | 0.79 | 0.58 | 0.17 | 0.01 | 0.00 | 0.00 | 0.00 | 0.00 |
| | **1982** | 0.87 | 0.85 | 0.81 | 0.61 | 0.20 | 0.01 | 0.00 | 0.00 | 0.00 | 0.00 |
| | **1983** | 0.87 | 0.86 | 0.82 | 0.63 | 0.24 | 0.02 | 0.00 | 0.00 | 0.00 | 0.00 |
| | **1984** | 0.87 | 0.86 | 0.83 | 0.66 | 0.27 | 0.03 | 0.00 | 0.00 | 0.00 | 0.00 |
| | **1985** | 0.87 | 0.87 | 0.84 | 0.69 | 0.31 | 0.05 | 0.00 | 0.00 | 0.00 | 0.00 |
| | **1986** | 0.88 | 0.88 | 0.84 | 0.71 | 0.33 | 0.07 | 0.00 | 0.00 | 0.00 | 0.00 |
| | **1987** | 0.88 | 0.88 | 0.85 | 0.73 | 0.37 | 0.09 | 0.00 | 0.00 | 0.00 | 0.00 |
| | **1988** | 0.89 | 0.88 | 0.87 | 0.75 | 0.40 | 0.12 | 0.00 | 0.00 | 0.00 | 0.00 |
| | **1989** | 0.89 | 0.88 | 0.87 | 0.76 | 0.43 | 0.15 | 0.01 | 0.00 | 0.00 | 0.00 |
| | **1990** | 0.89 | 0.88 | 0.88 | 0.78 | 0.46 | 0.19 | 0.01 | 0.00 | 0.00 | 0.00 |
| | **1991** | 0.89 | 0.89 | 0.88 | 0.80 | 0.50 | 0.23 | 0.02 | 0.00 | 0.00 | 0.00 |

*Supplementary Table 42: Probability of a female in Scotland in each age-band and transfusion year surviving to the end of 2019 assuming excess chronic HCV infection and excess post-transfusion hazard beyond 10 years.*

| **Age band at transfusion (years)** | | **0 - 9** | **10 - 19** | **20 - 29** | **30 - 39** | **40 - 49** | **50 - 59** | **60 - 69** | **70 - 79** | **80 - 89** | **90+** |
|---|---|---|---|---|---|---|---|---|---|---|---|
| **Midpoint age (years)** | | 5 | 15 | 25 | 35 | 45 | 55 | 65 | 75 | 85 | 95 |
| **Midpoint age after surviving 10 years (years)** | | 15 | 25 | 35 | 45 | 55 | 65 | 75 | 85 | 95 | 105 |
| **Transfusion year** | **1970** | 0.90 | 0.80 | 0.61 | 0.27 | 0.01 | 0.00 | 0.00 | 0.00 | 0.00 | 0.00 |
| | **1971** | 0.90 | 0.82 | 0.65 | 0.31 | 0.02 | 0.00 | 0.00 | 0.00 | 0.00 | 0.00 |
| | **1972** | 0.91 | 0.83 | 0.67 | 0.34 | 0.03 | 0.00 | 0.00 | 0.00 | 0.00 | 0.00 |
| | **1973** | 0.91 | 0.84 | 0.70 | 0.38 | 0.04 | 0.00 | 0.00 | 0.00 | 0.00 | 0.00 |
| | **1974** | 0.92 | 0.85 | 0.72 | 0.42 | 0.06 | 0.00 | 0.00 | 0.00 | 0.00 | 0.00 |
| | **1975** | 0.92 | 0.86 | 0.74 | 0.45 | 0.08 | 0.00 | 0.00 | 0.00 | 0.00 | 0.00 |
| | **1976** | 0.92 | 0.87 | 0.75 | 0.49 | 0.10 | 0.00 | 0.00 | 0.00 | 0.00 | 0.00 |
| | **1977** | 0.93 | 0.87 | 0.77 | 0.52 | 0.13 | 0.00 | 0.00 | 0.00 | 0.00 | 0.00 |
| | **1978** | 0.93 | 0.88 | 0.79 | 0.56 | 0.16 | 0.01 | 0.00 | 0.00 | 0.00 | 0.00 |
| | **1979** | 0.94 | 0.89 | 0.80 | 0.59 | 0.19 | 0.01 | 0.00 | 0.00 | 0.00 | 0.00 |
| | **1980** | 0.94 | 0.89 | 0.82 | 0.62 | 0.23 | 0.02 | 0.00 | 0.00 | 0.00 | 0.00 |
| | **1981** | 0.94 | 0.90 | 0.83 | 0.65 | 0.27 | 0.02 | 0.00 | 0.00 | 0.00 | 0.00 |
| | **1982** | 0.94 | 0.90 | 0.84 | 0.68 | 0.30 | 0.03 | 0.00 | 0.00 | 0.00 | 0.00 |
| | **1983** | 0.94 | 0.91 | 0.85 | 0.70 | 0.33 | 0.05 | 0.00 | 0.00 | 0.00 | 0.00 |
| | **1984** | 0.94 | 0.91 | 0.86 | 0.72 | 0.37 | 0.07 | 0.00 | 0.00 | 0.00 | 0.00 |
| | **1985** | 0.94 | 0.92 | 0.87 | 0.74 | 0.40 | 0.09 | 0.00 | 0.00 | 0.00 | 0.00 |
| | **1986** | 0.94 | 0.92 | 0.88 | 0.76 | 0.43 | 0.12 | 0.00 | 0.00 | 0.00 | 0.00 |
| | **1987** | 0.95 | 0.92 | 0.89 | 0.78 | 0.46 | 0.16 | 0.00 | 0.00 | 0.00 | 0.00 |
| | **1988** | 0.95 | 0.93 | 0.90 | 0.79 | 0.49 | 0.20 | 0.01 | 0.00 | 0.00 | 0.00 |
| | **1989** | 0.95 | 0.93 | 0.90 | 0.81 | 0.52 | 0.24 | 0.01 | 0.00 | 0.00 | 0.00 |
| | **1990** | 0.95 | 0.93 | 0.91 | 0.82 | 0.55 | 0.28 | 0.02 | 0.00 | 0.00 | 0.00 |
| | **1991** | 0.95 | 0.93 | 0.91 | 0.84 | 0.59 | 0.32 | 0.04 | 0.00 | 0.00 | 0.00 |

*Supplementary Table 43: Probability of a male in Scotland in each age-band and transfusion year surviving to the end of 2019 assuming excess chronic HCV infection and excess post-transfusion hazard beyond 10 years.*

| **Age band at transfusion (years)** | | **0 - 9** | **10 - 19** | **20 - 29** | **30 - 39** | **40 - 49** | **50 - 59** | **60 - 69** | **70 - 79** | **80 - 89** | **90+** |
|---|---|---|---|---|---|---|---|---|---|---|---|
| **Midpoint age (years)** | | 5 | 15 | 25 | 35 | 45 | 55 | 65 | 75 | 85 | 95 |
| **Midpoint age after surviving 10 years (years)** | | 15 | 25 | 35 | 45 | 55 | 65 | 75 | 85 | 95 | 105 |
| **Transfusion year** | **1970** | 0.80 | 0.69 | 0.47 | 0.14 | 0.00 | 0.00 | 0.00 | 0.00 | 0.00 | 0.00 |
| | **1971** | 0.80 | 0.71 | 0.51 | 0.17 | 0.00 | 0.00 | 0.00 | 0.00 | 0.00 | 0.00 |
| | **1972** | 0.80 | 0.73 | 0.55 | 0.20 | 0.01 | 0.00 | 0.00 | 0.00 | 0.00 | 0.00 |
| | **1973** | 0.81 | 0.74 | 0.57 | 0.23 | 0.01 | 0.00 | 0.00 | 0.00 | 0.00 | 0.00 |
| | **1974** | 0.81 | 0.76 | 0.60 | 0.26 | 0.02 | 0.00 | 0.00 | 0.00 | 0.00 | 0.00 |
| | **1975** | 0.82 | 0.77 | 0.62 | 0.29 | 0.02 | 0.00 | 0.00 | 0.00 | 0.00 | 0.00 |
| | **1976** | 0.82 | 0.78 | 0.64 | 0.33 | 0.04 | 0.00 | 0.00 | 0.00 | 0.00 | 0.00 |
| | **1977** | 0.83 | 0.78 | 0.67 | 0.36 | 0.05 | 0.00 | 0.00 | 0.00 | 0.00 | 0.00 |
| | **1978** | 0.83 | 0.78 | 0.69 | 0.40 | 0.07 | 0.00 | 0.00 | 0.00 | 0.00 | 0.00 |
| | **1979** | 0.83 | 0.79 | 0.71 | 0.44 | 0.09 | 0.00 | 0.00 | 0.00 | 0.00 | 0.00 |
| | **1980** | 0.83 | 0.79 | 0.73 | 0.48 | 0.11 | 0.00 | 0.00 | 0.00 | 0.00 | 0.00 |
| | **1981** | 0.84 | 0.80 | 0.74 | 0.52 | 0.13 | 0.01 | 0.00 | 0.00 | 0.00 | 0.00 |
| | **1982** | 0.84 | 0.80 | 0.76 | 0.55 | 0.16 | 0.01 | 0.00 | 0.00 | 0.00 | 0.00 |
| | **1983** | 0.84 | 0.81 | 0.78 | 0.59 | 0.18 | 0.02 | 0.00 | 0.00 | 0.00 | 0.00 |
| | **1984** | 0.85 | 0.81 | 0.79 | 0.61 | 0.21 | 0.02 | 0.00 | 0.00 | 0.00 | 0.00 |
| | **1985** | 0.86 | 0.81 | 0.80 | 0.63 | 0.23 | 0.04 | 0.00 | 0.00 | 0.00 | 0.00 |
| | **1986** | 0.87 | 0.82 | 0.81 | 0.66 | 0.26 | 0.05 | 0.00 | 0.00 | 0.00 | 0.00 |
| | **1987** | 0.87 | 0.82 | 0.82 | 0.68 | 0.29 | 0.07 | 0.00 | 0.00 | 0.00 | 0.00 |
| | **1988** | 0.87 | 0.82 | 0.83 | 0.70 | 0.33 | 0.09 | 0.00 | 0.00 | 0.00 | 0.00 |
| | **1989** | 0.88 | 0.82 | 0.83 | 0.72 | 0.36 | 0.12 | 0.00 | 0.00 | 0.00 | 0.00 |
| | **1990** | 0.89 | 0.83 | 0.84 | 0.74 | 0.40 | 0.15 | 0.01 | 0.00 | 0.00 | 0.00 |
| | **1991** | 0.90 | 0.84 | 0.85 | 0.76 | 0.44 | 0.18 | 0.01 | 0.00 | 0.00 | 0.00 |

*Supplementary Table 44: Probability of a female in Wales in each age-band and transfusion year surviving to the end of 2019 assuming excess chronic HCV infection and excess post-transfusion hazard beyond 10 years.*

| **Age band at transfusion (years)** | | **0 - 9** | **10 - 19** | **20 - 29** | **30 - 39** | **40 - 49** | **50 - 59** | **60 - 69** | **70 - 79** | **80 - 89** | **90+** |
|---|---|---|---|---|---|---|---|---|---|---|---|
| **Midpoint age (years)** | | 5 | 15 | 25 | 35 | 45 | 55 | 65 | 75 | 85 | 95 |
| **Midpoint age after surviving 10 years (years)** | | 15 | 25 | 35 | 45 | 55 | 65 | 75 | 85 | 95 | 105 |
| **Transfusion year** | **1970** | 0.91 | 0.82 | 0.66 | 0.32 | 0.02 | 0.00 | 0.00 | 0.00 | 0.00 | 0.00 |
| | **1971** | 0.92 | 0.84 | 0.69 | 0.36 | 0.03 | 0.00 | 0.00 | 0.00 | 0.00 | 0.00 |
| | **1972** | 0.92 | 0.85 | 0.71 | 0.40 | 0.04 | 0.00 | 0.00 | 0.00 | 0.00 | 0.00 |
| | **1973** | 0.93 | 0.86 | 0.74 | 0.44 | 0.06 | 0.00 | 0.00 | 0.00 | 0.00 | 0.00 |
| | **1974** | 0.93 | 0.87 | 0.75 | 0.48 | 0.08 | 0.00 | 0.00 | 0.00 | 0.00 | 0.00 |
| | **1975** | 0.93 | 0.88 | 0.77 | 0.51 | 0.10 | 0.00 | 0.00 | 0.00 | 0.00 | 0.00 |
| | **1976** | 0.93 | 0.88 | 0.79 | 0.55 | 0.14 | 0.00 | 0.00 | 0.00 | 0.00 | 0.00 |
| | **1977** | 0.94 | 0.89 | 0.80 | 0.58 | 0.17 | 0.00 | 0.00 | 0.00 | 0.00 | 0.00 |
| | **1978** | 0.94 | 0.90 | 0.81 | 0.62 | 0.21 | 0.01 | 0.00 | 0.00 | 0.00 | 0.00 |
| | **1979** | 0.94 | 0.90 | 0.82 | 0.64 | 0.25 | 0.01 | 0.00 | 0.00 | 0.00 | 0.00 |
| | **1980** | 0.94 | 0.91 | 0.83 | 0.66 | 0.28 | 0.02 | 0.00 | 0.00 | 0.00 | 0.00 |
| | **1981** | 0.94 | 0.91 | 0.84 | 0.69 | 0.32 | 0.03 | 0.00 | 0.00 | 0.00 | 0.00 |
| | **1982** | 0.94 | 0.92 | 0.86 | 0.72 | 0.36 | 0.05 | 0.00 | 0.00 | 0.00 | 0.00 |
| | **1983** | 0.95 | 0.92 | 0.87 | 0.74 | 0.39 | 0.07 | 0.00 | 0.00 | 0.00 | 0.00 |
| | **1984** | 0.95 | 0.92 | 0.88 | 0.76 | 0.43 | 0.09 | 0.00 | 0.00 | 0.00 | 0.00 |
| | **1985** | 0.96 | 0.93 | 0.89 | 0.77 | 0.46 | 0.12 | 0.00 | 0.00 | 0.00 | 0.00 |
| | **1986** | 0.96 | 0.93 | 0.89 | 0.79 | 0.49 | 0.16 | 0.00 | 0.00 | 0.00 | 0.00 |
| | **1987** | 0.96 | 0.93 | 0.90 | 0.81 | 0.52 | 0.20 | 0.01 | 0.00 | 0.00 | 0.00 |
| | **1988** | 0.97 | 0.93 | 0.91 | 0.82 | 0.55 | 0.24 | 0.01 | 0.00 | 0.00 | 0.00 |
| | **1989** | 0.97 | 0.94 | 0.92 | 0.83 | 0.58 | 0.28 | 0.02 | 0.00 | 0.00 | 0.00 |
| | **1990** | 0.97 | 0.94 | 0.92 | 0.84 | 0.61 | 0.33 | 0.03 | 0.00 | 0.00 | 0.00 |
| | **1991** | 0.97 | 0.94 | 0.93 | 0.85 | 0.64 | 0.37 | 0.05 | 0.00 | 0.00 | 0.00 |

*Supplementary Table 45: Probability of a male in Wales in each age-band and transfusion year surviving to the end of 2019 assuming excess chronic HCV infection and excess post-transfusion hazard beyond 10 years.*

| **Age band at transfusion (years)** | | **0 - 9** | **10 - 19** | **20 - 29** | **30 - 39** | **40 - 49** | **50 - 59** | **60 - 69** | **70 - 79** | **80 - 89** | **90+** |
|---|---|---|---|---|---|---|---|---|---|---|---|
| **Midpoint age (years)** | | 5 | 15 | 25 | 35 | 45 | 55 | 65 | 75 | 85 | 95 |
| **Midpoint age after surviving 10 years (years)** | | 15 | 25 | 35 | 45 | 55 | 65 | 75 | 85 | 95 | 105 |
| **Transfusion year** | **1970** | 0.82 | 0.74 | 0.54 | 0.18 | 0.00 | 0.00 | 0.00 | 0.00 | 0.00 | 0.00 |
| | **1971** | 0.83 | 0.76 | 0.57 | 0.21 | 0.01 | 0.00 | 0.00 | 0.00 | 0.00 | 0.00 |
| | **1972** | 0.84 | 0.77 | 0.60 | 0.25 | 0.01 | 0.00 | 0.00 | 0.00 | 0.00 | 0.00 |
| | **1973** | 0.84 | 0.79 | 0.63 | 0.29 | 0.02 | 0.00 | 0.00 | 0.00 | 0.00 | 0.00 |
| | **1974** | 0.85 | 0.80 | 0.65 | 0.32 | 0.03 | 0.00 | 0.00 | 0.00 | 0.00 | 0.00 |
| | **1975** | 0.85 | 0.80 | 0.67 | 0.36 | 0.04 | 0.00 | 0.00 | 0.00 | 0.00 | 0.00 |
| | **1976** | 0.85 | 0.81 | 0.70 | 0.40 | 0.05 | 0.00 | 0.00 | 0.00 | 0.00 | 0.00 |
| | **1977** | 0.85 | 0.82 | 0.72 | 0.44 | 0.07 | 0.00 | 0.00 | 0.00 | 0.00 | 0.00 |
| | **1978** | 0.85 | 0.82 | 0.73 | 0.48 | 0.09 | 0.00 | 0.00 | 0.00 | 0.00 | 0.00 |
| | **1979** | 0.86 | 0.82 | 0.75 | 0.52 | 0.12 | 0.00 | 0.00 | 0.00 | 0.00 | 0.00 |
| | **1980** | 0.86 | 0.83 | 0.76 | 0.55 | 0.14 | 0.00 | 0.00 | 0.00 | 0.00 | 0.00 |
| | **1981** | 0.86 | 0.83 | 0.78 | 0.58 | 0.17 | 0.01 | 0.00 | 0.00 | 0.00 | 0.00 |
| | **1982** | 0.86 | 0.84 | 0.80 | 0.62 | 0.20 | 0.01 | 0.00 | 0.00 | 0.00 | 0.00 |
| | **1983** | 0.86 | 0.84 | 0.81 | 0.64 | 0.23 | 0.02 | 0.00 | 0.00 | 0.00 | 0.00 |
| | **1984** | 0.87 | 0.84 | 0.82 | 0.66 | 0.27 | 0.03 | 0.00 | 0.00 | 0.00 | 0.00 |
| | **1985** | 0.88 | 0.85 | 0.83 | 0.68 | 0.30 | 0.05 | 0.00 | 0.00 | 0.00 | 0.00 |
| | **1986** | 0.89 | 0.85 | 0.84 | 0.70 | 0.33 | 0.07 | 0.00 | 0.00 | 0.00 | 0.00 |
| | **1987** | 0.89 | 0.86 | 0.85 | 0.72 | 0.37 | 0.09 | 0.00 | 0.00 | 0.00 | 0.00 |
| | **1988** | 0.90 | 0.86 | 0.86 | 0.74 | 0.41 | 0.12 | 0.00 | 0.00 | 0.00 | 0.00 |
| | **1989** | 0.91 | 0.86 | 0.87 | 0.76 | 0.44 | 0.16 | 0.01 | 0.00 | 0.00 | 0.00 |
| | **1990** | 0.91 | 0.86 | 0.87 | 0.77 | 0.47 | 0.19 | 0.01 | 0.00 | 0.00 | 0.00 |
| | **1991** | 0.92 | 0.87 | 0.88 | 0.79 | 0.51 | 0.23 | 0.02 | 0.00 | 0.00 | 0.00 |

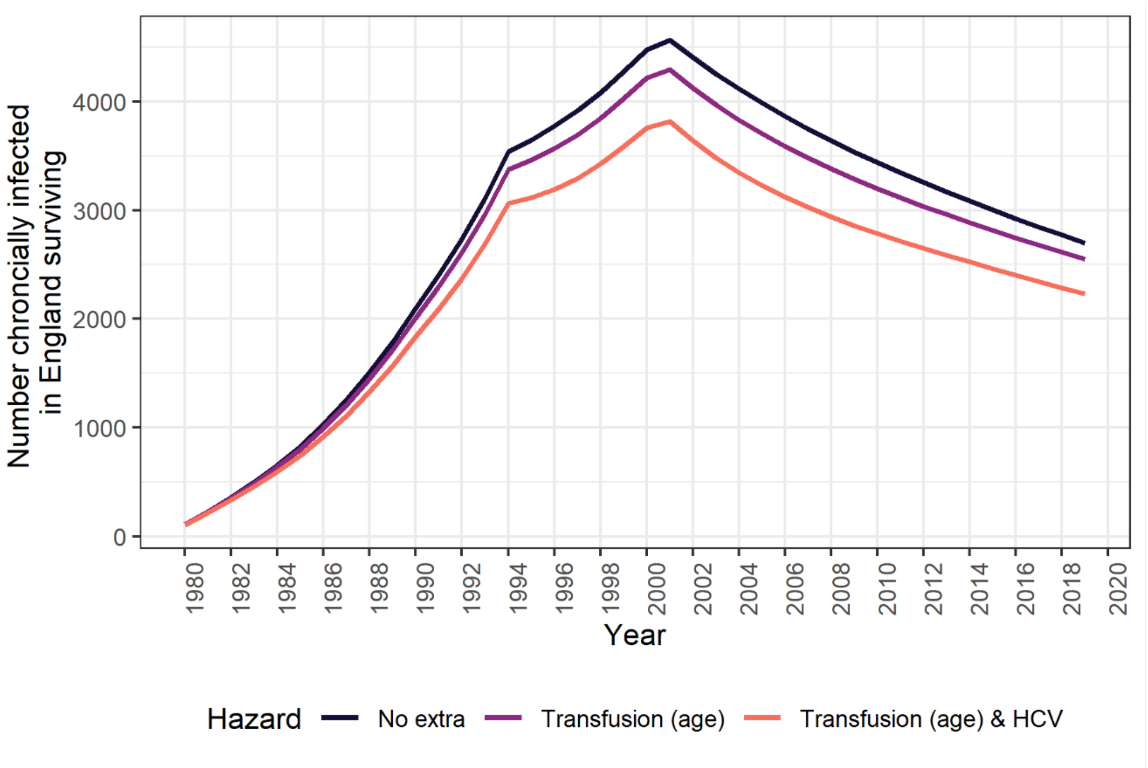

*Supplementary Figure 7: The number of chronically HCV-infected people in England surviving to each year under three different combinations of 'hazards' - the annual risk of dying - in the baseline deterministic model. (1) no increased risk from transfusion or chronic HCV infection, (2) increased risk following transfusion, (3) increased risk following transfusion and chronic HCV infection. There is an increase in the number of people surviving to 2001 (the point at which all people transfused between 1970 and 1991 could have survived to 10 years), and after this there is a steady fall in the number of people surviving.*

*Supplementary Table 46: Estimated annual number of individuals chronically HCV-infected by transfusion in England, 1970-August 1991, and surviving to the end of 2019, both without and with allowing for any effect of chronic HCV infection. Estimates are provided as median and 95% uncertainty intervals. The estimated numbers displayed for the year of transfusion of 1991 are for January to August 1991 and not the full 12-month year.*

| Year of transfusion | Year 10-years post-transfusion | (a) Estimated number chronically HCV-infected surviving 10 years post-transfusion (from Task 4) | (b) Estimated number chronically HCV-infected surviving to 2019, assuming no post-transfusion excess risk | (c) Estimated number surviving to 2019, assuming post-transfusion excess risk | (d) Estimated number surviving to 2019, assuming both post-transfusion and chronic HCV-infection excess risk |
|---|---|---|---|---|---|
| 1970 | 1980 | 120 (78 - 190) | 32 (19 - 54) | 30 (18 - 52) | 25 (15 - 44) |
| 1971 | 1981 | 130 (89 - 210) | 37 (23 - 62) | 35 (22 - 59) | 30 (18 - 51) |
| 1972 | 1982 | 150 (100 - 240) | 44 (27 - 73) | 41 (25 - 70) | 35 (21 - 60) |
| 1973 | 1983 | 170 (120 - 280) | 51 (32 - 85) | 49 (31 - 81) | 42 (25 - 70) |
| 1974 | 1984 | 190 (130 - 320) | 60 (38 - 100) | 57 (37 - 97) | 48 (30 - 83) |
| 1975 | 1985 | 210 (140 - 350) | 68 (44 - 110) | 65 (42 - 110) | 56 (35 - 95) |
| 1976 | 1986 | 260 (180 - 430) | 85 (55 - 140) | 81 (53 - 140) | 70 (44 - 120) |
| 1977 | 1987 | 280 (190 - 480) | 97 (64 - 170) | 93 (60 - 160) | 80 (52 - 140) |
| 1978 | 1988 | 330 (230 - 560) | 120 (78 - 200) | 110 (74 - 190) | 96 (63 - 170) |
| 1979 | 1989 | 370 (250 - 630) | 140 (89 - 230) | 130 (84 - 220) | 110 (71 - 190) |
| 1980 | 1990 | 410 (280 - 690) | 150 (100 - 260) | 140 (98 - 250) | 130 (83 - 220) |
| 1981 | 1991 | 410 (280 - 700) | 160 (100 - 270) | 150 (99- 260) | 130 (84 - 230) |
| 1982 | 1992 | 440 (300 - 750) | 180 (120 - 300) | 170 (110 - 290) | 140 (95 - 250) |
| 1983 | 1993 | 500 (340 - 870) | 210 (140 - 360) | 190 (130 - 340) | 170 (110 - 300) |
| 1984 | 1994 | 560 (380 - 960) | 240 (160 - 410) | 220 (150 - 380) | 200 (130 - 340) |
| 1985 | 1995 | 250 (200 - 310) | 110 (85 - 140) | 100 (79 - 130) | 91 (68 - 120) |
| 1986 | 1996 | 280 (220 - 340) | 120 (97 - 160) | 120 (90 - 150) | 100 (78 - 130) |
| 1987 | 1997 | 300 (240 - 360) | 140 (110 - 170) | 130 (100 - 160) | 110 (87 - 150) |
| 1988 | 1998 | 330 (270 - 400) | 160 (120 - 200) | 150 (120 - 190) | 130 (99 - 170) |
| 1989 | 1999 | 370 (300 - 450) | 180 (140 - 220) | 170 (130 - 210) | 150 (110 - 190) |
| 1990 | 2000 | 370 (300 - 450) | 190 (150 - 240) | 180 (140 - 220) | 160 (120 - 200) |
| 1991 | 2001 | 250 (210 - 300) | 130 (110 - 160) | 120 (100 - 150) | 110 (85 - 140) |
| Total | | 6,700 (5,200 - 9,800) | 2,700 (2,100 - 3,900) | 2,600 (2,000 - 3,700) | 2,200 (1,700 - 3,200) |
| Total for 1970-1979 | | 2,200 (1,600 - 3,600) | 720 (510 - 1,200) | 690 (490 - 1,100) | 590 (410 - 980) |
| Total for 1980-August 1991 | | 4,500 (3,600 - 6,200) | 2,000 (1,600 - 2,700) | 1,900 (1,500 - 2,500) | 1,600 (1,300 - 2,200) |

*Supplementary Table 47: Estimated annual number of individuals chronically HCV-infected by transfusion in Northern Ireland, 1970-August 1991, and surviving to the end of 2019, both without and with allowing for any effect of chronic HCV infection. Estimates are provided as median and 95% uncertainty intervals. The estimated numbers displayed for the year of transfusion of 1991 are for January to August 1991 and not the full 12-month year.*

| Year of transfusion | Year 10-years post-transfusion | (a) Estimated number chronically HCV-infected surviving 10 years post-transfusion (from Task 4) | (b) Estimated number chronically HCV-infected surviving to 2019, assuming no post-transfusion excess risk | (c) Estimated number surviving to 2019, assuming post-transfusion excess risk | (d) Estimated number surviving to 2019, assuming both post-transfusion and chronic HCV-infection excess risk |
|---|---|---|---|---|---|
| 1970 | 1980 | 4 (1 - 9) | 1 (0 - 3) | 1 (0 - 3) | 1 (0 -3) |
| 1971 | 1981 | 4 (1 - 10) | 1 (0 - 4) | 1 (0 - 4) | 1 (0 -3) |
| 1972 | 1982 | 5 (1 - 11) | 1 (0 - 4) | 1 (0 - 4) | 1 (0 - 4) |
| 1973 | 1983 | 5 (1 - 12) | 2 (0 - 5) | 2 (0 - 5) | 1 (0 - 4) |
| 1974 | 1984 | 6 (2 - 13) | 2 (0 - 6) | 2 (0 - 5) | 2 (0 - 5) |
| 1975 | 1985 | 7 (2 - 14) | 2 (0 - 6) | 2 (0 - 6) | 2 (0 - 5) |
| 1976 | 1986 | 8 (3 - 17) | 3 (0 - 7) | 3 (0 - 7) | 2 (0 - 6) |
| 1977 | 1987 | 9 (4 - 19) | 3 (0 - 8) | 3 (0 - 8) | 3 (0 - 7) |
| 1978 | 1988 | 11 (5 - 21) | 4 (1 - 9) | 4 (1 - 9) | 3 (0 - 8) |
| 1979 | 1989 | 12 (5 - 24) | 4 (1 - 10) | 4 (1 - 9) | 4 (1 - 9) |
| 1980 | 1990 | 14 (6 - 26) | 5 (1 - 11) | 5 (1 - 11) | 4 (1 - 10) |
| 1981 | 1991 | 14 (6 - 26) | 5 (1 - 11) | 5 (1 - 11) | 5 (1 - 10) |
| 1982 | 1992 | 15 (6 - 28) | 6 (2 - 13) | 6 (2 - 12) | 5 (1 - 11) |
| 1983 | 1993 | 17 (8 - 32) | 7 (2 - 15) | 7 (2 - 14) | 6 (2 - 12) |
| 1984 | 1994 | 19 (9 - 36) | 8 (3 - 17) | 8 (3 - 16) | 7 (2 - 14) |
| 1985 | 1995 | 8 (3 - 15) | 4 (1 - 8) | 3 (0 - 8) | 3 (0 - 7) |
| 1986 | 1996 | 9 (4 - 16) | 4 (1 - 9) | 4 (1 - 8) | 3 (0 - 8) |
| 1987 | 1997 | 10 (4 - 17) | 4 (1 - 9) | 4 (1 - 9) | 4 (1 - 8) |
| 1988 | 1998 | 11 (5 - 18) | 5 (1 - 10) | 5 (1 - 10) | 4 (1 - 9) |
| 1989 | 1999 | 12 (6 - 20) | 6 (2 - 12) | 5 (2 - 11) | 5 (1 - 10) |
| 1990 | 2000 | 12 (6 - 20) | 6 (2 - 12) | 6 (2 - 11) | 5 (1 - 10) |
| 1991 | 2001 | 8 (3 - 15) | 4 (1 - 9) | 4 (1 - 9) | 4 (0 - 8) |
| Total | | 220 (160 - 330) | 92 (65 - 140) | 87 (62 - 130) | 76 (53 - 110) |
| Total for 1970-1979 | | 73 (48 - 120) | 25 (14 - 43) | 24 (13 - 42) | 21 (11 - 36) |
| Total for 1980-August 1991 | | 150 (110 - 210) | 67 (47 - 96) | 63 (44 - 91) | 56 (38 - 82) |

*Supplementary Table 48: Estimated annual number of individuals chronically HCV-infected by transfusion in Scotland, 1970-August 1991, and surviving to the end of 2019, both without and with allowing for any effect of chronic HCV infection. Estimates are provided as median and 95% uncertainty intervals. The estimated numbers displayed for the year of transfusion of 1991 are for January to August 1991 and not the full 12-month year.*

| **Year of transfusion** | **Year 10-years post-transfusion** | **(a) Estimated number chronically HCV-infected surviving 10 years post-transfusion (from Task 4)** | **(b) Estimated number chronically HCV-infected surviving to 2019, assuming no post-transfusion excess risk** | **(c) Estimated number surviving to 2019, assuming post-transfusion excess risk** | **(d) Estimated number surviving to 2019, assuming both post-transfusion and chronic HCV-infection excess risk** |
|---|---|---|---|---|---|
| 1970 | 1980 | 12 (6 - 21) | 3 (0 - 7) | 3 (0 - 7) | 2 (0 - 6) |
| 1971 | 1981 | 13 (6 - 22) | 3 (0 - 7) | 3 (0 -7) | 3 (0 - 6) |
| 1972 | 1982 | 14 (7 - 23) | 4 (1 - 8) | 4 (1 - 8) | 3 (0 - 7) |
| 1973 | 1983 | 15 (8 - 24) | 4 (1 - 9) | 4 (1 - 8) | 3 (1 - 7) |
| 1974 | 1984 | 16 (8 - 26) | 5 (1 - 9) | 5 (1 - 9) | 4 (1 - 8) |
| 1975 | 1985 | 16 (8 - 26) | 5 (1 - 10) | 5 (1 - 9) | 4 (1 - 8) |
| 1976 | 1986 | 18 (9 - 29) | 6 (2 - 11) | 5 (1 -10) | 4 (1 - 9) |
| 1977 | 1987 | 21 (12 - 34) | 7 (3 -13) | 7 (3 - 13) | 6 (2 - 11) |
| 1978 | 1988 | 24 (14 - 39) | 8 (3 - 15) | 8 (3 - 15) | 7 (3 - 12) |
| 1979 | 1989 | 29 (17 -46) | 10 (4 - 18) | 10 (4 -17) | 8 (3 - 15) |
| 1980 | 1990 | 33 (20 - 54) | 12 (6 - 21) | 11 (5 - 20) | 9 (4 - 17) |
| 1981 | 1991 | 39 (24 - 64) | 15 (8 - 26) | 14 (7 - 24) | 12 (6 - 21) |
| 1982 | 1992 | 49 (30 - 82) | 19 (10 - 33) | 17 (9 - 31) | 15 (8 - 27) |
| 1983 | 1993 | 63 (39 - 110) | 24 (14 - 44) | 23 (12 - 41) | 20 (11 - 36) |
| 1984 | 1994 | 37 (24 - 52) | 15 (8 - 24) | 14 (8 - 22) | 12 (6 - 19) |
| 1985 | 1995 | 44 (30 - 61) | 19 (11 - 28) | 17 (10 - 26) | 15 (8 - 23) |
| 1986 | 1996 | 53 (36 - 72) | 23 (14 - 33) | 21 (13 - 31) | 18 (11 - 27) |
| 1987 | 1997 | 54 (38 - 74) | 24 (15 - 35) | 23 (14 - 32) | 19 (11 - 29) |
| 1988 | 1998 | 64 (46 - 86) | 30 (19 - 42) | 27 (18 - 38) | 24 (15 - 34) |
| 1989 | 1999 | 72 (51 - 96) | 34 (23 - 48) | 32 (21 - 44) | 27 (17 - 39) |
| 1990 | 2000 | 79 (58 - 100) | 39 (27 - 54) | 36 (25 - 49) | 31 (20 - 44) |
| 1991 | 2001 | 59 (42 - 80) | 31 (20 - 43) | 28 (18 - 40) | 24 (15 - 35) |
| Total | | 830 (660 - 1,100) | 340 (270 - 440) | 320 (250 - 410) | 270 (210 - 360) |
| Total for 1970-1979 | | 180 (130 - 25) | 56 (38 - 80) | 54 (37 - 77) | 45 (29 - 66) |
| Total for 1980-August 1991 | | 650 (520 - 820) | 290 (220 - 360) | 270 (210 - 340) | 230 (180 - 300) |

*Supplementary Table 49: Estimated annual number of individuals chronically HCV-infected by transfusion in Wales, 1970-August 1991, and surviving to the end of 2019, both without and with allowing for any effect of chronic HCV infection. Estimates are provided as median and 95% uncertainty intervals. The estimated numbers displayed for the year of transfusion of 1991 are for January to August 1991 and not the full 12-month year.*

| **Year of transfusion** | **Year 10-years post-transfusion** | **(a) Estimated number chronically HCV-infected surviving 10 years post-transfusion (from Task 4)** | **(b) Estimated number chronically HCV-infected surviving to 2019, assuming no post-transfusion excess risk** | **(c) Estimated number surviving to 2019, assuming post-transfusion excess risk** | **(d) Estimated number surviving to 2019, assuming both post-transfusion and chronic HCV-infection excess risk** |
|---|---|---|---|---|---|
| 1970 | 1980 | 7 (2 - 14) | 2 (0 - 5) | 2 (0 - 5) | 1 (0 - 4) |
| 1971 | 1981 | 8 (3 - 16) | 2 (0 - 6) | 2 (0 - 6) | 2 (0 - 5) |
| 1972 | 1982 | 9 (3 - 17) | 3 (0 - 6) | 3 (0 - 6) | 2 (0 - 5) |
| 1973 | 1983 | 10 (4 - 20) | 3 (0 - 7) | 3 (0 - 7) | 2 (0 - 6) |
| 1974 | 1984 | 11 (5 - 22) | 4 (1 - 9) | 3 (0 - 8) | 3 (0 - 7) |
| 1975 | 1985 | 13 (6 - 24) | 4 (1 - 9) | 4 (1 - 9) | 3 (0 - 8) |
| 1976 | 1986 | 15 (7 - 29) | 5 (1 - 11) | 5 (1 - 11) | 4 (1 - 9) |
| 1977 | 1987 | 17 (8 - 32) | 6 (2 - 12) | 6 (2 - 12) | 5 (1 - 10) |
| 1978 | 1988 | 20 (10 - 37) | 7 (2 - 15) | 7 (2 - 14) | 6 (2 - 12) |
| 1979 | 1989 | 22 (11 - 41) | 8 (3 - 16) | 8 (3 - 16) | 7 (2 - 14) |
| 1980 | 1990 | 25 (13 - 45) | 9 (4 - 18) | 9 (3 - 18) | 8 (3 - 15) |
| 1981 | 1991 | 25 (13 - 46) | 10 (4 - 19) | 9 (3 - 18) | 8 (3 - 16) |
| 1982 | 1992 | 26 (14 - 49) | 11 (4 - 21) | 10 (4 - 20) | 9 (3 - 17) |
| 1983 | 1993 | 30 (16 - 56) | 12 (5 - 24) | 12 (5 - 23) | 10 (4 - 20) |
| 1984 | 1994 | 33 (19 - 62) | 15 (7 - 28) | 13 (6 - 26) | 12 (5 - 23) |
| 1985 | 1995 | 15 (8 - 24) | 7 (2 - 12) | 6 (2 - 12) | 5 (2 - 10) |
| 1986 | 1996 | 16 (9 - 26) | 7 (3 - 13) | 7 (3 - 13) | 6 (2 - 12) |
| 1987 | 1997 | 18 (9 - 28) | 8 (3 - 14) | 8 (3 - 14) | 7 (2 - 12) |
| 1988 | 1998 | 20 (11 - 30) | 9 (4 - 16) | 9 (4 - 15) | 8 (3 - 14) |
| 1989 | 1999 | 22 (12 - 33) | 11 (5 - 18) | 10 (4 - 17) | 9 (4 - 15) |
| 1990 | 2000 | 22 (13 - 33) | 11 (5 - 18) | 10 (5 - 17) | 9 (4 - 16) |
| 1991 | 2001 | 15 (8 - 24) | 8 (3 - 14) | 7 (3 - 13) | 7 (2 - 12) |
| Total | | 400<br>(300 - 600) | 160<br>(120 - 240) | 160<br>(120 - 230) | 140<br>(98 - 200) |
| Total for 1970-1979 | | 130<br>(90 - 220) | 44<br>(27 - 76) | 43<br>(27 - 73) | 36<br>(22 - 62) |
| Total for 1980-August 1991 | | 270<br>(200 - 380) | 120<br>(88 - 170) | 110<br>(83 - 160) | 99<br>(72 - 140) |

*Supplementary Table 50: Estimated age distribution of people with chronic HCV infection from transfusion between January 1970 and August 1991 in each nation, who are alive in December 2019 by age-band at December 2019. Estimates are provided as median and 95% uncertainty intervals.*

| **Age in December 2019 (years)** | **Estimated number alive at the end of December 2019** | | | | | | | |
|---|---|---|---|---|---|---|---|---|
| | **England** | | **Northern Ireland** | | **Scotland** | | **Wales** | |
| | **Females** | **Males** | **Females** | **Males** | **Females** | **Males** | **Females** | **Males** |
| 30 – 39 | 62<br>(38 - 91) | 120<br>(81 - 160) | 2<br>(0 - 6) | 4<br>(1 - 9) | 12<br>(5 - 22) | 23<br>(13 - 34) | 4<br>(0 - 8) | 7<br>(2 - 14) |
| 40 – 49 | 150<br>(97 - 240) | 220<br>(140 - 370) | 5<br>(1 - 11) | 8<br>(3 - 16) | 18<br>(9 - 31) | 22<br>(13 - 36) | 9<br>(3 - 18) | 14<br>(7 - 26) |
| 50 – 59 | 280<br>(210 - 380) | 130<br>(88 - 200) | 10<br>(4 - 18) | 5<br>(1 - 11) | 46<br>(31 - 64) | 18<br>(10 - 28) | 18<br>(10 - 29) | 9<br>(3 - 17) |
| 60 – 69 | 450<br>(330 - 690) | 140<br>(98 - 220) | 17<br>(9 - 28) | 6<br>(2 - 11) | 52<br>(35 - 74) | 19<br>(11 - 29) | 28<br>(17 - 46) | 10<br>(4 - 18) |
| 70 – 79 | 320<br>(230 - 500) | 130<br>(86 - 210) | 12<br>(6 - 20) | 4<br>(1 - 9) | 33<br>(21 - 49) | 13<br>(6 - 22) | 20<br>(11 - 32) | 7<br>(3 - 14) |
| 80 – 89 | 140<br>(86 - 230) | 56<br>(29 - 99) | 2<br>(0 - 7) | 0<br>(0 - 2) | 11<br>(5 - 21) | 3<br>(0 - 8) | 6<br>(1 - 13) | 1<br>(0 - 5) |
| 90+ | 16<br>(4 - 41) | 4<br>(0 - 14) | 0<br>(0 - 0) | 0<br>(0 - 0) | 0<br>(0 - 2) | 0<br>(0 - 0) | 0<br>(0 - 1) | 0<br>(0 - 0) |
| Total | 1,400<br>(1,100 - 2,100) | 800<br>(590 - 1,200) | 49<br>(32 - 74) | 28<br>(16 - 44) | 170<br>(130 - 230) | 99<br>(70 - 130) | 86<br>(60 - 130) | 50<br>(32 - 76) |

Supplementary Figure 8: Distributions for quantities of interest arising from 10,000 simulations from the stochastic Monte Carlo baseline model for England.Supplementary Figure 8 presents the distributions of quantities of interest for England under 10,000 realisations of the stochastic model.

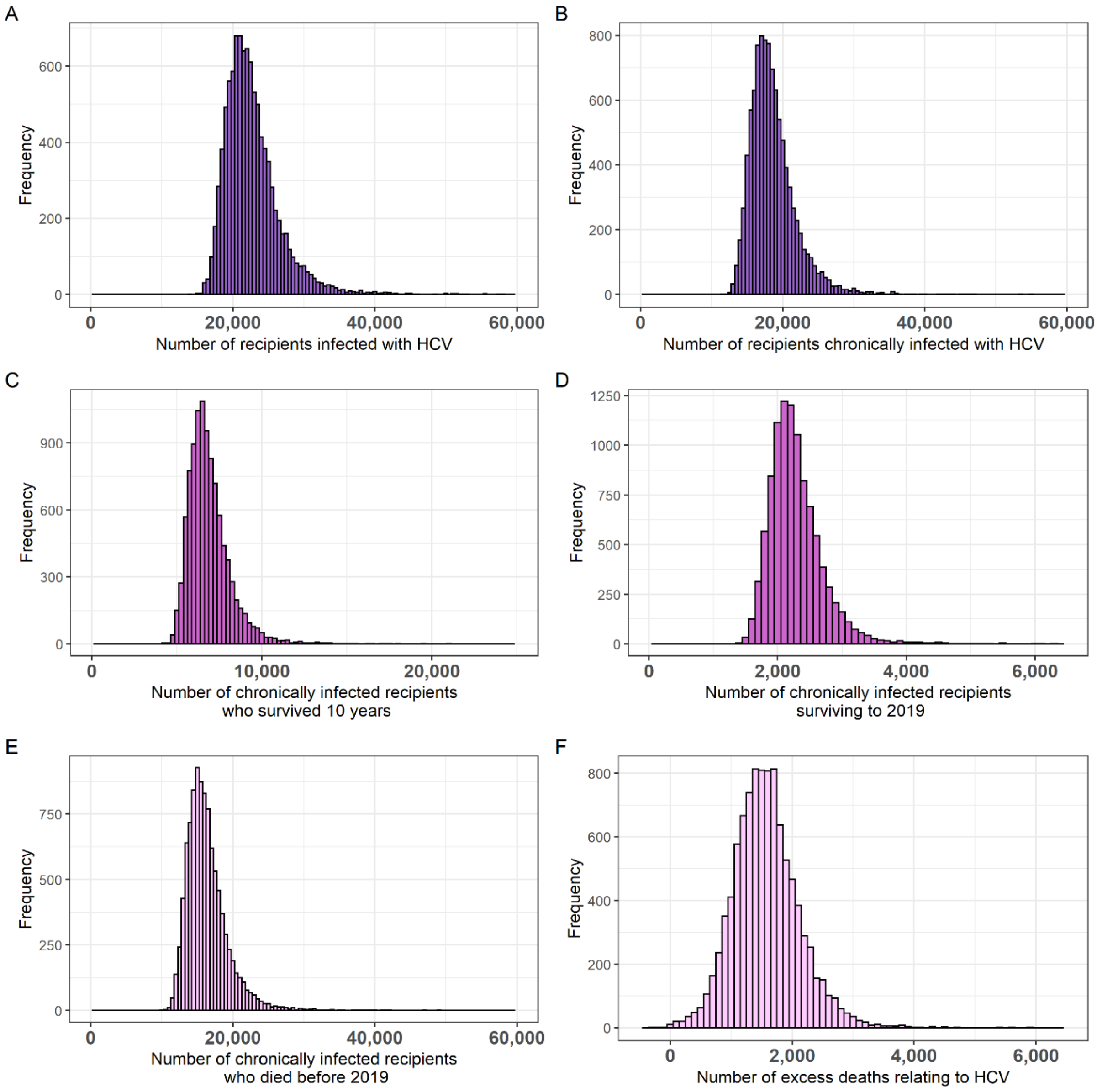

*Supplementary Figure 8: Distributions for quantities of interest arising from 10,000 simulations from the stochastic Monte Carlo baseline model for England.*

### Sensitivity analysis

An example of the sensitivity analyses conducted is illustrated in Supplementary Figure 9.

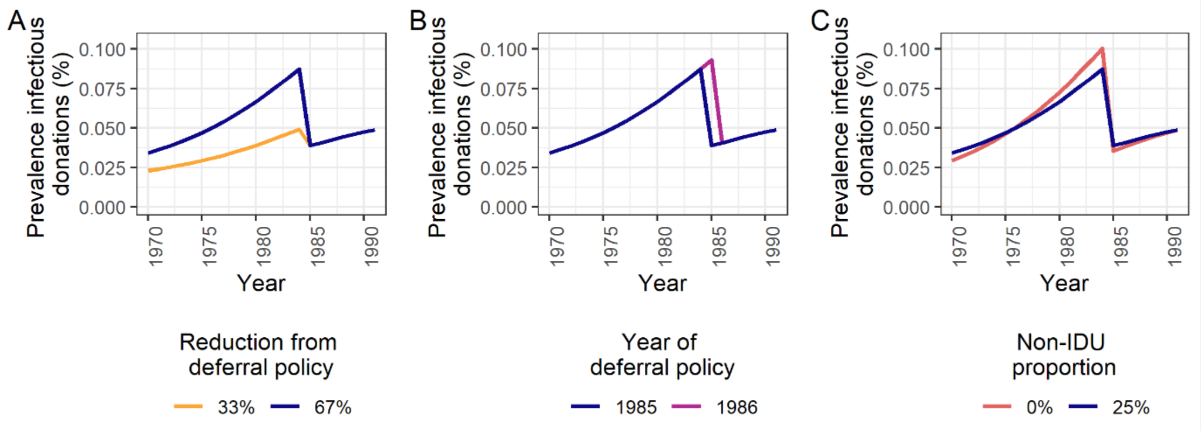

*Supplementary Figure 9: Example of the deterministic sensitivity analyses conducted for England. The blue line is the same throughout and corresponds to a deferral effect of 67% in 1985 for ever-IDUs (assumption (d)) with no proportion of prevalence coming from non-IDUs. (A) Shows the effect of a reduction in the deferral effect to 33%. (B) Shows the impact of the deferral effect occurring in 1986 rather than 1985 (assumption (c)). (C) Shows the effect of our baseline hybrid model (assumption (e)), in which there is a constant non-IDU contribution, set at 25% of the prevalence in 1991.*

Supplementary Table 51 presents the results of the 9 principal sensitivity analysis scenarios as presented in Table 3 for each of the four nations.

Supplementary Figure 10 presents the estimated HCV-IDU epidemics in England and Scotland, shown to be a key driver of the difference in results in these two countries.

*Supplementary Table 51: Results of the deterministic baseline model and the 9 deterministic sensitivity analyses. The full description of each sensitivity analysis scenario is presented in Table 3. No data on past-IDUs in Scotland were available hence scenario A is omitted for this setting.*

| **Scenario** | **Brief description** | **Infected** | **Chronically HCV-infected** | **Chronically HCV-infected, survived to 10 years post-transfusion** | **Chronically HCV-infected, survived to 2019 (assuming no extra HCV risk)** | **Chronically HCV-infected, died by 2019 (assuming no extra HCV risk)** | **Chronically HCV-infected, died by 2019, extra deaths related to HCV** |
|---|---|---|---|---|---|---|---|
| **England** | | | | | | | |
| **Baseline** | | 22,100 | 18,100 | 6,700 | 2,220 | 15,900 | 1,550 |
| **A** | Past IDUs; later (date) and lower (%) deferral | 15,700 | 12,800 | 4,740 | 1,640 | 11,200 | 1,080 |
| **B** | No non-IDUs | 22,800 | 18,700 | 6,900 | 2,300 | 16,400 | 1,590 |
| **C** | All IDUs | 20,100 | 16,500 | 6,190 | 2,020 | 14,500 | 1,410 |
| **D** | Deferral one year later (date) | 23,300 | 19,100 | 7,060 | 2,360 | 16,700 | 1,630 |
| **E** | Lower deferral effect (%) | 16,000 | 13,200 | 4,860 | 1,670 | 11,500 | 1,110 |
| **F** | 50% contribution from IDUs | 21,400 | 17,600 | 6,500 | 2,160 | 15,400 | 1,500 |
| **G** | No additional transfusion hazards beyond 10 years | 22,100 | 18,100 | 6,700 | 2,400 | 15,700 | 1,490 |
| **H** | Non-age-stratified transfusion hazards | 22,100 | 18,100 | 6,700 | 2,350 | 15,800 | 1,510 |
| **I** | No chronic HCV hazard | 22,100 | 18,100 | 6,700 | 2,550 | 15,600 | 0 |
| **Northern Ireland** | | | | | | | |
| **Baseline** | | 730 | 599 | 221 | 72 | 527 | 52 |
| **A** | Past IDUs; later (date) and lower (%) deferral | 517 | 424 | 157 | 53 | 371 | 36 |
| **B** | No non-IDUs | 752 | 617 | 228 | 74 | 542 | 53 |
| **C** | All IDUs | 664 | 544 | 201 | 65 | 479 | 47 |
| **D** | Deferral one year later (date) | 769 | 631 | 233 | 76 | 555 | 55 |
| **E** | Lower deferral effect (%) | 530 | 434 | 161 | 54 | 381 | 37 |
| **F** | 50% contribution from IDUs | 708 | 581 | 215 | 69 | 511 | 50 |
| **G** | No additional transfusion | 730 | 599 | 221 | 78 | 521 | 50 |

| | | | | | | | |
|---|---|---|---|---|---|---|---|
| | hazards beyond 10 years | | | | | | |
| **H** | Non-age-stratified transfusion hazards | 730 | 599 | 221 | 76 | 523 | 50 |
| **I** | No chronic HCV hazard | 730 | 599 | 221 | 81 | 516 | 0 |
| **Scotland** | | | | | | | |
| **Baseline** | | 2,717 | 2,228 | 823 | 267 | 1,961 | 193 |
| **B** | No non-IDUs | 2,252 | 1,847 | 683 | 230 | 1,617 | 157 |
| **C** | All IDUs | 4,111 | 3,371 | 1,246 | 380 | 2,991 | 300 |
| **D** | Deferral one year later (date) | 2,859 | 2,344 | 867 | 281 | 2,063 | 203 |
| **E** | Lower deferral effect (%) | 2,402 | 1,970 | 728 | 240 | 1,730 | 169 |
| **F** | 50% contribution from IDUs | 3,182 | 2,609 | 964 | 305 | 2,304 | 228 |
| **G** | No additional transfusion hazards beyond 10 years | 2,717 | 2,228 | 823 | 294 | 1,934 | 183 |
| **H** | Non-age-stratified transfusion hazards | 2,717 | 2,228 | 823 | 285 | 1,943 | 186 |
| **I** | No chronic HCV hazard | 2,717 | 2,228 | 823 | 310 | 1,918 | 0 |
| **Wales** | | | | | | | |
| **Baseline** | | 1,322 | 1,084 | 401 | 129 | 954 | 94 |
| **A** | Past IDUs; later (date) and lower (%) deferral | 936 | 768 | 284 | 95 | 673 | 65 |
| **B** | No non-IDUs | 1,362 | 1,116 | 413 | 133 | 983 | 97 |
| **C** | All IDUs | 1,202 | 986 | 364 | 118 | 868 | 86 |
| **D** | Deferral one year later (date) | 1,393 | 1,142 | 422 | 137 | 1,005 | 99 |
| **E** | Lower deferral effect (%) | 959 | 787 | 291 | 97 | 690 | 67 |
| **F** | 50% contribution from IDUs | 1,282 | 1,051 | 389 | 125 | 926 | 91 |
| **G** | No additional transfusion hazards beyond 10 years | 1,322 | 1,084 | 401 | 140 | 944 | 90 |
| **H** | Non-age-stratified | 1,322 | 1,084 | 401 | 137 | 947 | 91 |

| | | | | | | | |
|---|---|---|---|---|---|---|---|
| | transfusion hazards | | | | | | |
| **I** | No chronic HCV hazard | 1,322 | 1,084 | 401 | 149 | 935 | 0 |

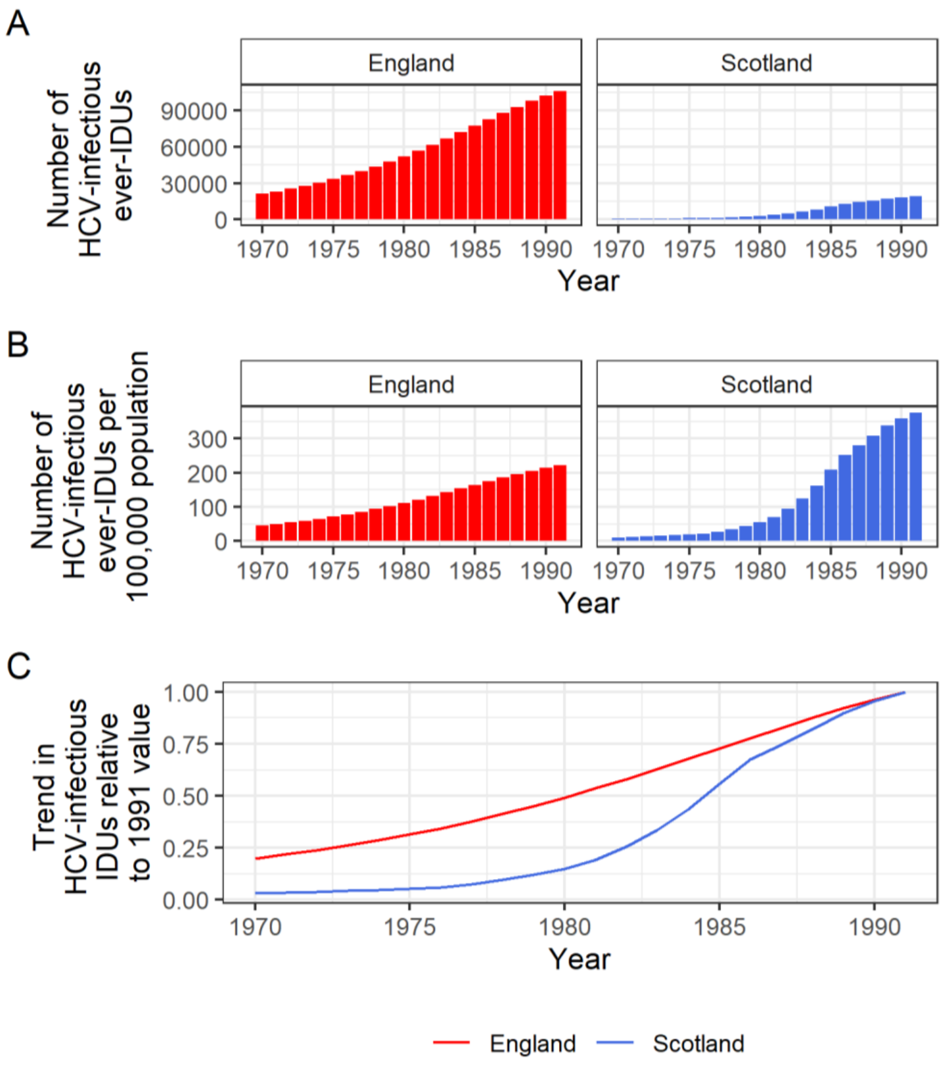

*Supplementary Figure 10: (A) The number of HCV-infectious ever-IDUs in England and Scotland. (B) As in (A) but scaled to per 100,000 population. (C) The trend in HCV-infectious ever-IDUs relative to the value observed in 1991.*

Supplementary Table 52 presents the estimated number of survivors to the end of 2014 and 2019 for males and females, respectively.

*Supplementary Table 52: Estimates from baseline stochastic model for England for the number, sex and age-distribution of survivors from chronic HCV infection by transfusion to 31 December 2014 and to 31 December 2019. Estimates are the median and 95% uncertainty interval and are rounded to the nearest 10. Note that no individual transfused up to August 1991 would have been aged 20 – 29 in 2019.*

| **Age band (in completed years) at 31 December 2014** | **Male survivors** | **Female survivors** | **Age band in completed years at 31 December 2019** | **Male survivors** | **Female survivors** |
|---|---|---|---|---|---|
| **20-29** | 30 (20 – 50) | 20 (20 – 30) | **20-29** | NA | NA |
| **30-39** | 220 (150 – 330) | 120 (80 – 190) | **30-39** | 120 (80 – 160) | 60 (40 – 90) |
| **40-49** | 170 (120 – 280) | 200 (140 – 290) | **40-49** | 220 (150 – 380) | 150 (100 – 240) |
| **50-59** | 130 (90 – 210) | 420 (310 – 630) | **50-59** | 130 (90 – 200) | 280 (220 – 380) |
| **60-69** | 170 (110 – 260) | 440 (320 – 680) | **60-69** | 140 (100 – 220) | 450 (330 – 700) |
| **70-79** | 130 (80 – 200) | 270 (190 – 420) | **70-79** | 130 (90 – 210) | 320 (230 – 500) |
| **80+ years** | 60 (30 – 110) | 140 (80 – 240) | **80+ years** | 60 (30 – 110) | 150 (90 – 270) |
| **Total** | **910** (670 – 1,350) | **1,600** (1,220 – 2,370) | **Total** | **800** (590 – 1,190) | **1,410** (1,070 – 2,090) |
